# A randomised clinical trial of azithromycin versus standard care in ambulatory COVID-19 – the ATOMIC2 trial

**DOI:** 10.1101/2021.04.21.21255807

**Authors:** Timothy SC Hinks, Lucy Cureton, Ruth Knight, Ariel Wang, Jennifer L Cane, Vicki S Barber, Joanna Black, Susan J Dutton, James Melhorn, Maisha Jabeen, Phil Moss, Rajendar Garlapati, Tanya Baron, Graham Johnson, Fleur Cantle, David Clarke, Samer Elkhodair, Jonathan Underwood, Daniel Lasserson, Ian D Pavord, Sophie Morgan, Duncan Richards

## Abstract

**Background:** The antibacterial, anti-inflammatory and antiviral properties of azithromycin suggest therapeutic potential against COVID-19. Randomised data in mild-moderate disease are lacking. We assessed whether azithromycin is effective in reducing hospitalisation in patients with mild-moderate COVID-19.

**Methods:** This open-label, randomised superiority clinical trial at 19 centres in the United Kingdom enrolled adults, ≥18 years, presenting to hospitals with clinically-diagnosed highly-probable or confirmed COVID-19 infection, with <14 days symptoms, considered suitable for initial ambulatory management. Patients were randomised (1:1) to azithromycin (500 mg daily orally for 14 days) or to standard care without macrolides. The primary outcome was the difference in proportion of participants with death or hospital admission from any cause over the 28 days from randomisation, assessed according to intention-to-treat (ITT). Trial registration: ClinicalTrials.gov, NCT04381962, Study closed.

**Findings:** 298 participants were enrolled from 3^rd^ June 2020 to 29^th^ January 2021. The primary outcome was assessed in 292 participants. The primary endpoint was not significantly different between the azithromycin and control groups (Adjusted OR 0·91 [95% CI 0·43-1·92], p=0·80). Rates of respiratory failure, progression to pneumonia, all-cause mortality, and adverse events, including serious cardiovascular events, were not significantly different between groups.

**Interpretation:** In patients with mild-moderate COVID-19 managed without hospital admission, adding azithromycin to standard care treatment did not reduce the risk of subsequent hospitalisation or death. Our findings do not support the use of azithromycin in patients with mild-moderate COVID-19.

**Funding:** NIHR Oxford BRC, University of Oxford and Pfizer Inc.

**Research in context:** *Evidence before this study:* We searched MEDLINE and the Cochrane Central register of Controlled Trials (CENTRAL) with the terms (“azithromycin”) AND (“COVID” OR “COVID-19”) AND (“clinical trials”), until March 25, 2021, with no language restrictions. We identified 42 studies, among which there were four completed randomised trials of azithromycin (with or without hydroxychloroquine) in hospitalised patients with severe disease, and three completed randomised trials of azithromycin in mild COVID-19 in primary care. The four trials in hospitalised patients randomised 8,988 participants to azithromycin or standard care or hydroxychloroquine and found no evidence of a difference in mortality, duration of hospital stay or peak disease severity. Of the three trials in primary care, these randomised participants with early disease to 3 or 5 days of therapy, of which only one assessed azithromycin as standalone therapy. This large, adaptive platform trial in the UK randomised 540 participants in primary care to 3 days treatment with azithromycin versus 875 to standard care alone and found no meaningful difference in time to first reported recovery, or of rates of hospitalisation (3% versus 3%) and there were no deaths. We did not identify any randomised trials in patients with COVID-19 managed in ambulatory care.

*Added value of this study:* The ATOMIC2 trial was uniquely-designed to assess azithromycin as a standalone therapy in those with mild-moderately COVID-19 presenting to emergency care, but assessed as appropriate for initial ambulatory management without hospital admission. ATOMIC2 also uniquely assessed high-dose, long-duration treatment to investigate the efficacy of putative anti-inflammatory effects. We found that azithromycin 500 mg daily for 14 days did not reduce the proportion of participants who died or required hospital admission from any cause over the 28 days from randomisation.

*Implications of all the available evidence:* Our findings, taken together with existing data, suggest there is no evidence that azithromycin reduces hospitalisation, respiratory failure or death compared with standard care, either in early disease in the community, or those hospitalised with severe disease, or in those with moderate disease managed on an ambulatory pathway.

## Introduction

Azithromycin (AZM) is an orally-active synthetic macrolide antibiotic with a wide range of antibacterial, anti-inflammatory and antiviral properties^1^. Early in 2020 it was highlighted by *in silico* and *in vitro*^2^ screens as a potential candidate therapy to be repurposed for treatment of COVID-19. Indeed macrolides, particularly azithromycin, had previously been used to treat other viral infections including one in three severe cases of MERS-CoV^3^ although randomised controlled trial data for its use in any coronavirus disease were lacking^4^. Azithromycin is inexpensive, safe and widely available, and stimulated by a small, non-randomised clinical report^5^ its use in the context of COVID-19 has become widespread in clinical practice and a number of clinical trials.

*In vitro* azithromycin has broad antiviral activity against human viruses including human rhinovirus, Zika, enteroviruses, Ebola, SARS^1^, and against SARS-CoV-2, being shown to reduce viral replication alone^2^ or in combination with hydroxychloroquine^6^. Azithromycin was also associated with reduced viral load of non-SARS-CoV-2 alpha- and beta-coronaviruses in children receiving azithromycin during a mass distribution programme^7^. Although antivirals are likely to have limited efficacy in severe disease after viraemia has peaked^8-10^ azithromycin also has anti-inflammatory properties including dose-dependent suppression of lymphocyte expression of perforin, and a range of proinflammatory cytokines, including IL-1β, IL-6 and TNF, IL-8(CXCL8), IL-18, G-CSF and GM-CSF^11,12^ and other components of the IL-1β,/IL-6-induced acute phase response such as serum amyloid protein A^12^.

Despite these theoretical considerations, large scale clinical trials of azithromycin either alone or co-administered with hydroxychloroquine have not shown clinical efficacy in reducing mortality, need for invasive mechanical ventilation, duration of hospitalisation, or clinical status on ordinal outcome scores amongst patients hospitalised with COVID-19^13-16^. However, these trials were all conducted in late-stage, severe disease with 17-40% mortality. They did not study patients at earlier stages of disease in the community and are not able to make conclusions about the effectiveness of azithromycin outside the hospital setting. The efficacy of therapies in COVID-19 depends on the timing in the course of disease and in the populations being studied. Dexamethasone showed a strong survival benefit with treatment in severe patients but no benefit, or even potential harm, in those not requiring oxygen therapy^17^. Conversely studies of neutralising antibodies showed benefits in early disease^18^ but not in hospitalised patients^19^. The antiviral and anti-inflammatory properties of azithromycin are suited to earlier stage disease, thus we studied an ambulatory population to establish whether it averts disease progression.

We conducted a randomised, open-label clinical trial to determine whether azithromycin is effective in preventing hospitalisation or death in adult patients with clinically-diagnosed COVID-19 infection being managed on an ambulatory care pathway.

## Methods

### Study design

ATOMIC2 was a prospective, open-label, two-arm, randomised superiority clinical trial of standard care and Azithromycin with standard care alone, conducted at 19 hospitals in the United Kingdom. The trial was conducted according to the published protocol v7.0^20^ (Appendix 1) and all applicable laws and regulations including, but not limited to, the principles as stated in the International Council for Harmonisation Guideline for Good Clinical Practice, the standards set out by the Research Governance Framework, the Medicines for Human Use (Clinical Trials) Regulations 2004, and the ethical principles that have their origin in the Declaration of Helsinki. Safety data were reviewed and monitored by an independent data safety monitoring committee. The trial protocol was reviewed and approved by the UK Medicines and Healthcare products Regulatory Agency and an independent ethical committee (London – Brent Research Ethics Committee, Research Ethics Committee reference number 20/HRA/2105).

### Participants

Eligible participants were adults, ≥18 years of age assessed in an acute hospital with a clinical diagnosis of highly-probable or confirmed COVID-19 infection made by the attending clinical team, with onset of first symptoms within the last 14 days, and assessed by the attending clinical team as appropriate for initial ambulatory (outpatient) management. Key exclusion criteria included known hypersensitivity to any macrolide or the excipients, fructose intolerance, glucose-galactose malabsorption or sucrose-isomaltase-insufficiency, current therapy with a macrolide antibiotic, hydroxychloroquine or chloroquine, significant myocarditis, prolongation of a corrected QT interval (QTc) >480 msec, significant electrolyte disturbance, clinically relevant bradycardia, ventricular tachycardia, unstable severe cardiac insufficiency, and inability to understand written English. The full protocol including a complete list of inclusion / exclusion criteria are provided in appendix 1^20^. All patients provided electronic informed consent before randomisation.

### Randomisation and masking

Patients were randomly assigned (1:1) to either azithromycin plus standard care or standard care alone using a web-based automated service, with a minimisation algorithm to ensure balanced allocation across treatment groups, stratified by centre, sex and presence of hypertension and diabetes. To ensure the unpredictability of treatment allocation the first 30 participants were randomised by simple randomisation and the minimisation algorithm included a probabilistic element (participants had an 80% chance of being allocated to the treatment which minimised imbalance between the groups). Patients, investigators, and health-care providers were not masked to study drug assignment.

### Procedures

Data on demographics, medical history, symptomatology, risk factors for disease progression and disease severity were collected on all patients at baseline. Results were recorded for vital signs (temperature, respiratory rate, heart rate, blood pressure, and oxygen saturation), physical examination (including chest auscultation) with clinical assessment including the COVID Core Outcomes Set^21^, and a nine-level severity score of respiratory illness (0-8, where 0 indicates “Ambulatory. No limitation of activities” and 8 indicates “Death”). Participants also had a 12-lead electrocardiogram. Optional study samples were taken at baseline and on one further occasion if the participant was admitted: oropharyngeal swab for SARS-CoV-2 PCR, nasal and blood samples for RNA transcriptomic analysis. Bloods and chest x-rays were done if clinically required.

Patients in the azithromycin group received 500 mg azithromycin once daily orally plus standard care for 14 days and those in the control group received standard care according to local guidelines. Use of corticosteroids, other immunomodulators, antibiotics, and antivirals was permitted after randomisation in the control group, but the protocol excluded concomitant use of quinolone or macrolides antibiotics at enrolment or during follow-up. Subsequent assessments were carried out by telephone at days 14 and 28, and radiology results and review of notes daily during hospital admission if this occurred.

### Outcomes

The primary outcome was the proportion of participants with hospital admission or death from any cause within 28 days from randomisation.

Secondary outcomes included the proportion of participants with hospital admission with respiratory failure requiring Non-Invasive Mechanical Ventilation (NIV) or Invasive Mechanical Ventilation (IMV) or death from any cause within 28 days from randomisation; the proportion of participants with hospital admission with respiratory failure requiring IMV or non-IMV support or death from any cause over 28 days from randomisation amongst those with a PCR-confirmed diagnosis of COVID-19 at randomisation; mortality or all-cause hospital admission amongst those with a PCR-confirmed diagnosis; all-cause mortality; the proportion progressing to clinician-diagnosed pneumonia; the proportion progressing to severe pneumonia; differences in the peak severity of illness according to a nine-level ordinal severity score for clinical condition described in the protocol (Appendix 1). We also assessed safety and tolerability of azithromycin based on adverse events.

### Statistical analysis and Protocol changes

We had originally planned for the trial to recruit up to 800 participants and use the following primary outcome: the proportion participants with hospital admission with respiratory failure requiring Non-Invasive Mechanical Ventilation (NIV) or Invasive Mechanical Ventilation (IMV) or death from any cause within 28 days from randomisation. However, a decision was made at the pre-planned interim analysis of 109 participants reaching the 28-day post-randomisation time-point to update the primary outcome as no primary outcomes had at that stage occurred. This implied that the original primary outcome, specified early in the pandemic when data on hospitalisation rates in this population were unknown, proved incorrect for this population. The change was adopted in line with advice from the data safety monitoring committee and in accordance with the recommendations of the World Health Organisation Blueprint for Covid-19 Therapeutic Trials^22^ that the primary endpoint should be responsive to the eligible patient population and the definition of the endpoint should be fine-tuned for the Pivotal Phase, based on the Pilot Phase of the Trial. This change was approved by the research ethics committee and the MHRA on February 4^th^ 2021 and implemented before final analyses were performed. Two other protocol amendments were performed while the trial was ongoing to broaden inclusion criteria to include and ensure the safety of participants taking serotonin-specific reuptake inhibitors (Appendix 1).

Based on WHO recommendations that a pilot phase with 100 patients would be sufficient to inform follow-on clinical research^22^ an interim analysis was planned to determine definitive sample size. Following the revision to the primary outcome and based on blinded data from the pilot phase, the definitive sample size was determined. Assuming a 15% rate of all cause hospitalisation or death in the standard care arm, we estimated a minimum of 276 participants providing primary end-point data, would provide 80% power and 5% (2-sided) significance to detect a difference from 15% to 5% in the Azithromycin arm, a relative reduction of 66%. To allow for 5% loss to follow-up we therefore aimed to recruit a minimum of 291 participants. The full statistical analysis plan is provided in Appendix 2.

For the primary outcome the difference in proportions between the treatment arms was assessed using a chi-squared test and a 5% (2-sided) significance level. Adjusted analysis was undertaken using logistic regression with progression as the binary outcome, adjusting for stratification factors: centre, hypertension, diabetes and sex. A supporting analysis was also undertaken to further adjust for other important prognostic variables: age ≥65 years, presence of chronic lung disease, and treatment for cancer. Time to event analysis was also undertaken to explore whether the active treatment delays progression. The success of the trial was based on the adjusted analysis. Both relative and absolute differences in proportions are reported together with 95% confidence intervals. Other binary outcomes were assessed using similar methods. Peak severity of illness was considered a categorical variable and assessed using ordinal logistic regression analysis. The change in severity scale score from baseline was summarised on a continuous scale using means, standard deviations, medians, interquartile ranges and ranges. Analyses were performed using Stata IC V15.1 (StataCorp LP, www.stata.com).

Efficacy and safety analyses were based on the intention-to-treat (ITT) population, defined as all randomised patients analysed according to their randomised allocation. A supplementary ITT population (ITT +ve) was defined as all randomised patients with a positive baseline COVID-19 test based on baseline swabs. This trial was registered with ClinicalTrials.gov (NCT04381962) and EudraCT (2020-001740-26).

## Results

From 3^rd^ June 2020 to 29^th^ January 2021 1192 patients were screened, of whom 649 were ineligible, 84 declined consent, 161 were excluded for other reasons and 298 were enrolled in the trial (Figure 1, and Appendix 3 Table S1). Three participants withdrew consent and requested removal of all data collected so are not presented in baseline data. 295 of the 298 randomised participants (99%) were included in the ITT population, of whom 147 were randomised to azithromycin plus standard care and 148 to standard care alone. Three of the 295 remaining patients withdrew consent after randomisation (Appendix 3 Table S9). Data on the primary outcome were available from 292 participants.

**Figure 1:**
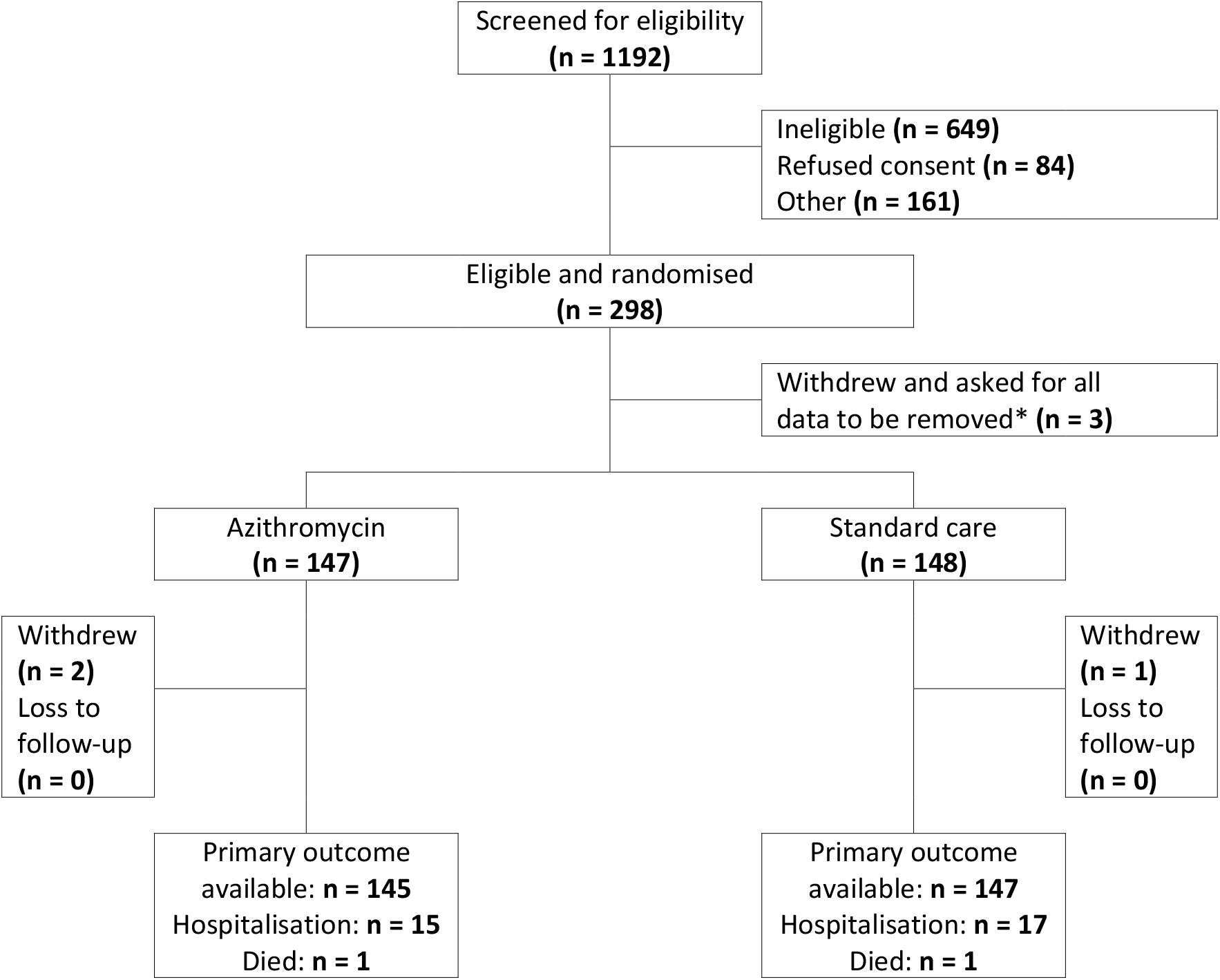
Trial profile. *These participants withdrew completely and asked for all of their data collected to date to be removed. Therefore, they have not been included in any further summaries or analyses.

Characteristics of participants assigned to azithromycin and to standard care were similar (Table 1, and Appendix 3 Tables S2 to S7). The mean participant age was 45·9 years (SD 14·9) and 51·5% were male. 68·1% were of White ethnicity, 15·9% Asian or Asian/British, 3·7% Black or Black/British, and 12·2% of Mixed or Other ethnicity. 70 (24·6%) of 284 participants had comorbidities, and the median duration of symptoms before enrolment was 6·02 (3·52) days. Enrolment was based on a clinical diagnosis of highly-probable Covid-19, but the definitive results of nasopharyngeal swabs for SARS-CoV-2 PCR were available from 231 individuals, of which 152/231 (65·8%) were positive, and constituted the ITT +ve population, of whom 76 were randomised to azithromycin (50%) and 76 to standard care alone (50%). 143/147 participants allocated to azithromycin (97·3%) commenced treatment, with 76 (51· 7%) achieving full compliance (taking a median 28 tablets), 51 (34·7%) non-compliant, taking a median 6 tablets (IQR 2 to 17), and compliance unknown in 20 (13·6%)(Appendix 3 Table S8).

**Table 1:** Baseline characteristics and concomitant treatments of the intention-to-treat population

|  | Azithromycin (n = 147) | Standard Care (n = 148) | Total (n = 295) |
| --- | --- | --- | --- |
| **Hypertension<sup>1</sup>** |  |  |  |
| Yes | 25 (17.0%) | 27 (18.2%) | 52 (17.6%) |
| No | 122 (83.0%) | 121 (81.8%) | 243 (82.4%) |
| **Diabetes<sup>1</sup>** |  |  |  |
| Yes | 11 (7.5%) | 14 (9.5%) | 25 (8.5%) |
| No | 136 (92.5%) | 134 (90.5%) | 270 (91.5%) |
| **Gender<sup>1</sup>** |  |  |  |
| Male | 76 (51.7%) | 76 (51.4%) | 152 (51.5%) |
| Female | 71 (48.3%) | 72 (48.6%) | 143 (48.5%) |
| **Age(years)<sup>2</sup>** | 45.53 (14.23) | 46.30 (15.53) | 45.92 (14.88) |
| **COS<sup>2,3</sup>** | 6.36 (3.64) | 7.00 (3.87) | 6.68 (3.77) |
| **COSTotalPlus<sup>2,3</sup>** | 7.66 (4.65) | 8.86 (5.25) | 8.26 (4.98) |
| **Duration of symptoms (days)<sup>2</sup>** | 5.77 (3.49) | 6.27 (3.55) | 6.02 (3.52) |
| **Ethnicity<sup>1</sup>** |  |  |  |
| White | 103 (70.1%) | 98 (66.2%) | 201 (68.1%) |
| Mixed | 0 (0.0%) | 4 (2.7%) | 4 (1.4%) |
| Asian/ Asian British | 23 (15.6%) | 24 (16.2%) | 47 (15.9%) |
| Black/ Black British | 6 (4.1%) | 5 (3.4%) | 11 (3.7%) |
| Other Ethnic Group | 15 (10.2%) | 17 (11.5%) | 32 (10.8%) |
| **Smoking<sup>1</sup>** |  |  |  |
| Never smoked | 81 (57.4%) | 76 (53.1%) | 157 (55.3%) |
| Ex-smoker | 25 (17.7%) | 26 (18.2%) | 51 (18.0%) |
| Current smoker | 16 (11.3%) | 17 (11.9%) | 33 (11.6%) |
| Ex-smoker & current vaper | 3 (2.1%) | 4 (2.8%) | 7 (2.5%) |
| Never smoked and current vaper | 0 (0.0%) | 1 (0.7%) | 1 (0.4%) |
| Not recorded | 16 (11.3%) | 19 (13.3%) | 35 (12.3%) |
| **Residence<sup>1</sup>** |  |  |  |
| Non-residential care | 132 (91.7%) | 137 (95.1%) | 269 (93.4%) |
| Residential care | 7 (4.9%) | 3 (2.1%) | 10 (3.5%) |
| No fixed address | 5 (3.5%) | 4 (2.8%) | 9 (3.1%) |
| **Live alone<sup>1</sup>** |  |  |  |
| Yes | 17 (13.6%) | 13 (10.6%) | 30 (12.1%) |
| No | 108 (86.4%) | 110 (89.4%) | 218 (87.9%) |
| **Work Status<sup>1</sup>** |  |  |  |
| Retired | 15 (10.9%) | 23 (16.5%) | 38 (13.7%) |
| Working | 101 (73.2%) | 95 (68.3%) | 196 (70.8%) |
| Houseperson | 22 (15.9%) | 21 (15.1%) | 43 (15.5%) |
| **Occupation<sup>1</sup>** |  |  |  |
| Not Healthcare related | 77 (78.6%) | 69 (74.2%) | 146 (76.4%) |
| Healthcare worker | 20 (20.4%) | 23 (24.7%) | 43 (22.5%) |
| Laboratory worker | 1 (1.0%) | 1 (1.1%) | 2 (1.0%) |
| **Total number of comorbidities<sup>1</sup>** |  |  |  |
| No comorbidities | 107 (76.4%) | 107 (74.3%) | 214 (75.4%) |
| ≥1 comorbidity | 33 (23.6%) | 37 (25.7%) | 70 (24.6%) |
| **Have asthma<sup>1</sup>** |  |  |  |
| Yes | 26 (17.7%) | 27 (18.2%) | 53 (18.0%) |
| No | 121 (82.3%) | 121 (81.8%) | 242 (82.0%) |
| **History of previous myocardial infarction<sup>1</sup>** |  |  |  |
| Yes | 5 (3.4%) | 7 (4.7%) | 12 (4.1%) |
| No | 142 (96.6%) | 141 (95.3%) | 283 (95.9%) |
| **Currently undergoing any cancer treatment<sup>1</sup>** |  |  |  |
| Yes | 1 (0.7%) | 0 (0.0%) | 1 (0.3%) |
| No | 146 (99.3%) | 148 (100.0%) | 294 (99.7%) |
| **Have chronic pulmonary disease<sup>1</sup>** |  |  |  |
| Yes | 7 (4.8%) | 5 (3.4%) | 12 (4.1%) |
| No | 140 (95.2%) | 143 (96.6%) | 283 (95.9%) |
| **The severity scale score<sup>1,4</sup>** |  |  |  |
| Ambulatory, no limitation of activities. | 61 (41.5%) | 66 (44.6%) | 127 (43.1%) |
| Limitation of simple activities | 85 (57.8%) | 81 (54.7%) | 166 (56.3%) |
| Hospitalised, mild disease, no oxygen therapy | 1 (0.7%) | 1 (0.7%) | 2 (0.7%) |
| **Pneumonia<sup>1,5</sup>** |  |  |  |
| Yes | 28 (19.0%) | 34 (23.0%) | 62 (21.0%) |
| No | 119 (81.0%) | 114 (77.0%) | 233 (79.0%) |
| Not available | 0 (0.0%) | 0 (0.0%) | 0 (0.0%) |
| **Swab results<sup>1,6</sup>** |  |  |  |
| Positive | 76 (51.7%) | 76 (51.4%) | 152 (51.5%) |
| Negative | 41 (27.9%) | 38 (25.7%) | 79 (26.8%) |
| Failed Assay | 0 (0.0%) | 3 (2.0%) | 3 (1.0%) |
| Not available | 30 (20.4%) | 31 (20.9%) | 61 (20.7%) |

<sup>1</sup>Summaries are n (%)
<sup>2</sup>Summaries are mean (SD)
<sup>3</sup> COVID-19 COS Score of clinical symptoms is a total score of six common and important clinical symptoms, each scored as 0 (no), 1 (mild), 2 (moderate), or 3 (significant). An amended version COVID-19 COS PLUS contains an additional two clinical symptoms: anosmia and dysgeusia. COS scores range from 0 to 18, COSPlus scores from 0 to 24, higher scores indicating more significant Covid symptoms.
<sup>4</sup> The severity scale scores range from 0 to 8 with higher scores indicating the most severe status, death.
<sup>5</sup> Note: Pneumonia is defined as 'consolidation on a chest X-ray', if a chest X-ray was not taken it is assumed there was no pneumonia.
<sup>6</sup>Note: SWAB test results are only available for those who had a Covid-19 swab at randomisation.

15 (10·3%) of 145 participants randomised to azithromycin and 17 (11·6%) of 147 participants randomised to standard care were hospitalised or died. The primary endpoint was not significantly different between the azithromycin and control groups (adjusted OR 0·91, 95% CI 0·43–1·92, p=0·80; Table 2 and Appendix 3 Table S10 to 12; Figure 2 and Appendix 3 Figure S1). There was no difference in the time to hospitalisation after adjusting for stratification factors (centre, hypertension, diabetes and sex) using logistic regression assessed by Cox’s proportional hazard ratio (HR) 0·95 (0·46 to 1·96, p=0·89; Appendix 3 Table S12; Figure 3a). There were also no differences in the combined primary outcome of hospitalisation or death (adjusted OR 1·02, 95% CI 0·40-2·57, p=0·97), or on time to hospitalisation (HR 1·17, 95% CI 0·49 to 2·77), p=0·72; Figure 3b) when these analyses were repeated on the ITT+ve population (Appendix 3 Table S12). In addition, unadjusted and fully adjusted analyses (further adjusted for age, chronic pulmonary disease and presence of cancer) were performed as well as analyses on the per protocol population. None of these demonstrated significant differences (Appendix 3 Tables S21 and S22).

**Table 2:**
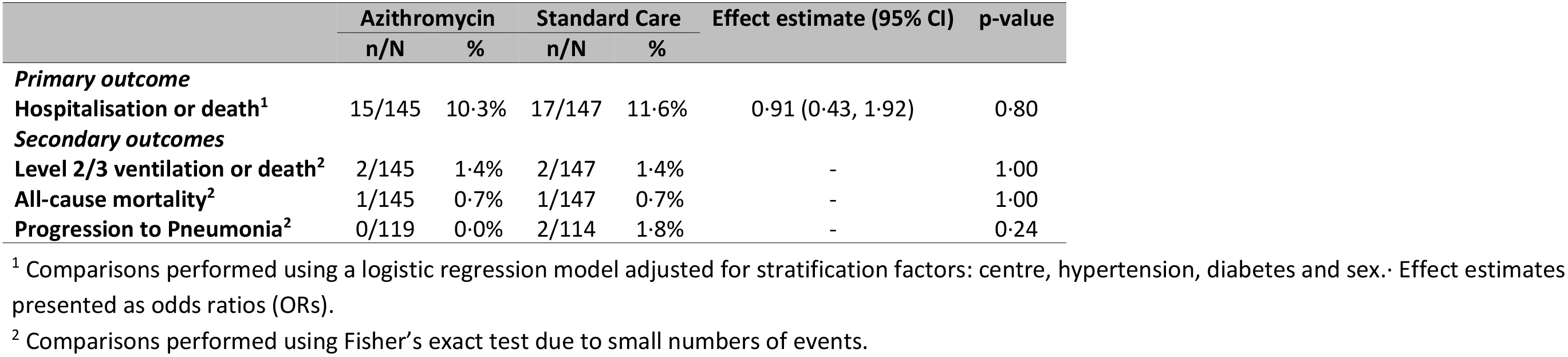
Comparison of primary and secondary binary outcomes in the intent-to-treat population

|  | Azithromycin |  | Standard Care |  | Effect estimate (95% CI) | p-value |
| --- | --- | --- | --- | --- | --- | --- |
|  | n/N | % | n/N | % |  |  |
| **Primary outcome** |  |  |  |  |  |  |
| **Hospitalisation or death<sup>1</sup>** | 15/145 | 10.3% | 17/147 | 11.6% | 0.91 (0.43, 1.92) | 0.80 |
| **Secondary outcomes** |  |  |  |  |  |  |
| **Level 2/3 ventilation or death<sup>2</sup>** | 2/145 | 1.4% | 2/147 | 1.4% | - | 1.00 |
| **All-cause mortality<sup>2</sup>** | 1/145 | 0.7% | 1/147 | 0.7% | - | 1.00 |
| **Progression to Pneumonia<sup>2</sup>** | 0/119 | 0.0% | 2/114 | 1.8% | - | 0.24 |

<sup>1</sup> Comparisons performed using a logistic regression model adjusted for stratification factors: centre, hypertension, diabetes and sex. Effect estimates presented as odds ratios (ORs).
<sup>2</sup> Comparisons performed using Fisher's exact test due to small numbers of events.

**Figure 2:**
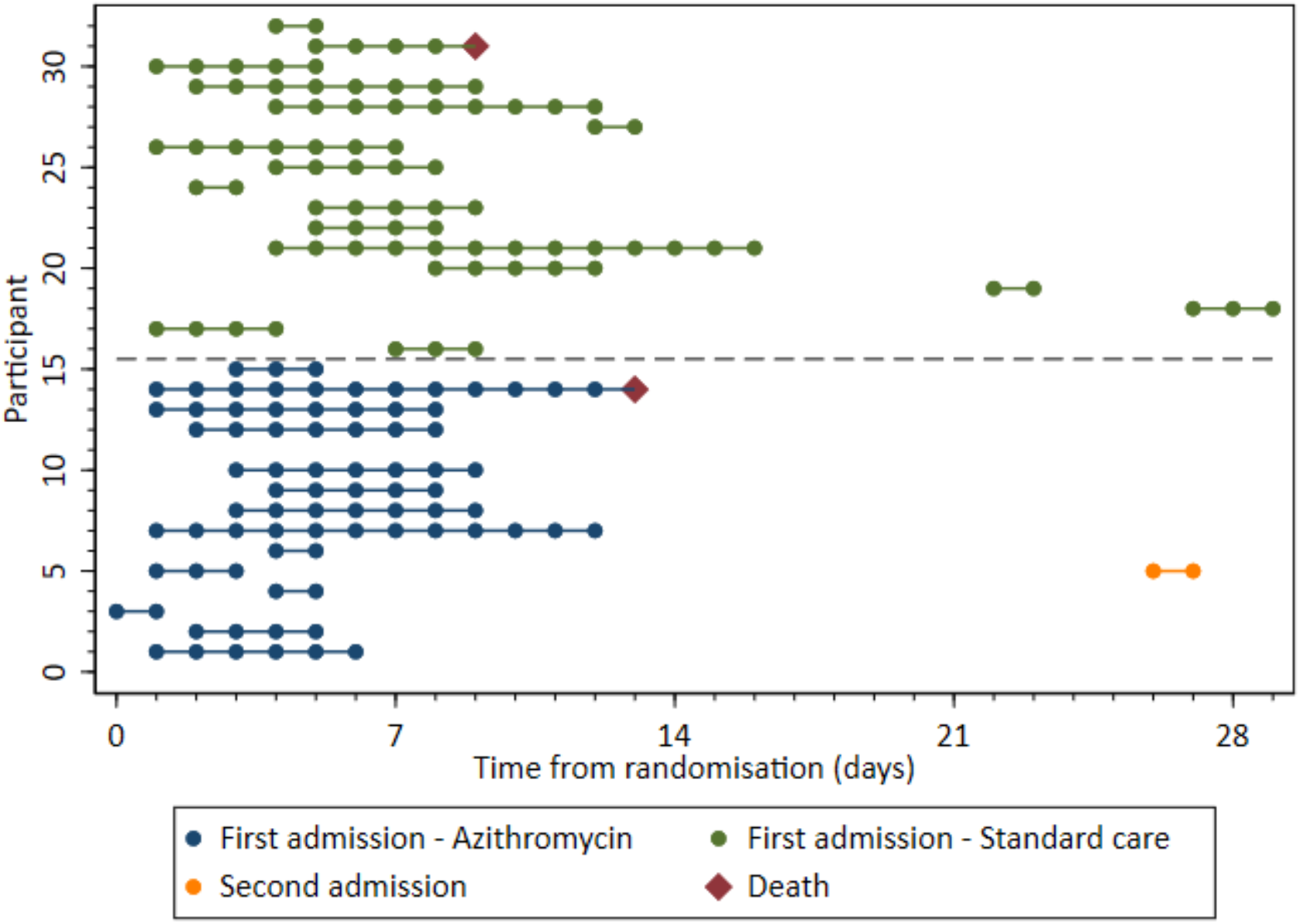
Time to hospitalisation, length of inpatient stay and time to death.

**Figure 3a:**
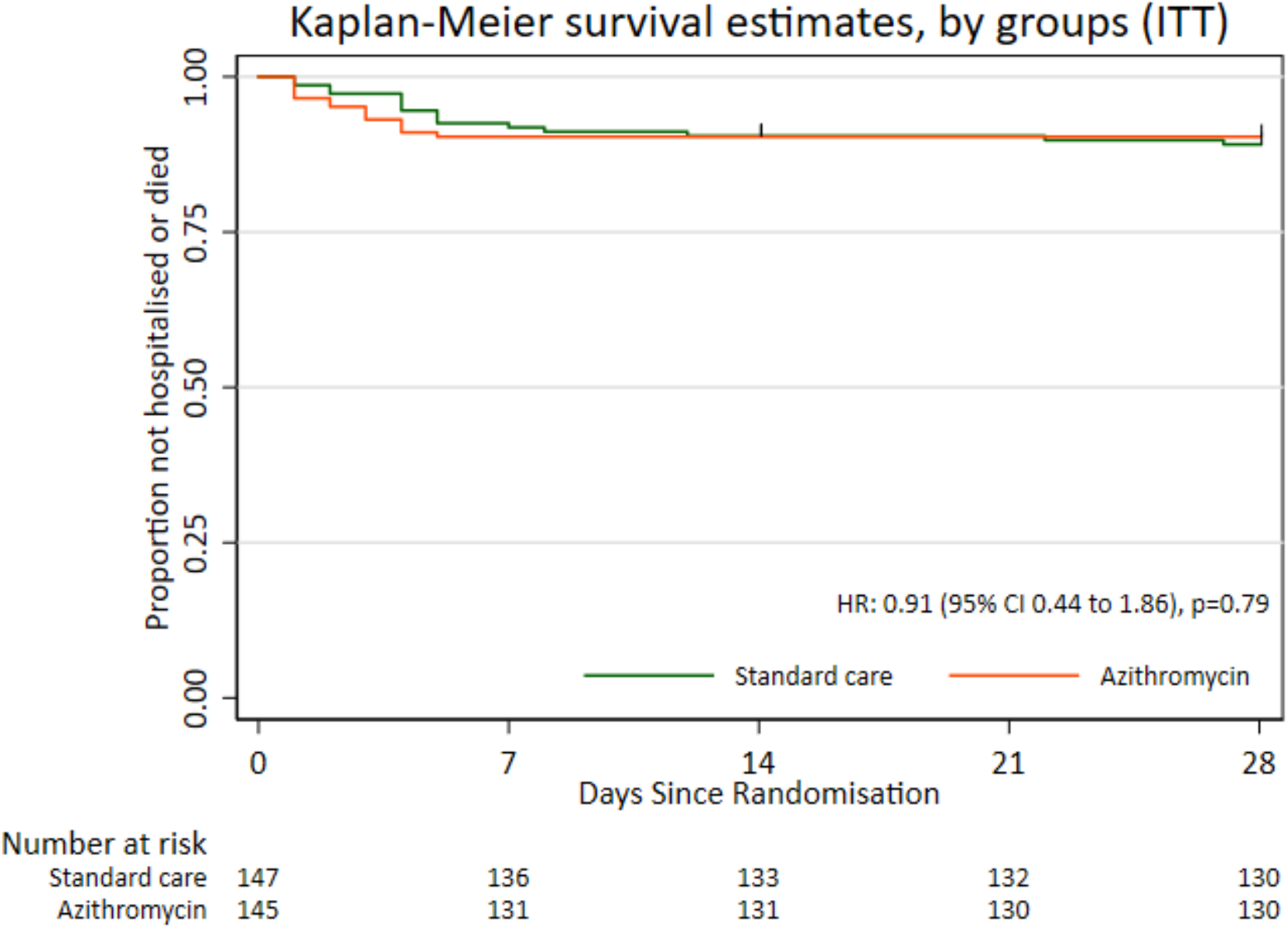
Kaplan-Meier plot of time to hospitalisation in the ITT population.

**Figure 3b:**
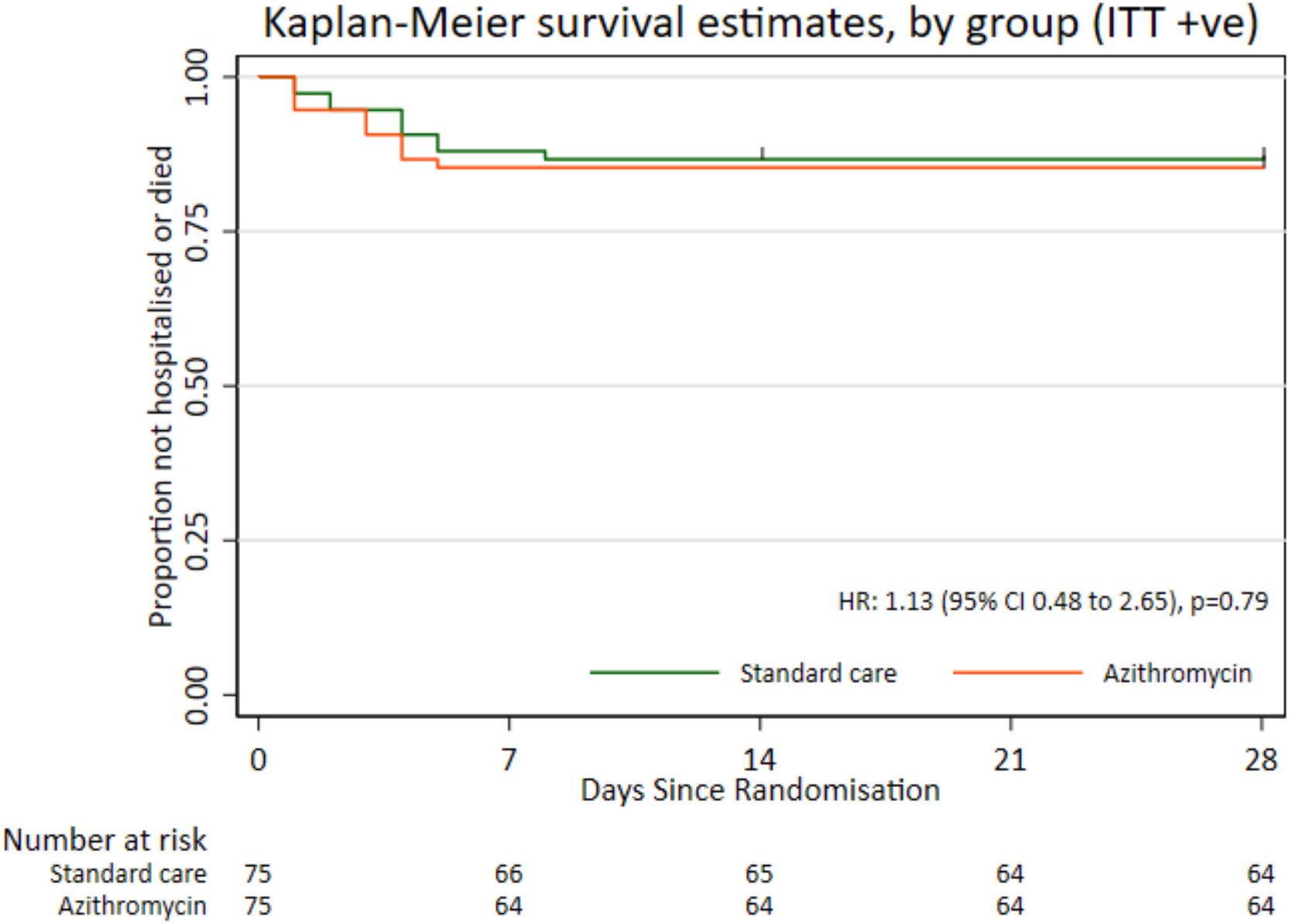
Kaplan-Meier plot of time to hospitalisation in the ITT +ve population.

Only two participants in each arm required level 2 or level 3 ventilation or died. Due to the very low number of events Fisher’s exact test was used to compare the azithromycin and control groups which were not significantly different (Table 2). The level of oxygen support required by each participant during hospitalisation is also summarised (Appendix 3 Figure S2). The number of both deaths from all-causes and participants progressing to pneumonia were similarly very low with no differences between the two groups identified (Table 2). No participants progressed to severe pneumonia during the trial. Full details of pneumonia status at baseline and follow-up is provided in Appendix 3 Table S14. Analyses of these outcomes were also repeated on the ITT +ve population with no significant differences identified (Appendix 3 Table S13).

Severity scores at baseline, days 14 and 28 are summarised in Figure 4 and Appendix 3 Table S15 and Figure S3. The peak severity score during follow-up was calculated for each participant. 62 (50·0%) of 124 participants randomised to azithromycin and 60 (46·2%) of 130 participants randomised to standard care reported no limitation of activities as their highest follow-up severity score (Table 3, Appendix 3 Tables S16 and S18 and Figure S4). There was not a significant difference between the two groups in terms of peak severity score (adjusted OR 0·91, 95% CI 0·57-1·46, p=0·69; Table 3). Analysis was repeated for the ITT +ve population with no significantly differences observed (Appendix 3 Table S17). The COVID-19 Core Outcome Set Plus (COS Plus) scores were similar in the two groups at both 14 days (7.62 in the azithromycin group versus 7·47 in the standard care group) and 28 days (3·06 in the azithromycin versus 3·38 in the standard care group) post-randomisation (Appendix 3 Table S19 and Figures S5 and S6).

**Figure 4:**
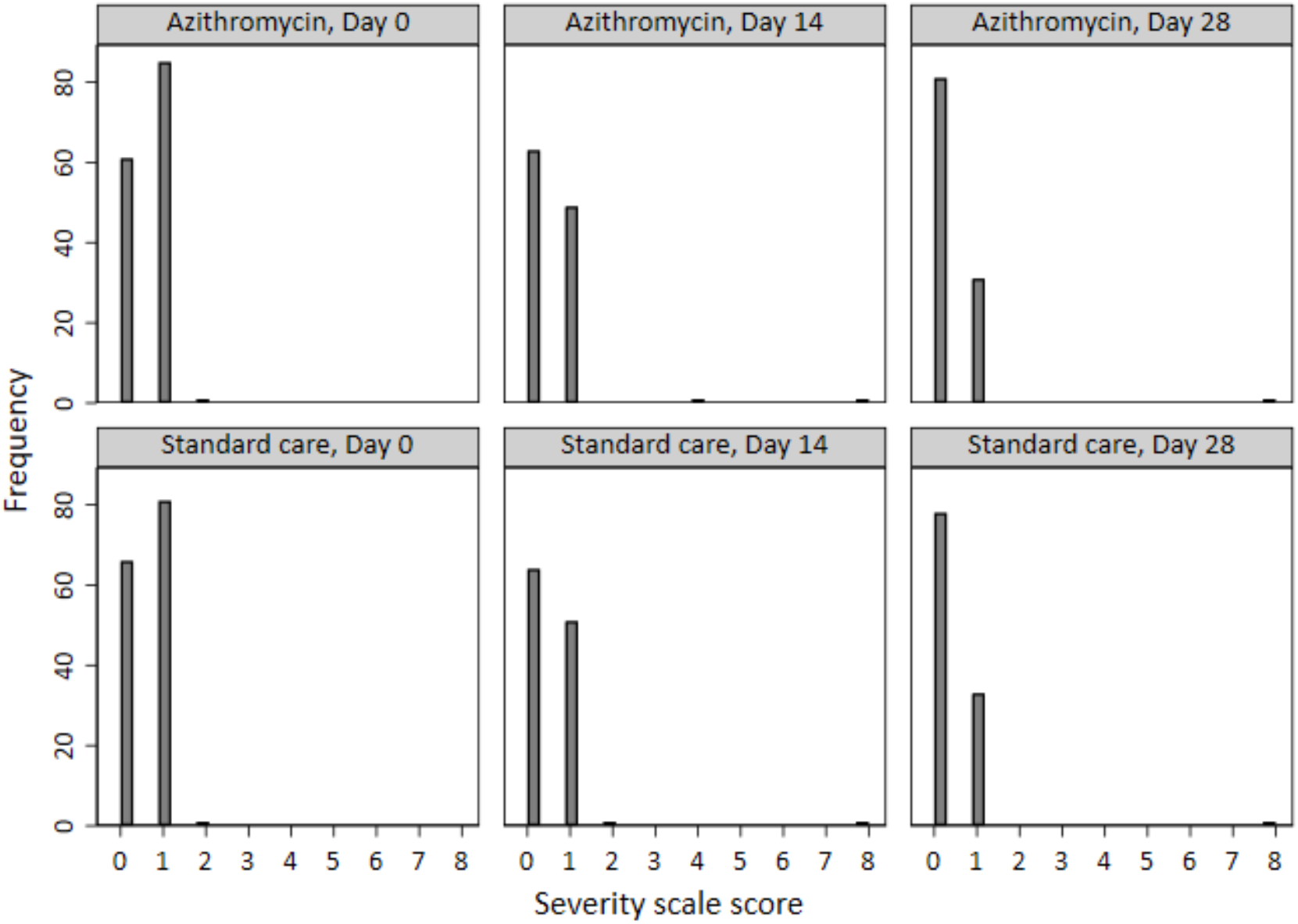
Severity scores at days 0, 14, 28.

**Table 3:**
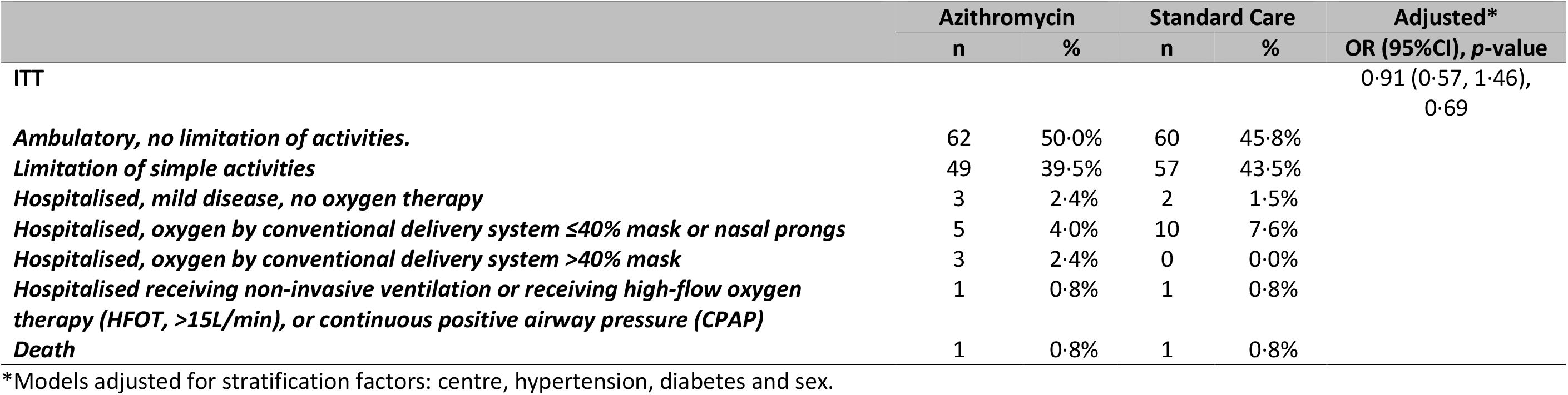
Comparison of peak severity scores in the intent-to-treat-population

During follow up additional antibiotics were co-prescribed in 23/147 (15·6%) participants in the azithromycin arm and 38/148 (25·7%) in the standard care arm (Appendix 3 Table S23). No SAEs were recorded in either treatment group during the course of follow-up. Three (2·1%) of 145 participants randomised to azithromycin and 4 (2·7%) of 147 participants randomised to standard care reported a complication during hospitalisation (Appendix 3 Table S20).

## Discussion

In this trial of people with clinically-diagnosed mild-moderate COVID-19 managed without hospital admission, adding azithromycin to standard care treatment did not reduce the risk of subsequent hospitalisation or death, or of time to hospitalisation.

Three recent, large, open-label, randomised controlled trials have assessed the use of azithromycin in patients hospitalised with severe COVID-19, and none have found a clinically-significant benefit in those populations. COALITION I randomised 667 patients hospitalised with COVID-19 to standard care, hydroxychloroquine or hydroxychloroquine with azithromycin 500mg for 7 days and found no difference in clinical status on an ordinal score at 15 days between hydroxychloroquine with azithromycin and hydroxychloroquine (odds ratio (OR), 0·82; 95% CI, 0·47 to 1·43; P=1·00)^16^. Likewise, COALITION II randomised 447 hospitalised patients to azithromycin 500 mg daily for 10 days or standard care, and again found no difference in day 15 ordinal score (OR 1·36, 95% CI 0·94–1·97, p=0·11).)^13^. RECOVERY randomised 7,763 of its participants to azithromycin 500 mg for 10 days or standard care and found no difference in 28-day mortality (rate ratio (RR) 0·97, 95% CI 0·87–1·07; p=0·50), length of stay, or invasive mechanical ventilation and death^15^. However, none of these trials assessed the potential for efficacy in early, milder disease.

Three trials in primary care have randomised participants with early disease to three^23^ or five^24,25^ days of therapy. Azithromycin was assessed as standalone therapy only in the PRINCIPLE trial^23^: a large, adaptive platform trial in the UK, which randomised 540 participants to 3 days treatment with azithromycin 500 mg daily versus 875 to standard care. This study found no difference in time to first reported recovery (hazard ratio (HR) 1·08, 95% Bayesian credibility interval 0·95 to 1·23), and, though only 3% of participants were hospitalised, there was no significant difference between groups (absolute benefit in 0·3%, 95% BCI –1·7 to 2·2). The remaining two trials also used short courses of azithromycin of 500 mg for one day followed by 250 mg for 4 days and taken in conjunction with hydroxychloroquine. Q-PROTECT recruited healthy, SARS-CoV-2-positive men in a quarantine site in Qatar and found no difference in time to virological cure (p=0·82)^24^, with low rates of hospitalisation in all groups (2·4%). A study in the USA assessed progression to lower respiratory tract infection, hospitalisation or death and time to viral clearance in SARS-CoV-2-positive outpatients, but was stopped early for futility due to a low rate of clinical outcomes in this population also, and found no difference in the co-primary outcome of time to virologic clearance (HR=1·25, 95% CI 0·75 to 2·07, p=0·39)^25^. No studies have assessed azithromycin in those presenting to hospital with significant symptoms, but early enough in the disease process to be managed in ambulant care, and neither have previous studies assessed high dose, long-duration azithromycin therapy in early disease.

Our study investigated this intermediate population with early disease, but at high risk of deterioration, in whom 10·6% required subsequent hospitalisation, and therefore represents a population with the optimal chance of demonstrating clinical benefit in early disease. This, taken together with clear negative results across the disease course from early, low risk patients, to severe hospitalised disease, provides strong confirmation that azithromycin is not effective in treating COVID-19.

A unique feature of ATOMIC2 was its successful implementation at the interface between community and secondary care – often a challenging location to recruit due to pressures for rapid clinical decision making – which was made possible by electronic screening, consent and recruitment. This platform, along with simplicity and broad inclusion criteria, have facilitated a second strength of the study: inclusion of an ethnically diverse population with 31·8% of participants recruited from Black and minority ethnic (BAME) backgrounds. This is important as COVID-19 is a global pandemic, with a disproportionate impact on BAME groups, yet these groups are underrepresented in many COVID-19 trials, which may reduce external validity^26^. Thirdly in contrast to other studies the high dose (500 mg daily) and long duration (14 days) of azithromycin was selected in our study to ensure that we adequately assessed potential antiviral, antibacterial and anti-inflammatory benefits. COVID-19 is considered to have a distinct early viraemic phase and a late inflammatory phase in some individuals, and therefore assessment of antiviral activity needs to be early in the disease course prior to onset of severe disease^20^. Conversely it was not known what doses might be required to produce an adequate anti-inflammatory effect and so it was necessary to give a high dose of long duration to ensure the anti-inflammatory effect was tested throughout the late stage of innate/ acute phase inflammatory cytokine dysregulation^1^. An additional strength is that we were also able to exclude a significant benefit from azithromycin’s antibiotic effects, which was not possible in hospitalised studies where co-prescribing of !-lactam and other antibiotics was common^15^. Our data show that secondary bacterial infection is not a major driver of admission in this population.

A limitation of our trial is that it was open-label, due to the difficulty obtaining appropriate placebos early in the pandemic, and is therefore at risk of bias particularly on patient reported outcomes. However, our choice of hospitalisation and death as the primary outcome is unlikely to be markedly influenced by selection, detection or observer bias. Detection of hospitalisation episodes has benefitted from restricted movement during lockdowns, from use of regional and national electronic health records and by systematic contacting of all participants, with >95% follow-up. Moreover, a placebo effect from perceived benefits of treatment would tend towards a positive effect of azithromycin which was not observed. Knowledge of treatment allocation may also lead to a tendency to more antibiotic prescribing in the placebo group – an effect we observed – which would tend to prejudice against azithromycin as an antibiotic, although concomitant use of macrolides specifically was prohibited by the protocol. A second limitation is that, like other studies^23^, we used a clinical diagnosis for inclusion, rather than requiring PCR confirmation, and PCR data were not available on all participants – particularly at the early stages of the pandemic in the UK where limited testing capacity was directed to patients who needed admission to hospital. Whilst it is likely some participants who did not ultimately have COVID-19 may have been enrolled, this decision reflects the situation in many urgent care settings globally where PCR confirmation is not immediately available, and enhances the generalisability of our findings. Nonetheless, SARS-CoV-2 was detected in 65·6% of those with successful PCR assays, much higher than the 31% PCR-positive rate observed in PRINCIPLE^23^, and study results were similar in the overall ITT group and the predefined PCR-positive subgroup analysis. Two other limitations are lack of data on microbiology and on long term outcomes beyond 28 days.

In the past year over 40 clinical trials of azithromycin in COVID-19 have been registered^1^. Given positive data from *in silico* and *in vitro*^2^ screens and data showing suppression of innate inflammatory cytokines *in vitro*, and clinical trial data in non-SARS-CoV-2 alpha and beta coronaviruses^7^, why might these have not translated into clinical efficacy? Other antiviral molecules have had little clinical effect in COVID-19 compared with immunosuppressive therapies, except in very early disease. In common with influenza, it is likely that antivirals are only efficacious in the early viraemic disease stage and are ineffective in severe disease which is more closely linked to differences in host immune factors. In contrast to influenza A pandemics, the antibacterial effects of azithromycin are unlikely to translate into significant clinical benefit in a disease where secondary bacterial pneumonia is rare^27^. Whilst many studies have shown azithromycin suppression of innate cytokines including IL-1β, IL-6, CXCL-8, TNF and GM-CSF – known to be key mediators of severe disease – some of these data may be confounded by antibacterial effects in the original studies, and it may be that the suppression achieved by azithromycin is simply insufficient to overcome the overwhelming cytokine production triggered by this virus in susceptible individuals.

We found no evidence of harm to individuals, despite the relatively high-dose and long course prescribed, and in particular there were no adverse cardiac events; a concern raised by studies of azithromycin and hydroxychloroquine co-prescribing^28^. In a Danish cohort analysis of 10·6 million prescriptions, azithromycin prescribing has been associated cardiovascular death (rate ratio 2·85) compared with no antibiotics, but this is likely to result from the underlying indication as, when compared with penicillin V, there was no increased risk once adjusted for propensity scores (rate ratio 0·93)^29^. This analysis was performed in a population with a low baseline risk of cardiovascular death, and it should be noted we excluded patients with a prolonged QT interval at baseline electrocardiogram. Nonetheless, there are considerable population risks of unwarranted prescribing of azithromycin, which is a highly valuable antimicrobial and yet has a particularly high propensity for inducing antimicrobial resistance, both to macrolides and to other drug classes including β-lactam antibiotics^30^.

In conclusion, our findings in mild-moderate COVID managed in ambulatory care, taken together with trials in early disease in primary care and from trials severe hospitalised disease, suggest there is no evidence that azithromycin reduces hospitalisation, respiratory failure or death compared with standard care, and should not be used in the treatment of COVID-19.

## Data Availability

The data analysed and presented in this study are available from the corresponding author
on reasonable request, providing the request meets local ethical and research governance
criteria after publication. Patient-level data will be anonymised and study documents will be
redacted to protect the privacy of trial participants. The study protocol is provided in the appendix.

## Contributors

TSCH, VSB, JB, SJD, MJ, JM, NR, DL, IDP, DR contributed to conceptualisation and design of the protocol. SJD performed the power calculation. TSCH, LC, VSB, SM, JC, JM, MJ, DL, PM, RB, TB, GJ, FC, DC, SE, JU, LC, BB, RP, CM, DC, KA contributed to acquisition of study data. JC was chair of the data safety monitoring committee. PL was chair of the trial steering committee. Data were analysed by AW, SJD, RK, AF. AW, RK, SJD, LC have accessed and verified the data. TSCH drafted this submission which was approved by all authors.

## Data sharing

The data analysed and presented in this study are available from the corresponding author on reasonable request, providing the request meets local ethical and research governance criteria after publication. Patient-level data will be anonymised and study documents will be redacted to protect the privacy of trial participants. The study protocol is provided in the appendix.

## Acknowledgements

The authors are grateful to all the participants who volunteered and to the clinical and research teams at all the participating centres. We are grateful to James Chalmers, Chris Rogers, Peter Howarth (Data Safety Monitoring Committee), to Ruth Knight, Anne Francis, Rebecca Brown, Samuel Mills, Elizabeth Hamilton (design, operational and statistical support), Mona Bafadhel (protocol design), Lucy Eldridge, Patrick Julier (software programming), and to Paul Little, Mike Bradburn, Peter McQuitty, Najib Rahman, Dominick Shaw (Trial Steering Committee), and to Lou Chan, Ben Bloom, Daniel Lasserson, Thomas Knight, Richard Procter, Claire Millins, David Connell, Kay Adeboye for site participant recruitment. The views expressed are those of the authors and not necessarily those of the NHS, the NIHR or the Department of Health and Social Care. The funders played no role in the study design. The authors vouch for the integrity and completeness of the data and the fidelity of the trial to the protocol.

This study has been conducted as part of the portfolio of trials in the registered UKCRC Oxford Clinical Trials Research Unit (OCTRU) at the University of Oxford. It has followed their Standard Operating Procedures ensuring compliance with the principles of Good Clinical Practice and the Declaration of Helsinki and any applicable regulatory requirements.

