## Appendix for "A randomised clinical trial of azithromycin versus standard care in ambulatory COVID-19 – the ATOMIC2 trial"

### Supplementary Appendix Content

**Appendix 1: ATOMIC2 Protocol V7.0 04Feb2021**

**Appendix 2: Statistical Analysis Plan V1.0 18Mar2021**

**Appendix 3: Supplementary Results**

### Appendix 1: ATOMIC2 Protocol V7.0 04Feb2021

**Trial Title:** A multi-centre open-label two-arm randomised superiority clinical trial of Azithromycin versus usual care In Ambulatory COVID-19 (ATOMIC2)
**Internal Reference Number / Short title:** ATOMIC2

**IRAS Project ID:** 282892 **REC Ref:** 20/HRA/2105

**EudraCT Number**: 2020-001740-26

**Date and Version No**: 04Feb2021, V7.0

**Chief Investigator:** Dr Timothy SC Hinks, University of Oxford

**Investigators:** Prof Mona Bafadhel, University of Oxford

Dr Maisha Jabeen, University of Oxford / Buckinghamshire Healthcare NHS Trust

[illegible]al Birmingham, Sandwell and [illegible]ust

Prof Ian Pavord, University of Oxford

Prof Duncan Richards, University of Oxford

**Sponsor:** University of Oxford, Clinical Trials and Research Governance, Joint Research Office, Joint Research Office, Boundary Brook House, Headington, Oxford, OX3 7GB

**Funder:** Oxford NIHR Biomedical Research Centre Respiratory Theme and University of Oxford MSD COVID-19 Research Response Fund and Pfizer Inc.

Additional funding applications pending.

**Chief Investigator Signature:**

**Statistician Signature:**

I declare the team have no relevant financial conflicts of interest

**Confidentiality Statement**

This document contains confidential information that must not be disclosed to anyone other than the Sponsor, the Investigator Team, HRA, host organisation, and members of the Research Ethics Committee and Regulatory Authorities unless authorised to do so.

© Copyright: The University of Oxford and Oxford University Hospitals NHS Foundation Trust 2019

© Copyright: The University of Oxford and Oxford University Hospitals NHS Foundation Trust 2019

9.6.1 Severity scale score for peak severity of illness ... 28
9.7 Subsequent assessments ... 29
9.7.1 At study recruitment (optional for all sites and participants) ... 30
9.7.2 Sample handling, processing and analysis ... 30
9.8 GP notification ... 31
9.9 Early Discontinuation/Withdrawal of Participants ... 31
9.10 Definition of End of Trial ... 32
10 SAFETY REPORTING ... 32
10.1 Adverse Event Definitions ... 32
10.2 Reportable Events for ATOMIC2 ... 33
10.3 Assessment of Causality ... 33
10.4 Procedures for Reporting Adverse Events ... 34
10.5 Reporting Procedures for Serious Adverse Events ... 34
10.5.1 Events exempt from reporting as SAEs ... 34
10.5.2 Procedure and timelines for reporting of Serious Adverse Events ... 35
10.6 Expectedness ... 35
10.7 SUSAR Reporting ... 35
10.8 Development Safety Update Reports ... 35
11 TRIAL INTERVENTIONS ... 36
11.1 Investigational Medicinal Product(s) (IMP) Description ... 36
11.1.1 Study intervention ... 36
11.1.2 Dose regimen ... 36
11.1.3 Authorisation and safety ... 36
11.1.4 Contraindications ... 37
11.1.5 Cautions ... 37
11.1.6 Pregnancy ... 38
11.1.7 Blinding of IMPs ... 38
11.1.8 Storage and dispensing of IMP ... 38
11.1.9 Compliance with Trial Treatment ... 39
11.1.10 Concomitant Medication ... 39
11.1.11 Post-trial treatment ... 39
11.2 Other Treatments (non-IMPs) ... 39

© Copyright: The University of Oxford and Oxford University Hospitals NHS Foundation Trust 2019

11.3 Other Interventions ... 39
12 STATISTICS ... 39
12.1 Statistical Analysis Plan (SAP) ... 39
12.2 Description of Statistical Methods ... 40
12.3 Sample Size Determination ... 40
12.3.1 Initial Estimate, Pilot Phase ... 40
12.3.2 Sample size: revised estimate and change in primary outcome for pivotal phase ... 41
12.4 Analysis Populations ... 41
12.5 Stopping Rules ... 41
12.6 The Level of Statistical Significance ... 41
12.7 Procedure for Accounting for Missing, Unused, and Spurious Data. ... 41
12.8 Procedures for Reporting any Deviation(s) from the Original Statistical Plan ... 42
13 DATA MANAGEMENT ... 42
13.1 Source Data ... 42
13.2 Access to Data ... 42
13.3 Data Recording and Record Keeping ... 42
13.4 Collection of data ... 43
14 QUALITY ASSURANCE PROCEDURES ... 43
14.1 Risk assessment ... 43
14.2 Monitoring ... 44
14.3 Quality assurance ... 44
14.4 Trial committees ... 44
14.4.1 Data Safety Monitoring Committee (DSMC) ... 44
14.4.2 Trial Steering Committee (TSC) ... 44
15 PROTOCOL DEVIATIONS ... 45
16 SERIOUS BREACHES ... 45
17 ETHICAL AND REGULATORY CONSIDERATIONS ... 45
17.1 Declaration of Helsinki ... 45
17.2 Guidelines for Good Clinical Practice ... 45
17.3 Approvals ... 46
17.4 Reporting ... 46
17.5 Transparency in Research ... 46

© Copyright: The University of Oxford and Oxford University Hospitals NHS Foundation Trust 2019

© Copyright: The University of Oxford and Oxford University Hospitals NHS Foundation Trust 2019

#### 1 KEY TRIAL CONTACTS

| | |
|---|---|
| **Chief Investigator** | Dr Timothy SC Hinks,<br>NDM, Level 5, John Radcliffe Hospital, Headley Way, OX3 9DU<br>T: +44 1865 220885<br>M: +44 771 2261970 |
| **Sponsor** | University of Oxford<br>Clinical Trials and Research Governance, Joint Research Office, Joint Research Office, Boundary Brook House, Headington, Oxford, OX3 7GB, |
| **Funder(s)** | Funded by the National Institute for Health Research (NIHR) Oxford Biomedical Research Centre (BRC) Respiratory Theme & University of Oxford MSD COVID-19 Research Response Fund |
| **Clinical Trials Unit** | Oxford Clinical Trials Research Unit<br>University of Oxford<br>Botnar Research Centre<br>Windmill Road<br>Oxford<br>OX3 7LF<br>Tel: 01865 223469 |
| **Statisticians** | Susan Dutton and Ariel Wang<br>Oxford Clinical Trials Research Unit, Tel: 01865 223469 |
| **Committees** | **Data safety monitoring committee (DSMC)**<br>Prof James Chalmers (Chair)<br>University of Dundee, Nethergate, Dundee, Scotland, UK, DD1 4HN<br><br>**Trial steering committee (TSC)**<br>Prof Paul Little (Chair)<br>University of Southampton; School of Primary Care, Population Sciences and Medical Education, Aldermoor Health Centre, Aldermoor Close, Southampton, SO16 5ST |

© Copyright: The University of Oxford and Oxford University Hospitals NHS Foundation Trust 2019

#### 2 LAY SUMMARY

Coronavirus-induced disease 2019 (COVID-19) is an infection caused by a virus whose full name is severe acute respiratory syndrome coronavirus 2 (SARS-CoV-2). This is a new and rapidly-spreading infectious disease. Those people that come into contact with the virus can have symptoms such as a mild fatigue, fever, loss of taste and a persistent cough, which can develop into severe respiratory failure requiring hospitalisation and mechanical ventilation. For those where the symptoms worsen this typically occurs 1 to 2 weeks into coming in contact with the virus. This provides a window of opportunity to potentially treat those patients who present with symptoms before becoming seriously ill to take a drug that might not result in them developing the severe symptoms. The ATOMIC2 study is investigating if a common antibiotic called Azithromycin (AZM) may prevent the patients from getting worse. Azithromycin is a safe, inexpensive, antibiotic that is available worldwide and is often prescribed by doctors across the world and it has been proved to have a wide range of antibacterial, anti-inflammatory and antiviral properties.

In this study, researchers want to investigate this medicine in patients who have mild symptoms of COVID-19 who go to the hospital, but who doctors decide there is no need to admit them for treatment. The study will investigate if half the patients are given Azithromycin for 14 days and half the patients do not receive Azithromycin, are there less people after 28 days in one of the groups who go on to develop more severe symptoms from COVID-19.

© Copyright: The University of Oxford and Oxford University Hospitals NHS Foundation Trust 2019

#### 3 SYNOPSIS

| | | | |
|---|---|---|---|
| Trial Title | A multi-centre open-label two-arm randomised superiority clinical trial of Azithromycin versus usual care In Ambulatory COVID-19 (ATOMIC2) | | |
| Internal ref. no. (or short title) | ATOMIC2 | | |
| Trial registration | This trial will be registered with https://clinicaltrials.gov/ | | |
| Sponsor | University of Oxford<br>Clinical Trials and Research Governance, Joint Research Office, Joint Research Office, Boundary Brook House, Headington, Oxford, OX3 7GB, | | |
| Funder | Funded by the National Institute for Health Research (NIHR) Oxford Biomedical Research Centre (BRC) Respiratory Theme and University of Oxford MSD COVID-19 Research Response Fund and Pfizer Inc | | |
| Clinical Phase | Phase II/III | | |
| Trial Design | Multi centre, prospective two-arm open-label randomized superiority clinical trial | | |
| Trial Participants | Adults, ≥18 years of age with clinically-diagnosed COVID-19 infection managed initially as outpatients. | | |
| Sample Size | Initial estimate: Approximately 800 participants (400 per arm). The DSMC will review the accruing data, safety and efficacy data and a futility analysis once the first 100 participants have completed their 28-day follow-up. If progression to the full trial is recommended the endpoints and sample size assumptions will also be reviewed blinded to treatment allocation to refine the final definitive study sample size.<br><br>The interim analysis proposed continuation of the trial with a change in the primary outcome to death or all-cause hospitalisation and recommended a final sample size of at least 291 participants to detect a difference in hospitalisation from 15% to 5% with 80% power. | | |
| Planned Trial Period | Treatment duration 14 days.<br>Duration of follow up: 28 days from randomisation unless admitted to hospital, when participant will be followed up until discharged.<br>Duration of study recruitment:<br>Anticipated – up to 6 months | | |
| Planned Recruitment period | May-December 2020 | | |
| | **Objectives** | **Outcome Measures** | **Timepoint(s)** |
| Primary | To compare the effect of Azithromycin in | Efficacy will be determined through differences in the | Day 28 |

© Copyright: The University of Oxford and Oxford University Hospitals NHS Foundation Trust 2019

| | | | |
|---|---|---|---|
| | participants with a clinical diagnosis of COVID-19 in reducing the proportion with either death or hospital admission from any cause over the 28 days from randomisation. | proportion requiring hospital admission, from any cause, or death over the 28 days from randomisation | |
| Secondary | To compare the effect of Azithromycin in participants with a clinical diagnosis of COVID-19 in reducing the proportion with either death or hospital admission with respiratory failure requiring Non-Invasive Mechanical Ventilation (NIV) or Invasive Mechanical Ventilation (IMV) over the 28 days from randomisation. | Efficacy will be determined through differences in the proportion with either death or admission with respiratory failure requiring level 2 ventilation (NIV/CPAP/ nasal high flow) or level 3 IMV) over the 28 days from randomisation | Day 28 |
| | To compare the effect of Azithromycin in participants with a PCR-confirmed diagnosis of COVID-19 in reducing the proportion with either death or hospital admission with respiratory failure requiring invasive or non-invasive mechanical ventilation over 28 days from randomisation. (For those with SWAB results) | Efficacy will be determined through differences in the proportion with either death or admission with respiratory failure requiring level 2 ventilation (NIV/CPAP/ nasal high-flow) or level 3 (invasive mechanical ventilation) over 28 days from randomisation using a retrospective analysis of oropharyngeal swabs for those patients who had a COVID-19 swab obtained at time of randomisation. | Day 28 |

© Copyright: The University of Oxford and Oxford University Hospitals NHS Foundation Trust 2019

| | | | |
|---|---|---|---|
| | To compare the effect of Azithromycin in participants with a PCR-confirmed diagnosis of COVID-19 in reducing the proportion with either death or all-cause hospital admission over 28 days from randomisation.<br>(For those with SWAB results) | Efficacy will be determined through differences in the proportion with all-cause hospital admission or death over 28 days from randomisation using a retrospective analysis of oropharyngeal swabs for those patients who had a COVID-19 swab obtained at time of randomisation. | Day 28 |
| | To compare differences in all-cause mortality. | Data on vital status | Day 28 |
| | To compare differences in pneumonia progression. | Progression to pneumonia. This will be diagnosed on a CXR or CT thorax with compatible clinical findings, where there was no consolidation on the baseline CXR. | Day 28 |
| | To compare differences in proportion progressing to severe pneumonia. | Retrospective review of CXRs or CT thorax to determine evolution of pneumonia from pneumonia on baseline (CXR). Severe pneumonia is defined as BTS CURB-65 score 3-5. | Day 28 |
| | To compare differences in peak severity of illness. | Maximum severity score during the study period will be recorded. | Ascertain from day 14 and 28 phone call and retrospective medical notes/ePR data at day 28 or death, whichever soonest. |
| | Safety and tolerability | Serious adverse events and concomitant medications. Record at enrolment, emergently during study period and proactively elicit at day 14 and at day 28. | Emergent data collection days 0-28 and elicit proactively at day 14 and day 28 post randomisation. |
| **Exploratory Objectives** | Mechanistic analysis of blood and nasal | The following samples may be taken Blood for serum, Tempus tube (whole blood | Samples to be collected prospectively at baseline and again if |

© Copyright: The University of Oxford and Oxford University Hospitals NHS Foundation Trust 2019

| | | | |
|---|---|---|---|
| | biomarkers if available | RNA), EDTA tubes (PBMC), nasal brush to be placed immediately into RNA lysis buffer (for subsequent PCR and transcriptomic analysis). | patient admitted, to be taken as soon as possible and within 72 hours of admission if possible. |
| Comparator | Standard care according to the hospital protocol where the patient is being triaged and there is a clinical decision not to admit the patient. | | |
| Intervention IMP(s) | Azithromycin 500 mg orally daily for 14 days. | | |

© Copyright: The University of Oxford and Oxford University Hospitals NHS Foundation Trust 2019

### 4 ABBREVIATIONS

| | |
|---|---|
| AE | Adverse event |
| ALT | Alanine aminotransferase |
| APTT | Activated partial thromboplastin time |
| AR | Adverse reaction |
| AST | Aspartate aminotransferase |
| AZM | Azithromycin |
| BP | Blood pressure |
| BTS | British Thoracic Society |
| CCL | Chemokine (C-C motif) ligand |
| CF | Cystic Fibrosis |
| CI | Chief Investigator |
| COPD | Chronic Obstructive Pulmonary Disease |
| CPAP | Continuous positive airway pressure (ventilation) |
| CRA | Clinical Research Associate (Monitor) |
| CRF | Case Report Form |
| CRP | C-reactive protein |
| CT | Computed tomogram |
| CTA | Clinical Trials Authorisation |
| CTRG | Clinical Trials and Research Governance |
| CURB-65 | Confusion, Urea >7.0 mmol/L, Respiratory Rate >/=30 breaths/min, Blood pressure <90 systolic or </=60 diastolic, Age >/= 65 years. |
| CXR | Chest X-Ray |
| DMSC | Data Monitoring Committee / Data Monitoring and Safety Committee |
| DPB | Diffuse Pan Bronchiolitis |
| DSUR | Development Safety Update Report |
| EDTA | Ethylenediaminetetraacetic acid |
| ePR | Electronic patient record |
| FBC | Full blood count |
| GCP | Good Clinical Practice |
| GCSF | Granulocyte Colony Stimulating Factor |
| GP | General Practitioner |

© Copyright: The University of Oxford and Oxford University Hospitals NHS Foundation Trust 2019

| | |
|---|---|
| HFOT | High-Flow Oxygen Therapy: warmed, humidified oxygen delivered via a nasal mask at >15 L/min |
| HR | Heart Rate |
| HRA | Health Research Authority |
| ICF | Informed Consent Form |
| ICH | International Conference on Harmonisation |
| IL | Interleukin |
| IMP | Investigational Medicinal Product |
| IMV | Invasive mechanical ventilation (ventilatory support delivered via an endotracheal tube) |
| ISARIC | International Severe Acute Respiratory and Emerging Infection Consortium |
| ISG | Interferon Stimulated Gene |
| MHRA | Medicines and Healthcare products Regulatory Agency |
| MERS | Middle East Respiratory Syndrome |
| MMRM | Mixed Model for Repeated Measurement |
| MxA | A membrane protein |
| NIV | Non-invasive ventilation (ventilatory support via an external face mask in a non-sedated person) |
| NHS | National Health Service |
| PCR | Polymerase chain reaction |
| PEEP | Positive End-Expiratory Pressure |
| PI | Principal Investigator |
| PIL | Participant/ Patient Information Leaflet |
| PROBE | Prospective randomized open blinded end-point (PROBE) clinical trial |
| PT | Prothrombin time |
| QTc | Corrected QT interval |
| R&D | NHS Trust R&D Department |
| RCT | Randomised Controlled Trial |
| REC | Research Ethics Committee |
| RES | Research Ethics Service |
| RNA | Ribonucleic acid |
| RR | Respiratory Rate |
| RSI | Reference Safety Information |

© Copyright: The University of Oxford and Oxford University Hospitals NHS Foundation Trust 2019

| | |
|---|---|
| RV | RhinoVirus |
| SAE | Serious Adverse Event |
| SAR | Serious Adverse Reaction |
| SARS | Severe Acute Respiratory Syndrome |
| SDV | Source Data Verification |
| SMPC | Summary of Medicinal Product Characteristics |
| SOP | Standard Operating Procedure |
| SSRI | Selective Serotonin Reuptake Inhibitor |
| SST | Serum Separating Tube |
| SUSAR | Suspected Unexpected Serious Adverse Reactions |
| Tempus tube | Blood collection tubes for RNA purification |
| TMF | Trial Master File |
| U+E | Urea and electrolytes |

### 5 BACKGROUND AND RATIONALE

Azithromycin (AZM) is an orally active synthetic macrolide antibiotic with a wide range of antibacterial, anti-inflammatory and antiviral properties. It is a safe, inexpensive, generic licensed drug available worldwide, on the WHO list of essential medications, and manufactured to scale and therefore an ideal candidate molecule to be repurposed as a potential candidate therapy for pandemic COVID-19. Macrolides, particularly Azithromycin, were used to treat 1/3 of severe cases of MERS-CoV[1] and Azithromycin has been tried in COVID-19 infection[2] although RCT data are lacking[3].

#### 5.1 Antiviral properties

Azithromycin has well-documented, broad antiviral properties *in vitro*. Numerous studies have shown it to be effective against respiratory viruses, including the picornavirus human rhinovirus (RV), where it enhances viral-induced type I and type III interferons and interferon-stimulated gene (ISG) expression and reduced RV replication and release[4-6]. Macrolides reduce RV replication *in vitro* by enhancing type I and III IFN and induce the antiviral ISGs viperin and MxA[6] (Figure 1c, Appendix A). *In vivo* in a large, well-designed, RCT of 420 adults with severe asthma long term AZM strikingly reduced exacerbations by 40% over 1 year (Gibson, Lancet 2017)[7] (Figure 1a, Appendix A). These effects occurred irrespective of inflammatory phenotype, and are likely mediated by the antiviral effects, as viruses trigger 80% of exacerbations in asthma[8,9].

© Copyright: The University of Oxford and Oxford University Hospitals NHS Foundation Trust 2019

Macrolides have shown efficacy *in vitro* against a wide range of other viruses. These include the flavivirus Zika, where AZM was a key hit in a drug screen of 2177 compounds and markedly reduce viral proliferation and virus-induced cytopathic effects[10] (Figure 1b, Appendix A). In Zika, AZM upregulates type 1 and type III interferon responses and the viral pathogen recognition receptors MDA5 and RIG-I, and increases the levels of phosphorylated TBK1 and IRF3[11]. There is also evidence of *in vitro* activity against enteroviruses[12], Ebola[13,14] and SARS[15]; with *in vivo* activity against influenza A, with reduction in IL-6, IL-8, IL-17, CXCL9, sTNF and CRP in a small open label RCT[16] (Figure 1d, Appendix A).

##### 5.2 Anti-inflammatory properties

It is likely that AZM's anti-inflammatory properties – rather than antiviral – will be more important in the treatment of severe COVID-19 disease in secondary care. Antivirals are likely to have limited efficacy in severe disease as they are administered late in the disease, after viraemia has peaked[17-19]. In stark contrast to the early cytokine storm responsible for 50% of deaths from influenza A, most COVID-19-related deaths occur due to sudden, late respiratory decompensation, peaking at day 14 after the onset of symptoms[20]. By this time viral loads are low, and it is during the adaptive immunity stage that a late increase of innate / acute phase inflammatory cytokines occurs, including IL-1β, IL-2, IL-6, IL-7, IL-8, GCSF, MCP, MIP1a, TNF[21] and associated with poor outcome[21]. These dysregulated cytokines are associated with features of hemophagocytic lymphohistiocytosis[22] and interstitial mononuclear inflammatory infiltrates, dominated by lymphocytes[23]. This points to a failure not of viral control, but of the ability to halt an over-exuberant inflammatory cascade. Therefore the priority should be to target the off-switch for these signalling cascades, which are characteristically steroid-resistant[19], and associated with pulmonary inflammation and extensive lung damage in SARS patients[24] and MERS-CoV[21,25].

AZM's anti-inflammatory properties include dose-dependent suppression of lymphocyte expression of perforin, and of many of these cytokines, including IL-1⍰, IL-6 and TNF, IL-8(CXCL8), IL-18, G-CSF and GM-CSF[26-29] and other components of the IL-1⍰/IL-6-induced acute phase response such as serum amyloid protein A[27]. For these reasons they have proven clinically efficacy in asthma, COPD, CF and obliterative bronchiolitis, post lung transplant obliterative bronchiolitis and diffuse pan bronchiolitis (DPB): a disease characterised by alveolar accumulation of foamy macrophages[26,30]. In DPB macrolide therapy has dramatically increased survival from 10-20% to 90%[26,31,32], attributed to AZM's inhibition of dysregulated IL-1, IL-2, TNF and GM-CSF[33].

A key cell in the steroid-resistant ARDS which develops in COVID-19 are pro-inflammatory monocyte-derived macrophages[34], which are increased in severe disease, replacing alveolar macrophages[35]. Macrophage-derived cytokines tend to be resistant to corticosteroids. It is also a cell type markedly impaired by diabetes, a dominant risk factor for COVID-19 related death. An important property of macrolides is that they accumulate 100-1000-fold[26,27] in lysosomes of

© Copyright: The University of Oxford and Oxford University Hospitals NHS Foundation Trust 2019

phagocytes and are released in those sites when they die. Within the alveolar macrophage AZM attenuates LPS-induced expression of pro-inflammatory cytokines through inhibition of AP-1[36,37], it inhibits arachidonic acid release in LPS-stimulated macrophages[38], inhibits GM-CSF[27,36,39] and increases phagocytosis, likely by upregulation of CD206, the macrophage mannose receptor[40]. AZM attenuates type 1 response and shifts macrophage polarization to a more immunosuppressive, tissue repair M2-phenotype[41-43]. Thus AZM reduces M1 macrophage markers CCR7, CXCL11, IL-12p70 and enhanced IL-10 and CCL18. This inhibitory profile is similar to that of chloroquine (another potential COVID-19 repurposed drug) due to their similar propensities for lysosomal accumulation.

#### 5.3 Antibacterial properties

Whilst not the main rationale for its use in COVID-19, the broad antibacterial properties of AZM which is active against a range of gram positive, gram negative, anaerobic and atypical infections, may reduce secondary infection which were found in 16% of COVID-19 deaths[20], which could be sufficient grounds for a clinical trial irrespective of other effects, and therefore data on development of pneumonia will be analysed as a secondary outcome.

#### 5.4 Justification for using Azithromycin and dose regimen

Azithromycin has marketing authorisation in the UK and in EU member states. AZM is generally well-tolerated with a very good and well-documented safety record. It is associated with diarrhoea. Whilst there have been concerns about cardiovascular risk, huge epidemiological studies suggest these are very small effects (e.g. 47 extra deaths / million prescriptions) or perhaps no effect when corrected for confounding. It is contraindicated in known hypersensitivity to the drug. It can be used in pregnancy. It should be used in caution in those receiving some other drugs including fluoroquinolones such as moxifloxacin and levofloxacin, and in patients with ongoing proarrhythmic conditions. [For full details see 'Investigational Medicinal Product Description' below and the Summary of Product Characteristics SmPC].

Due to its long half-life AZM accumulates over time, but to achieve a rapid effect we will use 500mg OD for 14 days, similar to the dose recommended for Lyme disease[44]

The trial will use commercial AZM provided by Pfizer. However, in the unlikely event of short supplies, the protocol will allow any authorised brand that contains the active ingredient azithromycin.

#### 5.5 Rationale for design

We will therefore perform an efficacy study of AZM to prevent and/or reduce the severity of lower respiratory tract illness in adult patients with clinically-diagnosed COVID-19 infection being assessed in secondary care but initially managed on an ambulatory care pathway. This provides a

© Copyright: The University of Oxford and Oxford University Hospitals NHS Foundation Trust 2019

therapeutic window of opportunity to avert development of more severe disease. Participants will be randomised to receive Azithromycin 500 mg daily for 14 days or standard care. The first dose will be within 4 hours of randomisation.

##### 5.6 Potential additional study arms

This area of research is rapidly evolving and it may become possible to include additional interventions as extra arms into this study to enable rapid assessment of potentially important treatments in this patient population. If this becomes possible a protocol amendment and amendments to associated documents (PIS, consent forms etc) will be submitted for approval to the sponsor, REC and Competent Authority before any changes are implemented and the comparison for any new intervention will use concurrent usual care controls.

#### 6 OBJECTIVES AND OUTCOME MEASURES

##### 6.1 Hypothesis

Use of Azithromycin 500 mg once daily for 14 days is effective in preventing and/or reducing the severity of lower respiratory illness of COVID-19 disease at 28 days.

| Objectives | Outcome Measures | Time point(s) of evaluation of this outcome measure (if applicable) |
|---|---|---|
| **Primary Objective**<br><br>To compare the effect of Azithromycin in participants with a clinical diagnosis of COVID-19 in reducing the proportion with hospital admission from any cause or death over the 28 days from randomisation. | Efficacy will be determined through differences in the proportion requiring hospital admission from any cause or death, over the 28 days from randomisation | Determined at day 28 from randomisation. |
| **Secondary Objectives** | | |
| To compare the effect of Azithromycin in participants with a clinical diagnosis of COVID-19 in reducing the proportion with | Efficacy will be determined through differences in the proportion with either death or admission with respiratory failure requiring level 2 | Determined at day 28 from randomisation. |

© Copyright: The University of Oxford and Oxford University Hospitals NHS Foundation Trust 2019

| Objectives | Outcome Measures | Time point(s) of evaluation of this outcome measure (if applicable) |
|---|---|---|
| either death or hospital admission with respiratory failure requiring invasive or non-invasive mechanical ventilation over 28 days from randomisation. | ventilation (NIV/CPAP/nasal high-flow) or level 3 (invasive mechanical ventilation) in the 28 days from randomisation. | |
| To compare the effect of Azithromycin in participants with a PCR-confirmed diagnosis of COVID-19 in reducing the proportion with either death or hospital admission with respiratory failure requiring invasive or non-invasive mechanical ventilatory support over 28 days from randomisation (for those who had a COVID-19 swab at randomisation) | Efficacy will be determined through differences in the proportion with either death or admission with respiratory failure requiring level 2 ventilatory support (NIV/CPAP/nasal high-flow) or level 3 (invasive mechanical ventilation) in the 28 days from randomisation using a retrospective analysis of COVID-19 oropharyngeal swabs for those who had one taken at time of randomisation. | Determined at day 28 from randomisation. |
| To compare the effect of Azithromycin in participants with a PCR-confirmed diagnosis of COVID-19 in reducing the proportion with all-cause hospital admission or death (for those who had a COVID-19 swab at randomisation) | Efficacy will be determined through differences in the proportion with all-cause hospital admission or death in the 28 days from randomisation using a retrospective analysis of COVID-19 oropharyngeal swabs for those who had one taken at time of randomisation. | Determined at day 28 from randomisation. |
| To compare differences in all-cause mortality. | Data on vital status (alive / dead, with date and presumed cause of death if appropriate) at 28 days from randomisation | Ascertain data at 28 days after randomisation. |

© Copyright: The University of Oxford and Oxford University Hospitals NHS Foundation Trust 2019

| Objectives | Outcome Measures | Time point(s) of evaluation of this outcome measure (if applicable) |
|---|---|---|
| To compare differences in proportion progressing to pneumonia. | Progression to pneumonia as diagnosed by chest x-ray (or CT thorax), with compatible clinical findings, if no pneumonia is present at time of enrolment. To be diagnosed by a medically qualified doctor and data obtained from review of case-notes and relevant radiology. | Ascertain this information at time of pneumonia diagnosis, or at 28 days after randomisation (whichever is sooner) |
| To compare differences in proportion progressing to severe pneumonia. | Evolution of pneumonia, as diagnosed by chest x-ray or CT thorax, if pneumonia is present at time of enrolment. To be diagnosed by a medically qualified doctor and data obtained from review of case-notes and relevant radiology. Severe pneumonia is defined as BTS CURB-65 score of 3-5. | Ascertain this information retrospectively at 28 days after randomisation |
| To compare differences in peak severity of illness. | The scoring system is described in section 9.6.1 reflects the severity of respiratory illness. The maximum severity score during the entire study period will be compared. | Ascertain from day 14 and day 28 telephone call and from retrospective ePR/medical notes data at 28 days after randomisation. |
| Safety and tolerability | Serious adverse events and concomitant medications. Record at enrolment, emergently during study | Emergent data collection days 0-28 and elicit proactively at day |

© Copyright: The University of Oxford and Oxford University Hospitals NHS Foundation Trust 2019

| Objectives | Outcome Measures | Time point(s) of evaluation of this outcome measure (if applicable) |
| --- | --- | --- |
| | period and proactively elicit at day 14 and at day 28. | 14 and day 28 post randomisation. |
| **Exploratory Objectives** Mechanistic analysis of blood and nasal biomarkers if available | The following samples may be taken. Blood for serum, Tempus tube (whole blood RNA), EDTA tubes (PBMC), nasal brush to be placed immediately into RNA lysis buffer (for subsequent PCR and transcriptomic analysis). | Samples to be collected prospectively at baseline and again if patient admitted, to be taken as soon as possible and within 72 hours of admission if possible. |

### 7 TRIAL DESIGN

**Study design:** Multi centre, prospective open label two-arm randomised superiority clinical trial of standard care and Azithromycin with standard care alone for those presenting to hospital with COVID-19 symptoms who are not admitted at initial presentation.

**Study setting:** Patients being assessed by secondary care NHS hospitals in the UK.

**Participants:** Adults, ≥18 years of age assessed in an acute hospital with clinical diagnosis of COVID-19 infection and where medically it is decided not to admit the patient and for the patient to be managed on an ambulatory (outpatient) care pathway at their usual residence (home or care home).

**Study schedule:** Enrolment on day 0. Telephone follow up at day 14 day, and day 28. If admitted between randomisation and day 28, data will be collected until hospital discharge (Figure 2, Appendix B).

© Copyright: The University of Oxford and Oxford University Hospitals NHS Foundation Trust 2019

**Intervention**: Azithromycin 500 mg orally once daily for 14 days. The first dose will be within 4 hours of randomisation. This is in addition to standard care as per local hospital advice for those patients with suspected COVID who are not admitted: i.e. symptomatic relief with rest, as-required paracetamol (where appropriate) and advice to seek further medical attention if significant worsening of breathlessness.

**Comparator:** Standard care as per local hospital advice for those patients with suspected COVID who are not admitted: i.e. symptomatic relief with rest, as-required paracetamol (where appropriate) and advice to seek further medical attention if significant worsening of breathlessness.

**Data collection:** Study data will be collected as per the study schedule. Serious adverse events and concomitant medications will be monitored from Day 0 until Day 28 via the day 14 and day 28 contacts made to participants, any direct contacts to the central trial team via the study contact details or via patient/notes reviews at sites if participants are admitted. Severity Scale for Clinical Improvement scores (see section 9.6.2) will be collected at days 0, 14 and 28 for patients who remain in an ambulatory pathway and daily for patients who are admitted until hospital discharge and at day 14 and day 28. Data will be collected by face-to-face discussion and assessment of patient (for hospital admissions), or by telephone discussion (for those ambulatory patients), as well as from retrospective patient records/notes and radiology results (for all patients).

© Copyright: The University of Oxford and Oxford University Hospitals NHS Foundation Trust 2019

##### 7.1 Study Schedule:

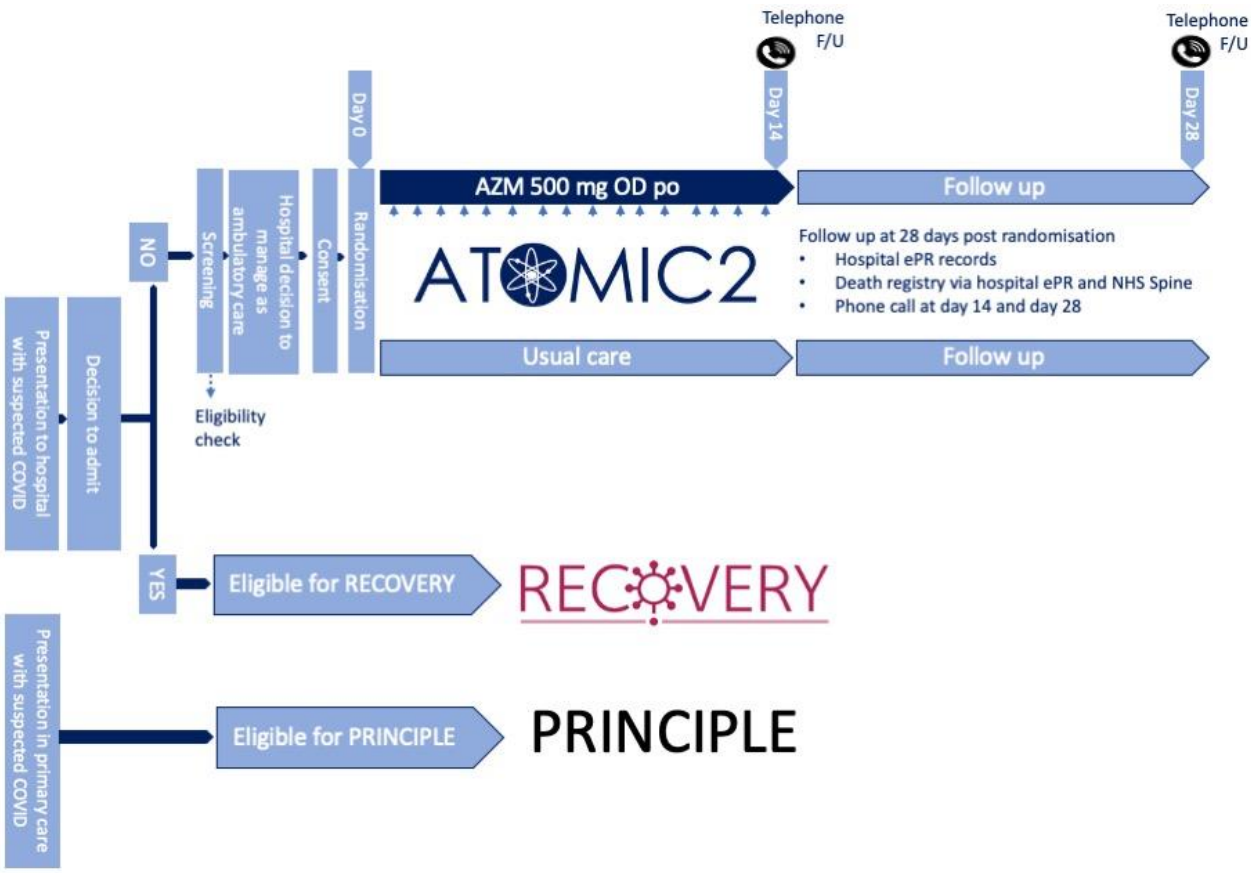

*Note – there is a national trial open in the UK recruiting called RECOVERY this only recruits patients who are hospitalised with COVID-19 – therefore ATOMIC2 is not competing for patients.*

##### 7.2 Sample size: initial estimate, pilot phase

The definitive trial will recruit approximately 800 participants (400 per arm). An interim analysis has been built into this trial after an initial 100 patients have been randomised, treated and followed-up for 28 days. The DSMC will review the accruing safety and efficacy data and the results from a futility analysis, which will assess whether the trial would be likely to confirm superiority of the active treatment if it was to continue as planned. They will provide recommendations to the TSC as to whether the trial should continue to the definitive trial or stop early for safety or futility.

*Note: Whilst the interim analysis is prepared for and undertaken, and the DSMC meeting held – recruitment will continue to the trial as per protocol.*

If the recommendation is to continue to the definitive trial they will also review the assumption on which the sample size is based, blinded to treatment allocation, and the final sample size for

© Copyright: The University of Oxford and Oxford University Hospitals NHS Foundation Trust 2019

the definitive study will be confirmed by the DSMC and TSC. This is a rapidly evolving disease area and information about the control rate for progression to hospitalisation or death is not yet fully known. More details will be available during the trial and will be assessed to refine the sample size. Initial assumptions are described below.

The total sample size of 778 participants with primary outcome data are required to reject the null hypothesis of no difference between the active treatment and usual care. This number is based on the following assumptions: 30-40% of patients following usual care will progress to hospitalisation or death within 28 days; 90% power, 2-sided 5% significance and a 30-35% reduction in progression or death for patients on Azithromycin (778 participants required to detect a 33.3% reduction from 30% to 20% in progression to hospitalisation or death or 646 required to detect a 30% reduction from 40% to 28% in progression to hospitalisation or death). To allow for uncertainty around the assumptions and allowing for a 2% loss to follow-up, approximately 800 participants will be required. Total sample size for the definitive study will be refined at the interim analysis if progression to the full trial is the option chosen.

##### 7.3 Sample size: revised estimate and change in primary outcome for pivotal phase

Data from the first 109 participants reaching the 28 day primary outcome time-point was reviewed by the DSMC. At this time no participants had been admitted to hospital requiring level 2 or 3 ventilatory support (the primary outcome), although some participants had been admitted to hospital. Rates of all-cause hospitalisation of this population were 15% over 28 days. Loss to follow up was <5%. The DSMC recommended to the TSC and TMG that the primary outcome should be reviewed in order for the trial to continue and enable it to answer the research question and that any updated primary outcome should not be subjective as the trial is not blinded.

Following further assessment of the blinded data the Research question has been updated from:

> 'To compare the effect of Azithromycin in participants with a clinical diagnosis of COVID-19 in reducing the proportion with either death or hospital admission with respiratory failure requiring Non-Invasive Mechanical Ventilation (NIV) or Invasive Mechanical Ventilation (IMV) over the 28 days from randomisation.

To:

> 'To compare the effect of Azithromycin in participants with a clinical diagnosis of COVID-19 in reducing the proportion with either death or hospital admission from any cause over the 28 days from randomisation.'

Note that the updated primary outcome, death or all-cause hospitalisation, includes the original primary endpoint, death or hospitalisation requiring level 2 or 3 ventilation, and the latter will still be reported as a secondary outcome at the end of the trial.

© Copyright: The University of Oxford and Oxford University Hospitals NHS Foundation Trust 2019

This change is consistent with the recommendations of the World Health Organisation Blueprint for Covid-19 Therapeutic Trials[44] that the primary endpoint should be responsive to the eligible patient population and the definition of the endpoint should be fine-tuned for the Pivotal Phase, based on the Pilot Phase of the Trial.

Following this change to the primary outcome and based on blinded data from the pilot phase of the study the sample size has been updated:

Assuming a rate of all cause hospitalisation/death of 15% in the usual care arm, then a minimum of 276 participants providing primary end-point data, will provide 80% power and 5% (2-sided) significance to detect a difference from 15% to 5% in the Azithromycin arm, a relative reduction of 66%. Allowing for 5% loss to follow-up, this number is increased to a minimum of 291 participants. If additional participants are recruited (this could potentially occur with the current increase in prevalence of COVID-19 and the rapid recruitment of participants) this will provide more power to estimate the treatment effects with the potential to detect a smaller difference if one exists.

##### 7.4 Study duration

Treatment duration 14 days.
Duration of follow up 28 days from randomisation, unless admitted to hospital within 28 days of randomisation, then the participant will be followed until their discharge from hospital.
Duration of study: Anticipated recruitment period up to 6 months

#### 8 PARTICIPANT IDENTIFICATION

##### 8.1 Trial Participants

Adults, ≥18 years of age assessed in an acute hospital with clinical diagnosis of COVID-19 infection.

##### 8.2 Inclusion Criteria

Patients are eligible for the study if all of the following are true:

- Male or Female, aged at least 18 years
- Assessed by the attending clinical team as appropriate for initial ambulatory (outpatient) management
- A clinical diagnosis of highly-probable or confirmed COVID-19 infection (diagnosis by the attending clinical team) with onset of first symptoms within the last 14 days
- No medical history that might, in the opinion of the attending clinician, put the patient at significant risk if he/she were to participate in the trial
- Able to understand written English (for the information and consent process) and be able to give informed consent

© Copyright: The University of Oxford and Oxford University Hospitals NHS Foundation Trust 2019

##### 8.3 Exclusion Criteria

The participant may not enter the trial if ANY of the following apply:

- Known hypersensitivity to any Macrolide including Azithromycin, Ketolide antibiotic, or the excipients including an allergy to soya or peanuts.
- Known fructose intolerance, glucose-galactose malabsorption or sucrose-isomaltase-insufficiency
- Currently on a Macrolide antibiotic (Clarithromycin, Azithromycin, Erythromycin, Telithromycin, Spiramycin)
- Elevated cardiac troponin at initial assessment suggestive of significant myocarditis (if clinically the clinical team have felt it appropriate to check the patient's troponin levels)
- Evidence of QTc prolongation: QTc>480ms
- Significant electrolyte disturbance (e.g. hypokalaemia $K^+ < 3.5$ mmol/L)
- Clinically relevant bradycardia (P<50 bpm), non-sustained ventricular tachycardia or unstable severe cardiac insufficiency
- Currently on hydroxychloroquine or chloroquine

#### 9 TRIAL PROCEDURES

For schedule of procedures see Appendix B.

##### 9.1 Recruitment

Patients will be identified by the clinical care teams as clinically diagnosed highly-probable or confirmed COVID-19 in acute ambulatory care units, acute medical units and emergency departments within acute hospitals. Initial onset of symptoms consistent with COVID-19 must be within the last 14 days. A member of clinical care team will then screen the patient for eligibility and discuss the trial with the patient, or alternatively, they will ask the patients if a member of the research team could approach them to talk to them about ATOMIC2.

##### 9.2 Screening and Eligibility Assessment

Any patient randomised must be able to receive the first dose of Azithromycin within 4 hours of randomisation. Protocol waivers are not permitted.

Patients will be screened by review of ePR/medical notes data for history of presentation, examination findings, ECG result (to be performed on all participants), $K^+$ and, concomitant medications by anyone listed on the site study delegation log.

© Copyright: The University of Oxford and Oxford University Hospitals NHS Foundation Trust 2019

##### 9.3 Informed Consent

Informed consent will be obtained from each patient before enrolment into the study, by a member of the study team who is listed on the delegation log for taking informed consent.

Written and verbal versions of the Participant Information and Informed Consent will be presented to the participants detailing no less than: the exact nature of the trial; what it will involve for the participant; the implications and constraints of the protocol; the known side effects and any risks involved in taking part. It will be clearly stated that the participant is free to withdraw from the trial at any time for any reason without prejudice to future care, without affecting their legal rights and with no obligation to give the reason for withdrawal.

The participant will be allowed time to consider the information, and the opportunity to question the Investigator, however due to the nature of the disease and limited time available, they may not be time for the individual to contact their GP or other independent parties to decide whether they will participate in the trial, as individuals can only enter the study whilst being reviewed in the COVID-19 areas of hospital, individuals cannot go home and then decide to take part in the study.

Written Informed Consent will then be obtained by means of participant dated signature and dated signature of the person who presented and obtained the Informed Consent. The person who obtained the consent must be suitably qualified and experienced, and have been authorised to do so by the Chief/Principal Investigator. Consent will be taken electronically, and a copy of the patient's consent form will be emailed to the study site and the patient from a secure NHS email address. Consent will be requested after presentation of the trial Patient Information Sheet and a discussion has been had with the patient.

##### 9.4 Randomisation

A medically qualified doctor must confirm that a patient is eligible for this CTIMP. Eligible patients will be randomised using the centralised validated computer randomisation program through a secure (encrypted) web-based service, RRAMP (https://rramp.octru.ox.ac.uk), provided by the Oxford Clinical Trials Research Unit (OCTRU), accessed via the study's RedCap instance, with a minimisation algorithm to ensure balanced allocation across treatment groups, stratified by centre, hypertension (yes/no), diabetes (yes/no) and sex (male/female) in a 1:1 ratio to either Azithromycin or usual care. To ensure the unpredictability of treatment allocation the minimisation algorithm will include a probabilistic element and a small number of participants randomised by simple randomisation.

*Note: Hypertension is defined as any hypertension previously diagnosed by a doctor prior to presenting to the hospital with COVID-19 symptoms.*
*Note: Diabetes is defined as any diabetes that is treated with oral or injectable therapy.*

© Copyright: The University of Oxford and Oxford University Hospitals NHS Foundation Trust 2019

Stratification by centre will help to ensure that any centre-effect will be equally distributed in the trial arms and enable practical issues associated with the active intervention to be overcome.

There is some emerging evidence that patients who have underlying hypertension, diabetes and are male are more likely to progress and require hospitalisation, so it is important for the two treatments to be balanced across these potentially important prognostic factors.

The following information will be recorded on a secure web-based form in the study randomisation system (RRAMP) by the attending clinician or delegate including a member of research team to enable follow-up:

- Patient details e.g. name, NHS number, date of birth, sex, telephone number and GP details

*Note: These data fields will allow sites to check their local hospital records and/or NHS Spine (or devolved nation equivalents) to check that a patient has not died or been admitted to avoid any upset of patient's relatives. The GP details are required to allow the central trial team to send a letter to the patient's GP informing them of their ATOMIC2 participation.*

- *An email address will also be recorded to enable a copy of the completed consent form to be sent to the patient or at their request a different individual for safe keeping.*

#### 9.5 Blinding and code-breaking

This is not applicable, as this is an open label study without blinding.

#### 9.6 Baseline Assessments (Day 0)

The following information will be recorded on the web-based form which goes straight into the password protected study database by the attending clinician or delegate including a member of research team:

- COVID-19 symptom onset date
- Presence or absence of COVID -19 symptoms using COVID COS scales (0=no, 1=mild, 2=moderate, 3=significant)
  - Shortness of Breath
  - Fever (Temperature ≥ 37.8°C, oral/rectal or tympanic)
  - Loss of taste
  - New persistent cough
  - Diarrhoea
  - Body pain
  - Changes to sense of smell
  - Fatigue
- COVID-19 symptoms history
- Latest vital signs (HR, RR, Pulse Oximetry, BP, Temperature)
- Major comorbidity and medical history
  - cardiovascular disease
  - diabetes,
  - chronic lung disease
  - asthma
  - hypertension
  - ongoing cancer treatment

© Copyright: The University of Oxford and Oxford University Hospitals NHS Foundation Trust 2019

- Date of initial assessment in an acute hospital
- Smoking status
- Ethnicity
- Occupation (including specifically whether a HealthCare or Laboratory worker)
- Charlson Index
- Severity score (as per section 9.6.1)
- Concomitant medications (specifically prednisolone, inhaled corticosteroids, ACE inhibitors, antibiotics)
- Results of chest auscultation
- CXR results i.e presence or absence of pneumonia
- Usual residence (home or residential care)
- Number of members in household
- Number of members including participants currently showing any COVID-19 symptoms
- If an oropharyngeal viral swab for COVID-19 was taken
- Usual care biochemistry results from hospital (U+E, FBC, Fibrinogen, D-Dimer, APTT, PT, Troponin)
- 12 lead ECG
- If the participant is known to be pregnant
- If the participant is lactating

##### 9.6.1 Severity scale score for peak severity of illness

The Severity scale score should be given based on clinical condition. If a patient is hospitalised for reasons of isolation or quarantine they should not automatically be given the score of 3, ascribe the score which is relevant to their clinical status, which maybe for example ambulatory score 1 even if they are hospitalised. The highest score obtained during the 28 day study period will be used in the final analysis, based on data obtained at 14 and 28 days.

| Descriptor | Score |
|---|---|
| Ambulatory. No limitation of activities | 0 |
| Limitation of simple activities | 1 |
| Hospitalised, mild disease, no oxygen therapy | 2 |
| Hospitalised, oxygen by conventional delivery system[1] ≤40% mask or nasal prongs | 3 |
| Hospitalised, oxygen by conventional delivery system[1] >40% mask | 4 |
| Hospitalised receiving non-invasive ventilation or receiving high-flow oxygen therapy (HFOT, >15 L/min), or continuous positive airway pressure (CPAP)[2] | 5 |
| Intubation and mechanical ventilation[3] | 6 |
| Ventilation + additional organ support | 7 |
| Death | 8 |

[1] Criteria filled if oxygen is required to maintain saturations >92% or above normal baseline for patients who use home oxygen [2] Any patient requiring any form of PEEP delivery, or those in whom such devices are not tolerated requiring >50% oxygen with a RR>25, or rising $CO_2$ in the absence of known lung disease

© Copyright: The University of Oxford and Oxford University Hospitals NHS Foundation Trust 2019

[3] In patients unsuitable for ventilation, criterion is met when requiring >50% oxygen with a RR>25, or rising $CO_2$.

#### 9.7 Subsequent assessments

Subsequent assessments will be carried out at days 14 and 28 by telephone call.

The following information will be recorded:

- For those randomised to AZM - date of starting treatment
- Presence of COVID-19 symptoms using COVID COS scales 0=no, 1=mild, 2=moderate, 3=significant)
    - Shortness of Breath
    - Fever (Temperature ≥ 37.8°C, oral/rectal or tympanic)
    - Loss of taste
    - New persistent cough
    - Diarrhoea
    - Body pain
    - Changes to sense of smell
    - Fatigue
- (At 14 day call only) For those randomised to AZM – number of tablets remaining
- COVID-19 symptoms history for past 14 days
- COVID-19 swab results from hospital records if a swab was taken
- Any change to concomitant medications
- Worse severity scale score in previous 14 days
- Any visits to Hospital due to COVID-19 symptoms in previous 14 days
- Any adverse events

There will be a mortality check at day 14 and day 28 **before any contact is made with participants**, using hospital systems and NHS Spine or equivalent devolved nation systems – it is anticipated that because the UK Government COVID-19 lockdown regulations prohibit travel and in the vast majority of cases patients will be readmitted to their local hospital after deterioration. If on calling the participant at day 14 or day 28 the participant has been readmitted, data will be collected by hospital note review instead of from the participant. The following data will be extracted:

- Date of admission
- Name of hospital
- Any level 2 ventilation received – duration, type, date initiated
    - Reason for stopping level 2 ventilation – Need for level 3 IMV, patient improvement, died
- Any level 3 ventilation received – duration, type, date initiated
    - Reason for stopping level 3 ventilation – Died, patient improvement, requirement by others for equipment
- Pneumonia – diagnosis, severity (using BTS CURB-65 score).

© Copyright: The University of Oxford and Oxford University Hospitals NHS Foundation Trust 2019

- Daily severity scale score
- Date of discharge from hospital, location of discharge
- Any adverse events
- Concomitant medications (specifically prednisolone, inhaled corticosteroids, ACE inhibitors, antibiotics, antivirals and antifungals)
- Any complications from COVID-19 occurring during admission
- Any other COVID-19 trials participated in

If participating sites allow, have staff available with suitable experience, equipment and time, then the following samples are optional to be given by participants, for the study's exploratory outcomes.

###### 9.7.1 At study recruitment (optional for all sites and participants)

- EDTA tube to be frozen for subsequent DNA analysis. (4ml blood)
- Serum SST tube to be frozen (5ml blood)
- Tempus blood RNA tube to be frozen (3ml blood)
- Nasal sampling using a small brush into lysis buffer to be frozen
- EDTA tubes for cell preparation of peripheral blood mononuclear cells to be frozen (1 x 10ml EDTA tubes, 1 x 4ml EDTA tube)

**On hospital admission if it occurs within 28 days of randomisation the following samples will be taken (optional for all sites and participants)**

- Serum SST tube to be frozen (5ml blood)
- Tempus blood RNA tube to be frozen (3ml blood)
- Nasal sampling using a small brush into lysis buffer to be frozen
- EDTA tubes for cell preparation of peripheral blood mononuclear cells to be frozen (1 x 10ml EDTA tubes, 1 x 4ml EDTA tube)

###### 9.7.2 Sample handling, processing and analysis

A sample handling manual will be provided separately. The trial team will have access to the samples. Samples may be used for analysis of immunological, virological and genetic parameters. If whole genome sequencing is performed to determine predisposition to severe disease, incidental findings will be of unknown significance to participants and will not be made available to them unless they are directly related to a relevant immune defect or of relevance to COVID-19. De-identified samples might be analysed by commercial organisations.

De-identified samples, where consent is in place, may be used for future ethically approved studies.

*Note: If a COVID-19 swab is taken at site that is not tested locally – these swabs will be stored in Category 3 freezers at site, then transferred to the University of Oxford at the end of the study in*

© Copyright: The University of Oxford and Oxford University Hospitals NHS Foundation Trust 2019

*a controlled manner. If the COVID-19 swab is processed at the hospital, the results of this test will be extracted from the patients' medical notes/ePR.*

##### 9.8 GP notification

Participants will be asked for permission for their GP to contacted to notify them of their participation in the study. As this study is open label, GPs will be informed of whether the participant has been dispensed a 14 day course of Azithromycin or no drug. No further communications will be made with the GP as any hospital admission and treatment will result in standard hospital discharge paperwork being automatically sent to the GP.

##### 9.9 Early Discontinuation/Withdrawal of Participants

During the course of the trial a participant may choose to withdraw early from the trial treatment at any time. This may happen for a number of reasons, including but not limited to:

- Inability to comply with trial procedures
- Participant decision

Participants may choose to stop treatment but may remain on study follow-up.

Each participant has the right to withdraw their consent to continue from the study at any time. The reason for withdrawal will be asked and this will be recorded in the CRF, but the participant is not obliged to give a reason. In the event of a discontinuation from the trial during the 14 days following randomisation, participants will be asked to stop taking the IMP. Participants may withdraw from active follow-up and further communication but allow the trial team to continue to access their medical records and any relevant hospital data that is recorded as part of routine standard of care; i.e., CT-Scans, blood results and disease progression data etc. Participants may request that samples they have donated be destroyed.

In addition, the Investigator may discontinue a participant from the trial treatment at any time if the Investigator considers it necessary for any reason including, but not limited to:

- Development of new significant hepatic dysfunction (See Cautions, section 10.1.5)
- Development of severe cardiac dysfunction.
- Ineligibility (either arising during the trial or retrospectively having been overlooked at screening)
- Significant non-compliance with treatment regimen or trial requirements

If the participant is withdrawn due to a serious adverse event, the Investigator will arrange for follow-up until the adverse event has resolved or stabilised.

© Copyright: The University of Oxford and Oxford University Hospitals NHS Foundation Trust 2019

##### 9.10 Definition of End of Trial

The end of trial will be when all samples taken have been analysed and all the data has been entered into the clinical database and all queries have been resolved.

#### 10 SAFETY REPORTING

##### 10.1 Adverse Event Definitions

| | |
|---|---|
| Adverse Event (AE) | Any untoward medical occurrence in a participant to whom a medicinal product has been administered, including occurrences which are not necessarily caused by or related to that product. |
| Adverse Reaction (AR) | An untoward and unintended response in a participant to an investigational medicinal product which is related to any dose administered to that participant.<br><br>The phrase "response to an investigational medicinal product" means that a causal relationship between a trial medication and an AE is at least a reasonable possibility, i.e. the relationship cannot be ruled out.<br><br>All cases judged by either the reporting medically qualified professional or the Sponsor as having a reasonable suspected causal relationship to the trial medication qualify as adverse reactions. |
| Serious Adverse Event (SAE) | A serious adverse event is any untoward medical occurrence that:<br>• results in death<br>• is life-threatening<br>• requires inpatient hospitalisation or prolongation of existing hospitalisation<br>• results in persistent or significant disability/incapacity<br>• consists of a congenital anomaly or birth defect*.<br><br>Other ‘important medical events’ may also be considered a serious adverse event when, based upon appropriate medical judgement, the event may jeopardise the participant and may require medical or surgical intervention to prevent one of the outcomes listed above. |

© Copyright: The University of Oxford and Oxford University Hospitals NHS Foundation Trust 2019

| | |
|---|---|
| | NOTE: The term "life-threatening" in the definition of "serious" refers to an event in which the participant was at risk of death at the time of the event; it does not refer to an event which hypothetically might have caused death if it were more severe. |
| Serious Adverse Reaction (SAR) | An adverse event that is both serious and, in the opinion of the reporting Investigator, believed with reasonable probability to be due to one of the trial treatments, based on the information provided. |
| Suspected Unexpected Serious Adverse Reaction (SUSAR) | A serious adverse reaction, the nature and severity of which is not consistent with information in the MHRA approved Reference Safety Information |

*Note: To avoid confusion or misunderstanding of the difference between the terms "serious" and "severe", the following note of clarification is provided:*

*"Severe" is often used to describe intensity of a specific event, which may be of relatively minor medical significance.*

*"Seriousness" is the regulatory definition supplied above.*

##### 10.2 Reportable Events for ATOMIC2

As discussed in section 10.5.1 reportable events are those events which are serious and related to AZM treatment . In addition, for those patients randomised to AZM all serious cardiovascular events irrespective of causality must also be reported as SAEs. .

SAEs must be recorded in the participant's medical notes and reported to the CTU as described below.

##### 10.3 Assessment of Causality

The relationship of each reportable adverse event to the trial medication must be determined by a medically qualified individual according to the following definitions:

**Unrelated** – Where an event is not considered to be related to the IMP / intervention

**Possibly Related** – although a relationship to the IMP / intervention cannot be completely ruled out, the nature of the event, the underlying disease, concomitant medication or temporal relationship make other explanations possible.

© Copyright: The University of Oxford and Oxford University Hospitals NHS Foundation Trust 2019

**Probably Related** – the temporal relationship and absence of a more likely explanation suggest the event could be related to the IMP / intervention

**Definitely Related** – the known effects of the IMP, its therapeutic class or based on challenge testing suggests that the IMP / intervention is the most likely cause.

##### 10.4 Procedures for Reporting Adverse Events

Azithromycin is a very well known, commonly used drug with a well-known safety profile. The aim of this trial is to test the effectiveness of the drug in preventing deterioration, not explicitly assessing safety of this well used drug. AEs considered related to the trial medication as judged by a medically qualified investigator or the Sponsor will be followed either until resolution, or the event is considered stable. The following information will be recorded: description, date of onset and end date, severity, assessment of relatedness to trial medication and action taken. Follow-up information should be provided as necessary. The severity of events will be assessed on the following scale: 1 = mild, 2 = moderate, 3 = severe.

It will be left to the Investigator's clinical judgment to decide whether or not an AE is of sufficient severity to require the participant's removal from treatment. A participant may also voluntarily withdraw from treatment due to what he or she perceives as an intolerable AE. If either of these occurs, the participant will be followed-up and be given appropriate care under medical supervision until symptoms cease, or the condition becomes stable.

##### 10.5 Reporting Procedures for Serious Adverse Events

All reportable SAEs occurring within the 14 days of the IMP administration and up to 28 days after randomisation will be reported. All SAE information must be recorded on an ATOMIC2-Trial specific SAE form as described below. OCTRU will perform an initial check of the report, request any additional information and forward to a Medical Reviewer (Nominated Person as per OCTRU SOP) for review.

Additional and further requested information (follow-up or corrections to the original case) will be detailed on a new SAE Report Form and emailed to OCTRU.

###### 10.5.1 Events exempt from reporting as SAEs

It is important to consider the natural history of COVID-19 with regards to this study, the expected sequelae of the illness, and the relevance of these complications to the trial treatment. All eligible participants have a potential poor prognosis, and due to the complexity of their condition are at increased risk of experiencing multiple adverse events. Additionally, Azithromycin has a very well known safety profile. Therefore taking a risk adapted approach the labelling of a reportable Serious Adverse Event (SAE) will be limited to serious events which might reasonably occur as a

© Copyright: The University of Oxford and Oxford University Hospitals NHS Foundation Trust 2019

consequence of the trial treatment. In addition any serious cardiovascular event in patients randomised to AZM will be a reportable SAE irrespective of causality.

Events that are part of the natural history of COVID-19 such as hospitalisation and deaths are exempt from reporting as SAEs as they will be captured as part of the primary outcome. An AE should not be recorded for the positive SAR-CoV-19 infection, this will be known at time of inclusion into the study and should be recorded as medical history. Worsening of COVID-19 symptoms is captured as an efficacy measure and in general will not be considered an adverse event.

##### 10.5.2 Procedure and timelines for reporting of Serious Adverse Events

- SAEs must be reported immediately i.e. within 24 hours of site study team becoming aware of the event.
- Site study team will complete an ATOMIC specific SAE report form. The SAE form can either be accessed within the study CRF system or be completed using the paper-based form.
- Site study team will provide additional, missing or follow up information in a timely fashion.

#### 10.6 Expectedness

Assessment of Expectedness will be determined centrally by the Nominated Person and according to the current MHRA approved RSI section of the Summary of Product Characteristics.

#### 10.7 SUSAR Reporting

All SUSARs will be reported by the sponsor delegate to the relevant Competent Authority and to the REC and other parties as applicable. For fatal and life-threatening SUSARS, this will be done no later than 7 calendar days after the Sponsor or delegate is first aware of the reaction. Any additional relevant information will be reported within 8 calendar days of the initial report. All other SUSARs will be reported within 15 calendar days.

Principal Investigators will be informed of any SUSARs with this IMP at the same time as reporting to the MHRA and REC.

#### 10.8 Development Safety Update Reports

The CI will submit (in addition to the expedited reporting above) DSURs once a year throughout the clinical trial, or on request, to the Competent Authority (MHRA in the UK), Ethics Committee, HRA (where required) and Sponsor.

© Copyright: The University of Oxford and Oxford University Hospitals NHS Foundation Trust 2019

### 11 TRIAL INTERVENTIONS

#### 11.1 Investigational Medicinal Product(s) (IMP) Description

##### 11.1.1 Study intervention

Subjects will be randomised to receive Azithromycin 500 mg daily orally for 14 days or standard care. The first dose will be taken within 4 hours of randomisation. Participants will be asked to take the AZM at the approximately the same time every day for 14 days. The drug should be taken ideally 1 hour before a meal or 2 hours afterwards.

The comparator will be usual care. i.e. symptomatic relief with rest, as-required paracetamol (where appropriate) and advice to seek further medical attention if significant worsening breathlessness. No specific therapies are yet available for COVID-19. Should additional interventions become evidence-based standard practice during the conduct of this study these would also be permitted to be provided and will be recorded in the concomitant medications.

##### 11.1.2 Dose regimen

Due to its long half-life AZM accumulates over time, but to achieve a rapid effect we will use 500mg OD for 14 days, similar to the dose recommended for Lyme disease[45].

##### 11.1.3 Authorisation and safety

Azithromycin has a marketing authorisation in the UK and in EU member states. Azithromycin is generally well-tolerated with a very good and well-documented safety record. Even in long term administration (500mg thrice weekly for 48 weeks, n=213 individuals, median age 61y) there was no increase in serious adverse events v placebo, the main adverse event being an increase in diarrhoea (34% v 19% not associated with study withdrawal)[7]. The main adverse event of concern would be potential cardiovascular toxicity. Although macrolides have a class warning for potential cardiac QT prolongation, Azithromycin does not show this effect under experimental conditions[46]. Only a few cases of QT prolongation have been reported for patients treated with the drug[47], mainly because Azithromycin, unlike other macrolide antibiotics, does not interact with CYP3A4, despite a minor interaction with the anti-coagulant warfarin[48]. In the large AMAZES RCT there was no increase in QTc prolongation, although this study excluded participants with QTc>480ms[7]. Recently a large study of Medicaid prescriptions reported an additional risk of cardiovascular death of 47 extra deaths / million v amoxicillin (relative risk (RR) for cardiovascular death 2.49[49], and a meta-analysis of 20 million patients suggested a RR for cardiac death or VT of 2.42[50]. However these effects are very small and subject to confounding, and at odds with more recent studies: in a review of 185,000 Medicare patients odds ratio for CV death was only 1.35, and after controlling for covariates decreased to 1.01 (0.95-1.08)[51], whilst a large Cochrane review of 183 trials found no evidence of an increase in cardiac disorders with macrolides (OR 0.87)[52]. Overall

© Copyright: The University of Oxford and Oxford University Hospitals NHS Foundation Trust 2019

the risk to a patient treated would be low compared with the considerable mortality of COVID-19, particularly if patients with QTc>480ms were excluded.

##### 11.1.4 Contraindications

Hypersensitivity to the active substance, any macrolide, ketolide antibiotic, or the excipients. Known fructose intolerance, glucose-galactose malabsorption or sucrose-isomaltase-insufficiency.

##### 11.1.5 Cautions

Use with caution in patients with ongoing proarrhythmic conditions (Prolonged QT, coadministration with quinidine, procainamide, dofetilide, amiodarone, sotalol, cisapride, terfenadine, antipsychotic agents such as pimozide; antidepressants such as citalopram; and fluoroquinolones such as moxifloxacin and levofloxacin).

Caution with electrolyte disturbance, particularly in cases of hypokalaemia; with clinically relevant bradycardia, non-sustained ventricular tachycardia or unstable severe cardiac insufficiency.

Caution with hepatic dysfunction. Hepatic dysfunction is common in COVID-19 disease, with elevation of alanine aminotransferase (ALT) or aspartate aminotransferase (AST) occurring at some stage during disease in 14–53% of hospitalised cases[53]. However this is typically mild and clinically significant liver injury is uncommon even if severe patients[54]. There is no evidence later presentation is associated with worse hepatic derangement[54]. Mild hepatic dysfunction is not a contraindication to AZM prescription. The SmPC by Pfizer states: A dose adjustment is not necessary for patients with mild to moderately impaired liver function. Since liver is the principal route of elimination for Azithromycin, the use of Azithromycin should be undertaken with caution in patients with significant hepatic disease. Cases of fulminant hepatitis potentially leading to life-threatening liver failure have been reported with Azithromycin. Some patients may have had pre-existing hepatic disease or may have been taking other hepatotoxic medicinal products. In case of signs and symptoms of liver dysfunction, such as rapid developing asthenia associated with jaundice, dark urine, bleeding tendency or hepatic encephalopathy, liver function tests / investigations should be performed immediately[55]. Azithromycin administration should be stopped if significant liver dysfunction (e.g. AST and/or ALT >5x upper limit of normal[56]) has emerged since commencing the drug.

Coumarin-type oral anticoagulants: The SmPC states in a pharmacokinetic interaction study, azithromycin did not alter the anticoagulant effect of a single dose of 15 mg warfarin administered to healthy volunteers. There have been reports received in the post-marketing period of potentiated anticoagulation subsequent to co-administration of azithromycin and coumarin-type oral anticoagulants. Although a causal relationship has not been established, consideration should be given to the frequency of monitoring prothrombin time when azithromycin is used in patients receiving coumarin-type oral anticoagulants. It should also be noted that many potential

© Copyright: The University of Oxford and Oxford University Hospitals NHS Foundation Trust 2019

participants may be prescribed other antibiotics as part of standard care. Therefore concomitant use of coumarins is not contraindicated, but it is recommended in the PIS that if prescribed azithromycin or any other antibiotic participants should attend their GP surgery for a repeat INR check after 3 to 5 days to check.

Nelfinavir: The SmPC states: Co-administration of azithromycin (1200 mg) and nelfinavir at steady state (750 mg three times daily) resulted in increased azithromycin concentrations. No clinically significant adverse effects were observed and no dose adjustment was required. Concomitant use of nelfinavir is therefore not contraindicated.

Trimethoprim/sulfamethoxazole: Co-administration of trimethoprim/sulfamethoxazole DS (160 mg/800 mg) for 7 days with 1200mg azithromycin on Day 7 had no significant effect on peak concentrations, total exposure or urinary excretion of either trimethoprim or sulfamethoxazole. Azithromycin serum concentrations were similar to those seen in other studies. Therefore concomitant use of co-trimoxazole is not contraindicated.

##### 11.1.6 Pregnancy

The SmPC states only to be used during pregnancy if the benefit outweighs the risk. Given the high (30-40%) mortality of this condition in hospitalised patients, and lack of teratogenic effect of azithromycin in animal studies, large human database studies including >1000 live births[57], or post-marketing surveillance[58], pregnancy will not be a contraindication to the study medication, but this decision will be subject to the principal investigator's discretion. Lactation: avoid breastfeeding till 2 days after discontinuation of treatment.[55]

Pregnancy itself is not an AE unless there is a suspicion that the study medication may have interfered with the effectiveness of a contraceptive medication.

##### 11.1.7 Blinding of IMPs

This is an open-label study. However, while the study is in progress, access to tabular results by allocated treatment allocation will not be available to the research team, patients, or members of the Steering Committee (unless the DSMC advises otherwise).

##### 11.1.8 Storage and dispensing of IMP

Commercial stock of study medication will be delivered to participating hospital pharmacies. Depending on supplies, the drug is either provided in blister packs or 100 capsules/bottle.

Hospital pharmacies will work under Exemption 37 to assemble the drug for individual use and will add the approved ATOMIC2 clinical trial label. A copy of the standard drug patient information leaflet normally supplied with Azithromycin will be also be provided.

Once Pharmacy prepares the labelled drug packs these will be sent for secure storage to the acute medicine areas where patients are being triaged. The prescribing doctor will complete the details

© Copyright: The University of Oxford and Oxford University Hospitals NHS Foundation Trust 2019

of the patient and of the dispense date on the label. The research team will also complete drug accountability logs.

###### 11.1.9 Compliance with Trial Treatment

Compliance will be assessed by telephone discussion with patient on day 14 of treatment with specific questioning as to the number of pills remaining. Adequate compliance will be defined as the first dose being administered within 4 hours of randomisation and at least 80% of doses i.e. a maximum of 4 / 28 tablets remaining at the end of day 14.

###### 11.1.10 Concomitant Medication

Cautions and precautions summarised in the SmPC will be followed during this trial. Co-administration of the following medications will not be allowed and will be criteria for exclusion or withdrawal from the trial treatment: quinidine, procainamide, dofetilide, amiodarone, sotalol, cisapride, terfenadine, antipsychotic agents such as pimozide; antidepressants such as citalopram; fluoroquinolones such as moxifloxacin, levofloxacin and ciprofloxacin; chloroquine and hydroxychloroquine; use of another macrolide antibiotic (clarithromycin, erythromycin, azithromycin, telithromycin, spiramycin).

###### 11.1.11 Post-trial treatment

There will be no provision of the IMP beyond the trial period.

##### 11.2 Other Treatments (non-IMPs)

None.

##### 11.3 Other Interventions

There are no other interventions in the trial design.

To assess safety and tolerability: records of adverse events and concomitant medications, reported emergently to the study team and directly elicited on days 14 and 28 at telephone follow-up. In the event of an adverse event the Investigator will arrange for follow-up visits or telephone calls until the adverse event has resolved or stabilised.

#### 12 STATISTICS

##### 12.1 Statistical Analysis Plan (SAP)

Full details of the statistical analysis will be detailed in a separate statistical analysis plan (SAP) which will be drafted early in the trial and finalised prior to the interim analysis data lock. Stata

© Copyright: The University of Oxford and Oxford University Hospitals NHS Foundation Trust 2019

(StataCorp LP) or other appropriate validated statistical software will be used for analysis. A summary of the planned statistical analysis is included here.

#### 12.2 Description of Statistical Methods

Standard descriptive statistics will be used to describe the demographics between the treatment groups reporting means and standard deviations or medians and interquartile ranges as appropriate for continuous variables and numbers and percentages for binary and categorical variables.

The proportion of patients progressing to hospitalisation or death by day 28 post-randomisation is the primary outcome for this study. The difference in proportions between the treatment arms will be assessed using a chi-squared tests and a 5% (2-sided) significance level. Difference in proportions together with the 95% confidence intervals will be reported. Adjusted analyses will also be undertaken using logistic regression with progression as the binary outcome, adjusting for stratification factors (centre, hypertension, diabetes and sex) and other important prognostic variables, which will be fully defined in the SAP. Time to event analysis will also be undertaken to explore whether the active treatment delays progression. The success (or otherwise) of the trial will be based on the adjusted analysis. Both relative and absolute differences in proportions will be reported together with 95% confidence intervals.

Other binary outcomes will be assessed using similar methods and continuous variables will be assessed using linear regression analysis. Where appropriate longitudinal methods will incorporate different time points.

The number and percentage of subjects with each score on the severity scale for clinical improvement will be presented at baseline and each post baseline time point. In addition, the change in severity scale score from baseline will be summarised on both a categorical scale, using counts and percentages and on a continuous scale using descriptive statistics. Inferential statistical analyses such as ordered logistic regression, mixed model for repeated measurement (MMRM) or Mann-Whitney may be conducted in an exploratory fashion to aid the understanding of the data. Binary interpretations of the severity scale for clinical improvement may also be defined in the SAP, such as responders (any improvement at day 14) and complete responders (score of 0 at day 14) and these will be compared using logistic regression as for the primary outcome.

#### 12.3 Sample Size Determination

##### 12.3.1 Initial Estimate, Pilot Phase

See section 7.2.

© Copyright: The University of Oxford and Oxford University Hospitals NHS Foundation Trust 2019

###### 12.3.2 Sample size: revised estimate and change in primary outcome for pivotal phase

See section 7.3.

##### 12.4 Analysis Populations

The intention-to-treat (ITT) population is defined as all randomised patients analysed according to their randomised allocation.

A supplementary ITT population (ITT +ve) is defined as all randomised patients with a positive COVID PCR result.

All efficacy and safety analyses will be based on the ITT population and repeated on the ITT +ve population.

##### 12.5 Stopping Rules

A Data and Safety Monitoring Committee will be set up to review recruitment, trial conduct, safety and efficacy data.

The first formal interim analysis will take place after 100 patients have completed the trial (i.e. approximately 28 days after the 100$^{th}$ patient has been randomised). The DSMC will review the safety of participants and will review the results of the futility analysis (i.e. no evidence of sufficient clinical efficacy to reasonably justify continuing the trial) in order to make recommendations about continuation or otherwise to the full trial. This will be based on Bayesian predictive probabilities, the full details of which will be described in the SAP. If the decision is to continue, then a review of the endpoints and the assumptions taken for the sample size will be undertaken, blinded to treatment allocation, and recommendations provided as to the final sample size for the full trial. Further interim analyses for futility using the same method will be undertaken at the discretion of the DSMC.

##### 12.6 The Level of Statistical Significance

All tests will be completed at a 5% 2-sided significance level. All comparative outcomes will be presented as summary statistics with 95% confidence intervals and reported in accordance with the CONSORT Statement (http://www.consort-statement.org).

##### 12.7 Procedure for Accounting for Missing, Unused, and Spurious Data.

Missing data will be minimised by careful data management. Missing data will be described with reasons given where available; the number and percentage of individuals in the missing category will be presented by treatment arm. All data collected on data collection forms will be used, since only essential data items will be collected. No data will be considered spurious in the analysis since all data will be checked and cleaned before analysis.

© Copyright: The University of Oxford and Oxford University Hospitals NHS Foundation Trust 2019

The nature and mechanism for missing variables and outcomes will be investigated, and if appropriate multiple imputation will be used. In this situation sensitivity analyses will be undertaken assessing the underlying missing data assumptions. Any imputation techniques will be fully described in the Statistical Analysis Plan.

##### 12.8 Procedures for Reporting any Deviation(s) from the Original Statistical Plan

A detailed statistical analysis plan will be drawn up early in the trial with review and appropriate sign-off following OCTRU SOPs. Any changes to the statistical analysis plan during the trial will be subject to the same review and sign-off procedure with details of changes being included in the new version. Any changes/deviations from the original SAP will be described and justified in protocol and/or in the final report, as appropriate.

#### 13 DATA MANAGEMENT

The plans for the data management of the study are outlined below.

##### 13.1 Source Data

Source documents are where data are first recorded, and from which participants' CRF data are obtained. These include, but are not limited to, hospital records (from which medical history and previous and concurrent medication may be summarised into the CRF), clinical and office charts, laboratory and pharmacy records, and radiographs. The study will have a data management plan. All documents will be stored safely in confidential conditions.

##### 13.2 Access to Data

Direct access will be granted to authorised representatives from the Sponsor, host institution and the regulatory authorities to permit trial-related monitoring, audits and inspections.

##### 13.3 Data Recording and Record Keeping

The Investigators will maintain appropriate medical and research records for this trial, in compliance with the principles of GCP and regulatory and institutional requirements for the protection of confidentiality of volunteers. The Chief Investigator, site teams and central study team will have access to records. The Investigators will permit authorised representatives of the Sponsor, as well as ethical and regulatory agencies to examine (and when required by applicable law, to copy) clinical records for the purposes of quality assurance reviews, audits and evaluation of the study safety and progress.

All study data will be captured directly in the study's instance of RedCap or the study instance of their randomisation system RRAMP. There are no paper CRFs, worksheets, questionnaires that will be completed as part of this study.

© Copyright: The University of Oxford and Oxford University Hospitals NHS Foundation Trust 2019

Identifiable information will be recorded on a secure web-based form in the study randomisation system (RRAMP) by the attending clinician or delegate including a member of research team to enable follow-up:

- Patient details e.g. name, NHS (or CHI for Scottish patients) number, date of birth, sex, telephone number and GP details

*Note: These data fields will allow sites to check their local hospital records and NHS Spine or other devolved nation systems to check when to contact a patient that they have not deceased or been admitted to avoid any upset of patient's relatives. The GP details are required to allow the central trial co-ordinating team to ensure a letter is sent out to the patient's GP informing them of their participation in the ATOMIC2 study.*

- *An email address will also be recorded to enable a copy of the completed consent form to be sent to the patient or at their request a different individual for safe keeping.*

The Investigator and/or Sponsor must retain copies of the essential documents for a minimum of 5 years following the end of the study. Site investigators will always have contemporaneous access to all data entered into the system for patients from their site.

The Investigator will inform the Sponsor of the storage location of the essential documents and of any changes in the storage location should they occur. The Investigator must contact the Sponsor for approval before disposing of any documentation. The Investigator should take measures to prevent accidental or premature destruction of these documents.

##### 13.4 Collection of data

Data will be collected by a member of the clinical or study team. Data will also be collected from ePR/medical notes and NHS Spine and by telephone call.

After day 28 of the study, sites will be asked to conduct a notes review to check for any hospital admissions.

#### 14 QUALITY ASSURANCE PROCEDURES

##### 14.1 Risk assessment

A risk assessment will be conducted according to OCTRU's process and a monitoring plan will be drafted to include all central monitoring activities. The trial will be conducted in accordance with

© Copyright: The University of Oxford and Oxford University Hospitals NHS Foundation Trust 2019

the current approved protocol, Principles of GCP, relevant regulations and OCTRU standard operating procedures.

#### 14.2 Monitoring

Due to the nature of this study and timelines and in accordance with the risk assessment in section 14.1 monitoring will be limited to central monitoring activities – there will be no site monitoring, missing data will be queried with sites where mandatory. Monitoring of the data will occur as the data is being entered into the database.

#### 14.3 Quality assurance

The Sponsor or its designated representative will assess each study site to verify the qualifications of each Investigator and the site staff and to ensure that the site has all of the required equipment. A virtual study Initiation meeting will occur where among other things the Investigator will be informed of their responsibilities and procedures for ensuring adequate and correct study documentation.

#### 14.4 Trial committees

##### 14.4.1 Data Safety Monitoring Committee (DSMC)

The DSMC is a group of independent experts external to the trial who assess the progress, conduct, participant safety and critical endpoints of the study. The study DSMC will adopt a DAMOCLES charter which defines its terms of reference and operation in relation to the oversight of the trial. They will review the interim analysis after 100 participants have completed the trial and make recommendations to the TSC as to the continuation or otherwise of the trial. They will also review the endpoints and sample size assumptions to finalise the sample size for the full definitive trial.

This Group will provide advice and recommendations to the TSC and may correspond directly with the Sponsor if potential safety concerns are raised.

##### 14.4.2 Trial Steering Committee (TSC)

The TSC include independent members and members of the research team and provides overall supervision of the trial on behalf of the funder. Its terms of reference will be agreed and will be recorded in a TSC charter.

© Copyright: The University of Oxford and Oxford University Hospitals NHS Foundation Trust 2019

#### 15 PROTOCOL DEVIATIONS

A trial related deviation is a departure from the ethically and regulatory approved trial protocol or other trial document or process (e.g. consent process or IMP administration) or from Good Clinical Practice (GCP) or any applicable regulatory requirements. Any deviations from the protocol will be documented in a protocol deviation form, discussed as per the deviation SOP and filed in the trial master file.

#### 16 SERIOUS BREACHES

The Medicines for Human Use (Clinical Trials) Regulations contain a requirement for the notification of "serious breaches" to the MHRA within 7 days of the Sponsor becoming aware of the breach.

A serious breach is defined as "A breach of GCP or the trial protocol which is likely to affect to a significant degree –

(a) the safety or physical or mental integrity of the subjects of the trial; or

(b) the scientific value of the trial".

In the event that a serious breach is suspected the Sponsor must be contacted within 1 working day. In collaboration with the CI the serious breach will be reviewed by the Sponsor and, if appropriate, the Sponsor will report it to the REC committee, Regulatory authority and the relevant NHS host organisation within seven calendar days.

#### 17 ETHICAL AND REGULATORY CONSIDERATIONS

##### 17.1 Declaration of Helsinki

The Investigator will ensure that this trial is conducted in accordance with the principles of the Declaration of Helsinki.

##### 17.2 Guidelines for Good Clinical Practice

The Investigator will ensure that this trial is conducted in accordance with the principles of Good Clinical Practice.

© Copyright: The University of Oxford and Oxford University Hospitals NHS Foundation Trust 2019

##### 17.3 Approvals

The protocol, informed consent form, participant information sheet and any proposed advertising material will be submitted to an appropriate Research Ethics Committee (REC), HRA, regulatory authorities (MHRA in the UK), and host institution(s) for written approval.

The Investigator will submit and, where necessary, obtain approval from the above parties for all substantial amendments to the original approved documents.

##### 17.4 Reporting

The CI shall submit once a year throughout the clinical trial, or on request, an Annual Progress Report to the REC, HRA, host organisation, funder (where required) and Sponsor, and a DSUR to the MHRA. In addition, an End of Trial notification and final report will be submitted to the MHRA, the REC, host organisation and Sponsor.

##### 17.5 Transparency in Research

Prior to the recruitment of the first participant, the trial will have been registered on a publicly accessible database.

Results will be uploaded to the European Clinical Trial (EudraCT) Database within 12 months of the end of trial declaration by the CI or their delegate.

Where the trial has been registered on multiple public platforms, the trial information will be kept up to date during the trial, and the CI or their delegate will upload results to all those public registries within 12 months of the end of the trial declaration.

##### 17.6 Participant Confidentiality

The study will comply with the General Data Protection Regulation (GDPR) and Data Protection Act 2018, which require data to be de-identified as soon as it is practical to do so. The processing of the personal data of participants will be minimised by making use of a unique participant study number only on all study documents and any electronic database(s), with the exception of the storage of patient and their GP contact details to enable follow-up of the participants. This data is stored in an encrypted form. All documents will be stored securely and only accessible by study staff and authorised personnel. The study staff will safeguard the privacy of participants' personal data.

Instances of missing, discrepant, or uninterpretable data will be queried with the Investigator for resolution. Any changes to study data will be documented in an audit trail, which will be maintained within the clinical database.

© Copyright: The University of Oxford and Oxford University Hospitals NHS Foundation Trust 2019

In compliance with the principles of ICH GCP and regulatory requirements, the Sponsor, a third party on behalf of the Sponsor, regulatory agencies or Independent Ethics Committees (IEC) may conduct quality assurance audits at any time during or following a study. In the event of monitoring, the Investigator must agree to allow monitoring of the study according to ICH GCP requirements and The Medicines for Human Use (Clinical Trials) Regulations 2004 (including all modifications [Statutory Instruments] made since 2004).

The Investigator should also agree to allow auditors direct access to all study-related documents including source documents. They must also agree to allocate their time and the time of their study staff to the auditors in order to discuss findings and issues.

##### 17.7 CTU Involvement

This study will be coordinated by the UKCRC registered Oxford Clinical Trials Research Unit (OCTRU) at the University of Oxford.

#### 18 FINANCE AND INSURANCE

##### 18.1 Funding

This study is supported by the Oxford Respiratory NIHR Biomedical Research Centre and University of Oxford MSD COVID-19 Research Response Fund, and through funding of a BRC clinical fellow and through an NIHR Senior Research Fellowship to the CI. The CI salary is funded by the Wellcome Trust. The trial drug is provided at no cost by Pfizer Inc; but if it is no longer possible to source the drug from Pfizer any brand can be used. Pfizer Inc has also provided a grant to the study team to enable this study.

##### 18.2 Insurance

The University has a specialist insurance policy in place which would operate in the event of any participant suffering harm as a result of their involvement in the research (Newline Underwriting Management Ltd, at Lloyd's of London). NHS indemnity operates in respect of the clinical treatment that is provided.

##### 18.3 Contractual arrangements

Appropriate contractual arrangements will be put in place with all third parties.

© Copyright: The University of Oxford and Oxford University Hospitals NHS Foundation Trust 2019

#### 19 PUBLICATION POLICY

Publications will acknowledge the funders with the following text: The research was funded by the Wellcome Trust (104553/z/14/z, 211050/Z/18/z),the National Institute for Health Research (NIHR) Oxford Biomedical Research Centre (BRC), the University of Oxford and Pfizer Inc. The study drug is initially being provided free-of-charge by Pfizer Inc. who had no part in the study design, conduct or analysis.

The following disclaimer after the acknowledgement must be added: “The views expressed are those of the author(s) and not necessarily those of the NHS, the NIHR or the Department of Health, the University of Oxford or Pfizer Inc.”

#### 20 DEVELOPMENT OF A NEW PRODUCT/ PROCESS OR THE GENERATION OF INTELLECTUAL PROPERTY

Not applicable.

#### 21 ARCHIVING

Both paper and electronic trial data will be retained through an archiving service as per the sponsoring institute’s policy, and data will be retained for a minimum of 5 years after termination of the trial.

© Copyright: The University of Oxford and Oxford University Hospitals NHS Foundation Trust 2019

© Copyright: The University of Oxford and Oxford University Hospitals NHS Foundation Trust 2019

18. Baden LR, Rubin EJ. Covid-19 - The Search for Effective Therapy. N Engl J Med 2020.
19. Martinez MA. Compounds with therapeutic potential against novel respiratory 2019 coronavirus. Antimicrob Agents Chemother 2020.
20. Ruan Q, Yang K, Wang W, Jiang L, Song J. Clinical predictors of mortality due to COVID-19 based on an analysis of data of 150 patients from Wuhan, China. Intensive care medicine 2020.
21. Huang C, Wang Y, Li X, et al. Clinical features of patients infected with 2019 novel coronavirus in Wuhan, China. Lancet 2020;395:497-506.
22. Shakoory B, Carcillo JA, Chatham WW, et al. Interleukin-1 Receptor Blockade Is Associated With Reduced Mortality in Sepsis Patients With Features of Macrophage Activation Syndrome: Reanalysis of a Prior Phase III Trial. Crit Care Med 2016;44:275-81.
23. Xu Z, Shi L, Wang Y, et al. Pathological findings of COVID-19 associated with acute respiratory distress syndrome. Lancet Respir Med 2020.
24. Wong CK, Lam CW, Wu AK, et al. Plasma inflammatory cytokines and chemokines in severe acute respiratory syndrome. Clin Exp Immunol 2004;136:95-103.
25. Mahallawi WH, Khabour OF, Zhang Q, Makhdoum HM, Suliman BA. MERS-CoV infection in humans is associated with a pro-inflammatory Th1 and Th17 cytokine profile. Cytokine 2018;104:8-13.
26. Altenburg J, de Graaff CS, van der Werf TS, Boersma WG. Immunomodulatory effects of macrolide antibiotics - part 1: biological mechanisms. Respiration 2011;81:67-74.
27. Parnham MJ, Erakovic Haber V, Giamarellos-Bourboulis EJ, Perletti G, Verleden GM, Vos R. Azithromycin: mechanisms of action and their relevance for clinical applications. Pharmacology & therapeutics 2014;143:225-45.
28. Marjanovic N, Bosnar M, Michielin F, et al. Macrolide antibiotics broadly and distinctively inhibit cytokine and chemokine production by COPD sputum cells in vitro. Pharmacol Res 2011;63:389-97.
29. Shinkai M, Foster GH, Rubin BK. Macrolide antibiotics modulate ERK phosphorylation and IL-8 and GM-CSF production by human bronchial epithelial cells. Am J Physiol Lung Cell Mol Physiol 2006;290:L75-85.
30. Hui D, Yan F, Chen RH. The effects of azithromycin on patients with diffuse panbronchiolitis: a retrospective study of 29 cases. J Thorac Dis 2013;5:613-7.
31. Kudoh S, Azuma A, Yamamoto M, Izumi T, Ando M. Improvement of survival in patients with diffuse panbronchiolitis treated with low-dose erythromycin. Am J Respir Crit Care Med 1998;157:1829-32.
32. Nagai H, Shishido H, Yoneda R, Yamaguchi E, Tamura A, Kurashima A. Long-term low-dose administration of erythromycin to patients with diffuse panbronchiolitis. Respiration 1991;58:145-9.
33. Weng D, Wu Q, Chen XQ, et al. Azithromycin treats diffuse panbronchiolitis by targeting T cells via inhibition of mTOR pathway. Biomed Pharmacother 2019;110:440-8.
34. Zhou Y, Fu B, Zheng X, et al. Aberrant pathogenic GM-CSF+ T cells and inflammatory CD14+CD16+ monocytes in severe pulmonary syndrome patients of a new coronavirus. bioRxiv 2020.
35. Liao M, Liu Y, Yuan J, et al. The landscape of lung bronchoalveolar immune cells in COVID-19 revealed by
single-cell RNA sequencing bioRxiv 2020.

© Copyright: The University of Oxford and Oxford University Hospitals NHS Foundation Trust 2019

36. Bosnar M, Cuzic S, Bosnjak B, et al. Azithromycin inhibits macrophage interleukin-1beta production through inhibition of activator protein-1 in lipopolysaccharide-induced murine pulmonary neutrophilia. Int Immunopharmacol 2011;11:424-34.
37. Meyer M, Huaux F, Gavilanes X, et al. Azithromycin reduces exaggerated cytokine production by M1 alveolar macrophages in cystic fibrosis. Am J Respir Cell Mol Biol 2009;41:590-602.
38. Banjanac M, Munic Kos V, Nujic K, et al. Anti-inflammatory mechanism of action of azithromycin in LPS-stimulated J774A.1 cells. Pharmacol Res 2012;66:357-62.
39. Khan AA, Slifer TR, Araujo FG, Remington JS. Effect of clarithromycin and azithromycin on production of cytokines by human monocytes. Int J Antimicrob Agents 1999;11:121-32.
40. Hodge S, Hodge G, Jersmann H, et al. Azithromycin improves macrophage phagocytic function and expression of mannose receptor in chronic obstructive pulmonary disease. Am J Respir Crit Care Med 2008;178:139-48.
41. Murphy BS, Sundareshan V, Cory TJ, Hayes D, Jr., Anstead MI, Feola DJ. Azithromycin alters macrophage phenotype. The Journal of antimicrobial chemotherapy 2008;61:554-60.
42. Yamauchi K, Shibata Y, Kimura T, et al. Azithromycin suppresses interleukin-12p40 expression in lipopolysaccharide and interferon-gamma stimulated macrophages. Int J Biol Sci 2009;5:667-78.
43. Legssyer R, Huaux F, Lebacq J, et al. Azithromycin reduces spontaneous and induced inflammation in DeltaF508 cystic fibrosis mice. Respir Res 2006;7:134.
44. Organisation WH. WHO R&D Blueprint novel Coronavirus COVID-19 Therapeutic Trial Synopsis. Geneva: World Health Organisation; 2020 18/02/2020.
45. Press BGP. The British National Formulary. London, UK2020.
46. Milberg P, Eckardt L, Bruns HJ, et al. Divergent proarrhythmic potential of macrolide antibiotics despite similar QT prolongation: fast phase 3 repolarization prevents early afterdepolarizations and torsade de pointes. J Pharmacol Exp Ther 2002;303:218-25.
47. Kezerashvili A, Khattak H, Barsky A, Nazari R, Fisher JD. Azithromycin as a cause of QT-interval prolongation and torsade de pointes in the absence of other known precipitating factors. J Interv Card Electrophysiol 2007;18:243-6.
48. Kanoh S, Rubin BK. Mechanisms of action and clinical application of macrolides as immunomodulatory medications. Clin Microbiol Rev 2010;23:590-615.
49. Ray WA, Murray KT, Hall K, Arbogast PG, Stein CM. Azithromycin and the risk of cardiovascular death. N Engl J Med 2012;366:1881-90.
50. Cheng YJ, Nie XY, Chen XM, et al. The Role of Macrolide Antibiotics in Increasing Cardiovascular Risk. J Am Coll Cardiol 2015;66:2173-84.
51. Polgreen LA, Riedle BN, Cavanaugh JE, et al. Estimated Cardiac Risk Associated With Macrolides and Fluoroquinolones Decreases Substantially When Adjusting for Patient Characteristics and Comorbidities. J Am Heart Assoc 2018;7.
52. Hansen MP, Scott AM, McCullough A, et al. Adverse events in people taking macrolide antibiotics versus placebo for any indication. Cochrane Database Syst Rev 2019;1:CD011825.
53. Zhang C, Shi L, Wang FS. Liver injury in COVID-19: management and challenges. Lancet Gastroenterol Hepatol 2020.
54. Bangash MN, Patel J, Parekh D. COVID-19 and the liver: little cause for concern. Lancet Gastroenterol Hepatol 2020.
55. Limited S. Summary of Product Characteristics 2019 25/09/2019

© Copyright: The University of Oxford and Oxford University Hospitals NHS Foundation Trust 2019

56. Severity Grading In Drug Induced Liver Injury. LiverTox: Clinical and Research Information on Drug-Induced Liver Injury. Bethesda (MD)2012.
57. Bahat Dinur A, Koren G, Matok I, et al. Fetal safety of macrolides. Antimicrob Agents Chemother 2013;57:3307-11.
58. LAB-0606-6.0. 2019. (Accessed 30/04/2020, 2020, at http://labeling.pfizer.com/ShowLabeling.aspx?id=650#Data1.)

© Copyright: The University of Oxford and Oxford University Hospitals NHS Foundation Trust 2019

#### 23 APPENDIX A: Figure 1 – Background data for trial rationale

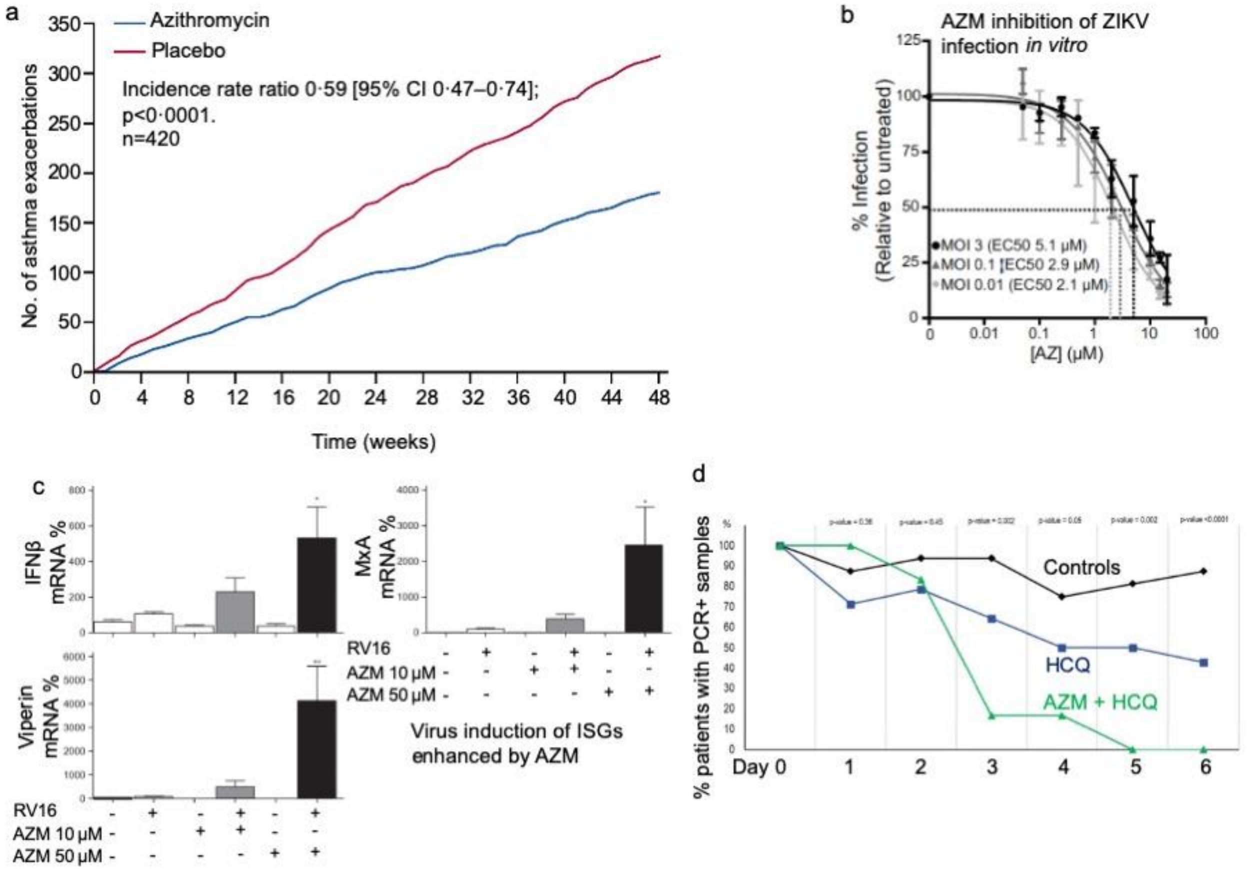

Figure 1 (a) Cumulative severe and moderate asthma exacerbations during 48 weeks of treatment with Azithromycin 500 mg, three times per week, or placebo in AMAZES[7] (b) Azithromycin inhibits ZIKV infection in U87 glial cells. $EC_{50}$ values for AZM-mediated reduction of ZIKV infection were 5.1 µM, 2.9 µM, 2.1 µM for an MOI of 3, 0.1, 0.01 respectively[10]. (c) AZM enhanced rhinovirus-induced interferon-stimulated genes in human bronchial epithelial cells[4]. (d) PCR + status in small, non-randomised clinical trial suggesting earlier return to PCR-ve status with combined hydroxychloroquine (HCQ) and Azithromycin (AZM)[2].

© Copyright: The University of Oxford and Oxford University Hospitals NHS Foundation Trust 2019

#### 24 APPENDIX B: SCHEDULE OF PROCEDURES

| | Study day | | | |
|---|---|---|---|---|
| **Procedures** | Day 0 (Day of randomisation) | 14 days after randomisation (Study Day 14) – Participant contacted by phone | 28 days after randomisation (Study Day 28) – Participant contacted by phone | Any hospital admission |
| Consent | ✓ | | | |
| Eligibility check | ✓ | | | |
| Demographics | ✓ | | | |
| Medical history | ✓ | | | |
| ECG | ✓ | | | |
| Medication history | ✓ | ✓ | ✓ | ✓ |
| Swab taken for COVID PCR test (if possible) | ✓ | | | |
| Randomisation | ✓ | | | |
| Dispensing of 14 day course of IMP (if randomised to IMP) | ✓ | | | |
| Medical notes / ePR / biochemistry results/ microbiology results review | ✓ | | | ✓ |
| Radiology review (if any performed on clinical grounds) | ✓ | | | ✓ |
| Assessment of outcome measures (vital status, history of admission)(ePR/ notes / Death register / Telephone call) | | ✓ | ✓ | ✓ |
| Compliance assessment (telephone call) | | ✓ | ✓ | |
| Study Blood sampling (optional) | ✓ (serum sample + Tempus, EDTA) | | | ✓ |
| Nasal brush (optional, for observational) | ✓ | | | |
| SAE/AE reporting | ✓ | ✓ | ✓ | ✓ |

© Copyright: The University of Oxford and Oxford University Hospitals NHS Foundation Trust 2019

#### 25 APPENDIX C: EXAMPLE OF PARTICIPANT'S STUDY JOURNEY

| Study Day | Day of the Week | Study procedures |
|---|---|---|
| 0 | Monday | Recruitment<br>Consent<br>Randomisation<br>First dose of IMP (if allocated) |
| 1 | Tuesday | Second dose of IMP (if allocated) |
| 2 | Wednesday | Third dose of IMP (if allocated) |
| 3 | Thursday | Fourth dose of IMP (if allocated) |
| 4 | Friday | Fifth dose of IMP (if allocated) |
| 5 | Saturday | Sixth dose of IMP (if allocated) |
| 6 | Sunday | Seventh dose of IMP (if allocated) |
| 7 | Monday | Eighth dose of IMP (if allocated) |
| 8 | Tuesday | Ninth dose of IMP (if allocated) |
| 9 | Wednesday | Tenth dose of IMP (if allocated) |
| 10 | Thursday | Eleventh dose of IMP (if allocated) |
| 11 | Friday | Twelfth dose of IMP (if allocated) |
| 12 | Saturday | Thirteenth dose of IMP (if allocated) |
| 13 | Sunday | Fourteenth dose of IMP (if allocated) (THIS IS THE LAST DOSE) |
| 14 | Monday | Check at site that it is suitable to contact the participant<br><br>Day 14 follow-up of patient by telephone |
| 28 | Monday | Check at site that it is suitable to contact the participant<br><br>Day 28 follow-up of patient by telephone |

The above assumes that a study participant does not get admitted to hospital nor dies within 28 days of randomisation.

If a participant gets admitted to hospital within 28 days of randomisation – then they are followed up whilst in hospital until they are discharged – this may go past 28 days since randomisation.

If a participant dies before 28 days of randomisation – this will be recorded at recruiting sites using data either held in hospital records or on NHS Spine and there may not be any data collection at day 14 or day 28.

© Copyright: The University of Oxford and Oxford University Hospitals NHS Foundation Trust 2019

### 26 APPENDIX D: AMENDMENT HISTORY

| Protocol Version No. | Date issued | Author(s) of changes | Details of Changes made | Justification for change |
|---|---|---|---|---|
| 7.0 | 04Feb2021 | Timothy Hinks and Susan Dutton (as advised by TSC) | 1. Change in primary outcome to all-cause hospitalisation<br>2. Revised power calculation in light<br>3. Addition of new secondary endpoint<br>4. Typographical corrections. | 1. The pre-planned interim analysis after enrolment of the first 100 patients showed no patients had developed a primary outcome (death or hospital admission with respiratory failure requiring mechanical ventilation). Therefore the DSMC and TSC recommended, after blinded review of data, that the outcome be changed to all cause hospitalisation, consistent with the WHO Covid-19 Trial Blueprint recommendations that endpoints should be fine-tuned based on the pilot phase of a trial.<br><br>2. Power calculations were revised in light of pilot data on hospitalisation rates and rates of loss to follow up.<br>3. New secondary endpoint in light of change to primary endpoint. |
| 6.0 | 14Aug2020 | Lucy Cureton (as advised by MHRA) | Clarified role of ECG in section 9.6 baseline assessments. | After reviewing the previous amendment, the MHRA noted that this section had not been updated to bring it in line with the change to the exclusion in the last version of the protocol. |
| 5.0 | 07Jul2020 | Timothy Hinks and Joanna Black | 1. Updated inclusion criteria to include symptom duration.<br><br>2. Updated inclusion criteria to include confirmed COVID cases.<br><br>3. Updated exclusion criteria to remove exclusion of patients taking SSRIs. | 1. Recommendation of the DSMC.<br><br>2. Due to the wider availability of PCR testing for SARS-CoV-2, including contact screening and point of care testing, the PIs have requested clarification on the inclusion criteria to prevent inadvertent exclusion of those with a compatible clinical syndrome and PCR proven infection.<br><br>3. SSRIs are not contra-indicated with AZM. The protocol has robust monitoring for cardiovascular events, in addition the protocol excludes patients with a QTc prolongation greater than 480ms. A further detailed justification has |

© Copyright: The University of Oxford and Oxford University Hospitals NHS Foundation Trust 2019

| | | | | |
|---|---|---|---|---|
| | | | 4. Added reporting all cardiovascular events irrespective of causality.<br><br>5. Clarification to reporting procedures and timelines for SAEs.<br><br>6. Section 11: removed the sentence to take two 250mg capsules. | already been agreed by the MHRA. This MHRA correspondence is attached to the amendment application to further support the justification to the change in the exclusion.<br><br>4. Pfizer have a special interest in these events and have requested these as a condition of IMP supply.<br>5. Clarification to ensure accurate reporting and timelines.<br><br>6. Protocol is clear that the dose is 500mg. We removed the sentence two 250mg capsules, to allow for any change in capsule strength for example supply of 500mg tablets. |
| 4.0 | 29June2020 | | Not listed, as MHRA gave GNA. | |
| 3.0 | 07May2020 | Timothy Hinks and Susan Dutton | Updated following meeting of TSC/DSMC requesting clarity on analyses to be undertaken.<br><br>Typographical changes made<br>Reduction of number of optional blood tubes to be taken.<br>Inclusion at baseline data collection for females' questions about pregnancy and lactation, and data on any other COVID-19 trials participants enter if admitted to hospital. | |
| 2.0 | 30Apr2020 | Timothy Hinks | Updated with changes requested by the REC, specifically more information on pregnancy and the risks of the trial (section 11.1.6) and some potential drug interactions (section 11.1.5) specifically | |

© Copyright: The University of Oxford and Oxford University Hospitals NHS Foundation Trust 2019

| | | | |
|---|---|---|---|
| | | | Coumarin-type oral anticoagulants, Nelfinavir, Trimethoprim/ Sulfamethoxazole. |
| 1.0 | 23Apr2020 | First issue | |

© Copyright: The University of Oxford and Oxford University Hospitals NHS Foundation Trust 2019

#### Appendix 2: Statistical Analysis Plan V1.0 18Mar2021

### ATOMIC2 – A multi-centre open-label two-arm randomised superiority clinical trial of Azithromycin versus usual care In Ambulatory COVID-19

#### Statistical Analysis Plan

Version 1.0 – 18Mar2021

Based on Protocol version V7.0_04Feb2021

Trial registration: IRAS ID: 282892

| Role | Name | Title | Signature | Date |
|---|---|---|---|---|
| Author | Ariel Wang | Trial Statistician | | 18Mar2021 |
| Senior Statistician | Ruth Knight | Senior Statistician | | 18Mar2021 |
| Lead Statistician | Susan Dutton | Lead Statistician | | 18Mar2021 |
| Chief Investigator | Dr Timothy SC Hinks | Chief Investigator | | 18Mar2021 |

**Oxford Clinical Trials Research Unit (OCTRU)**
**Centre for Statistics in Medicine (CSM)**

### 1. INTRODUCTION

This document details the proposed data presentation and analysis for the main paper(s) and final study reports from ***A multi-centre open-label two-arm randomised superiority clinical trial of Azithromycin versus usual care In Ambulatory COVID-19 (ATOMIC2)***. The results reported in these papers should follow the strategy set out here. Subsequent analyses of a more exploratory nature will not be bound by this strategy, though they are expected to follow the broad principles laid down here. The principles are not intended to curtail exploratory analysis (for example, to decide cut-points for categorisation of continuous variables), nor to prohibit accepted practices (for example, data transformation prior to analysis), but they are intended to establish the rules that will be followed, as closely as possible, when analysing and reporting the trial.

The analysis strategy will be available on request when the principal papers are submitted for publication in a journal. Suggestions for subsequent analyses by journal editors or referees, will be considered carefully, and carried out as far as possible in line with the principles of this analysis strategy; if reported, the source of the suggestion will be acknowledged.

Any deviations from the statistical analysis plan will be described and justified in the final report of the trial. The analysis should be carried out by an identified, appropriately qualified and experienced statistician, who should ensure the integrity of the data during their processing. Examples of such procedures include quality control and evaluation procedures.

#### 1.1 Key personnel

**Author(s) (Trial statistician(s))**

Ariel Wang

**Reviewers (Trial Manager, DSMC, TSC, Statistician as appropriate)**

**Trial Manager**

Lucy Cureton

**Senior Statistician**

Ruth Knight

**DSMC**

Prof James Chalmers (Chair)

Professor Chris Rogers (Independent member)

Peter Howarth (Independent member)

**TSC**

Prof Paul Little (Chair)

Dr Timothy Hinks (Chief Investigator)

Mike Bradburn (Independent member)

Peter Mcquitty (Independent member)

Najib Rahman (Non-independent member)

Dominic Shaw (Independent member)

**Approver (Senior Statistician, Chief Investigator)**

**Senior Statistician (Lead Statistician)**

Susan Dutton

**Chief Investigator**

Dr Timothy Hinks

#### 1.2 Changes from previous version of SAP

A summary of key changes from earlier versions of SAP, with particular relevance to protocol changes that have an impact on the design, definition, sample size, data quality/collection and analysis of the outcomes will be provided. Include protocol version number and date.

| Version number<br>Issue date | Author of this issue | Protocol Version & Issue date | Significant changes from previous version together with reasons |
|---|---|---|---|
| V1.0_18Mar2021 | Ariel Wang | Protocol_ V7.0_04Feb2021 | Not applicable as this is the 1st issue |
| | | | *Add to or delete as required* |

### 2. BACKGROUND AND OBJECTIVES

#### 2.1 Background and rationale

Coronavirus-induced disease 2019 (COVID-19) is an infection caused by a virus whose full name is severe acute respiratory syndrome coronavirus 2 (SARS-CoV-2). This is a new and rapidly-spreading infectious disease. Those people that come into contact with the virus can have symptoms such as a mild fatigue, fever, loss of taste and a persistent cough, which can develop into severe respiratory failure requiring hospitalisation and mechanical ventilation. For those where the symptoms worsen this typically occurs 1 to 2 weeks into coming in contact with the virus. This provides a window of opportunity to potentially treat those patients who present with symptoms before becoming seriously ill to take a drug that might result in them not developing the severe symptoms.

The ATOMIC2 study is investigating if a common antibiotic called Azithromycin (AZM) may prevent the patients from getting worse. Azithromycin (AZM) is an orally active synthetic macrolide antibiotic with a wide range of antibacterial, anti-inflammatory and antiviral properties. It is a safe, inexpensive, generic licensed drug available worldwide, on the WHO list of essential medications, and manufactured to scale and therefore an ideal candidate molecule to be repurposed as a potential candidate therapy for pandemic COVID-19. Macrolides, particularly Azithromycin, were used to treat 1/3 of severe cases of MERS-CoV[1] and Azithromycin has been tried in COVID-19 infection[2] although RCT data are lacking[3].

#### 2.2 Objectives

In this study, researchers want to investigate this medicine in patients who have mild symptoms of COVID-19 who go to the hospital, but who doctors decide there is no need to admit them for treatment. The study will investigate whether if half the patients are given Azithromycin for 14 days and half the patients do not receive Azithromycin, there are less people after 28 days in one of the groups who go on to develop more severe symptoms from COVID-19.

The primary outcome was updated following the interim analysis of 109 participants reaching the 28 day time-point (23Nov2020). In line with the advice from DSMC (07Dec2020) and TSC (27Jan2021) and in accordance to the recommendations of the World Health Organisation Blueprint for Covid-19 Therapeutic Trials[4] that the primary endpoint should be responsive to the eligible patient population and the definition of the endpoint should be fine-tuned for the Pivotal Phase, based on the Pilot Phase of the Trial. Following further assessment of the blinded data the primary outcome has been updated from:

'To compare the effect of Azithromycin in participants with a clinical diagnosis of COVID-19 in reducing the proportion with either death or hospital admission with respiratory failure requiring Non-Invasive Mechanical Ventilation (NIV) or Invasive Mechanical Ventilation (IMV) over the 28 days from randomisation.

To:

'To compare the effect of Azithromycin in participants with a clinical diagnosis of COVID-19 in reducing the proportion with death or hospital admission from any cause over the 28 days from randomisation.'

Note that the updated primary outcome, death or all-cause hospitalisation, includes the original primary endpoint, death or hospitalisation requiring level 2 or 3 ventilation, and the latter will still be reported as a secondary outcome at the end of the trial. The primary and secondary endpoints and objectives for this study are as described in Table 1.

**Table 1**: Primary and secondary objectives and endpoints

| Objectives | Outcome Measures | Time point(s) |
|---|---|---|
| **Primary Objective**<br>To compare the effect of Azithromycin in participants with a clinical diagnosis of COVID-19 in reducing the proportion with either death or hospital admission from any cause over the 28 days from randomisation. | Efficacy will be determined through differences in the proportion with either death or requiring hospital admission, from any cause, over the 28 days from randomisation | Determined at day 28 from randomisation. |
| **Secondary Objectives** | | |
| To compare the effect of Azithromycin in participants with a clinical diagnosis of COVID-19 in reducing the proportion with either death or hospital admission with respiratory failure requiring invasive or non-invasive mechanical ventilation over 28 days from randomisation. | Efficacy will be determined through differences in the proportion with either death or admission with respiratory failure requiring level 2 ventilation (NIV/CPAP/nasal high-flow) or level 3 (invasive mechanical ventilation) in the 28 days from randomisation. | Determined at day 28 from randomisation. |
| To compare the effect of Azithromycin in participants with a PCR-confirmed diagnosis of COVID-19 in reducing the proportion with either death or hospital admission with respiratory failure requiring invasive or non-invasive mechanical ventilatory support over 28 days from randomisation (for those who had a COVID-19 swab at randomisation) | Efficacy will be determined through differences in the proportion with either death or admission with respiratory failure requiring level 2 ventilatory support (NIV/CPAP/nasal high-flow) or level 3 (invasive mechanical ventilation) in the 28 days from randomisation using a retrospective analysis of COVID-19 oropharyngeal swabs for those who had one taken at time of randomisation. | Determined at day 28 from randomisation. |
| To compare the effect of Azithromycin in participants with a PCR-confirmed diagnosis of COVID-19 in reducing the proportion with either death or all-cause hospital admission (for those who had a COVID-19 swab at randomisation) | Efficacy will be determined through differences in the proportion with either death or all-cause hospital admission in the 28 days from randomisation using a retrospective analysis of COVID-19 oropharyngeal swabs for those who had one taken at time of randomisation. | Determined at day 28 from randomisation. |
| To compare differences in all-cause mortality. | Data on vital status (alive / dead, with date and presumed cause of death if appropriate) at 28 days from randomisation | Ascertain data at 28 days after randomisation. |
| To compare differences in proportion progressing to pneumonia. | Progression to pneumonia as diagnosed by chest x-ray (or CT thorax), with compatible clinical findings, if no pneumonia is present at time of enrolment. To be diagnosed by a medically qualified doctor and data obtained from review of case-notes and relevant radiology. | Ascertain this information at time of pneumonia diagnosis, or at 28 days after randomisation (whichever is sooner) |

| Objectives | Outcome Measures | Time point(s) |
|---|---|---|
| To compare differences in proportion progressing to severe pneumonia. | Evolution of pneumonia, as diagnosed by chest x-ray or CT thorax, if pneumonia is present at time of enrolment. To be diagnosed by a medically qualified doctor and data obtained from review of case-notes and relevant radiology. Severe pneumonia is defined as BTS CURB-65 score of 3-5. | Ascertain this information retrospectively at 28 days after randomisation |
| To compare differences in peak severity of illness. | The scoring system is described in section 6.1 and reflects the severity of respiratory illness. The maximum severity score during the entire study period will be compared. | Ascertain from day 14 and day 28 telephone call and from retrospective ePR/medical notes data at 28 days after randomisation. |
| Safety and tolerability | Serious adverse events and concomitant medications. Record at enrolment, emergently during study period and proactively elicit at day 14 and at day 28. | Emergent data collection days 0-28 and elicit proactively at day 14 and day 28 post randomisation. |
| **Exploratory Objectives** Mechanistic analysis of blood and nasal biomarkers if available | The following samples may be taken. Blood for serum, Tempus tube (whole blood RNA), EDTA tubes (PBMC), nasal brush to be placed immediately into RNA lysis buffer (for subsequent PCR and transcriptomic analysis). | Samples to be collected prospectively at baseline and again if patient admitted, to be taken as soon as possible and within 72 hours of admission if possible. |

### 3. STUDY METHODS

#### 3.1 Trial Design/framework

ATOMIC2 is a multi-centre, prospective open label two-arm randomised superiority clinical trial of standard care and Azithromycin versus standard care alone for those presenting to hospital with COVID-19 symptoms who are not admitted at initial presentation.

We will perform a study of the efficacy of AZM to prevent and/or reduce the severity of lower respiratory tract illness in adult patients with clinically diagnosed COVID-19 infection being assessed in secondary care but initially managed on an ambulatory care pathway. This provides a therapeutic window of opportunity to avert development of more severe disease. Participants will be randomised to receive Azithromycin 500 mg daily for 14 days or standard care. The first dose will be within 4 hours of randomisation.

#### 3.2 Randomisation and Blinding

Eligible patients will be randomised using the centralised validated computer randomisation program through a secure (encrypted) web-based service, RRAMP (https://rramp.octru.ox.ac.uk), provided by the Oxford Clinical Trials Research Unit (OCTRU) with a minimisation algorithm to ensure balanced allocation across treatment groups, stratified by centre, hypertension (yes/no), diabetes (yes/no) and sex (male/female) in a

1:1 ratio to either Azithromycin (AZM) or usual care. To ensure the unpredictability of treatment allocation the minimisation algorithm will include a probabilistic element and a small number of participants will be randomised by simple randomisation to seed the system.

Note: Hypertension is defined as any hypertension previously diagnosed by a doctor prior to presenting to the hospital with COVID-19 symptoms.

Note: Diabetes is defined as any diabetes that is treated with oral or injectable therapy.

Stratification by centre will help to ensure that any centre-effect will be equally distributed in the trial arms and enable practical issues associated with the active intervention to be overcome.

There is some emerging evidence that patients who have underlying hypertension, diabetes and are male are more likely to progress and require hospitalisation and have an increased mortality, so it is important for the two treatments to be balanced across these potentially important prognostic factors.

Note: Eligibility for a patient to enter the trial must be confirmed by a medically qualified doctor.

Full details of the randomisation are available in ATOMIC2_RBP_v1.0_05May2020, stored in the confidential statistical section of the TMF.

ATOMIC2 is an open label study without blinding.

##### 3.3 Sample Size: initial estimate, pilot phase

The definitive trial will recruit approximately 800 participants (400 per arm). An interim analysis has been built into this trial after an initial 100 patients have been randomised, treated and followed-up for 28 days. The DSMC will review the accruing safety and efficacy data and the results from a futility analysis, which will assess whether the trial would be likely to confirm superiority of the active treatment if it was to continue as planned. They will provide recommendations to the TSC as to whether the trial should continue to the definitive trial or stop early for safety or futility. If the recommendation is to continue to the definitive trial they will also review the assumptions on which the sample size is based and the final sample size for the definitive study will be confirmed by the DSMC and TSC. This is a rapidly evolving disease area and information about the control rate for progression to hospitalisation or death is not yet fully known. More details will be available during the trial and will be assessed to refine the sample size. Initial assumptions are described below.

The total sample size of 778 participants with primary outcome data are required to reject the null hypothesis of no difference between the active treatment and usual care. This number is based on the following assumptions: 30-40% of patients following usual care will progress to hospitalisation or death within 28 days; 90% power, 2-sided 5% significance and a 30-35% reduction in progression or death for patients on Azithromycin (778 participants required to detect a 33.3% reduction from 30% to 20% in progression to hospitalisation or death or 646 required to detect a 30% reduction from 40% to 28% in progression to hospitalisation or death). To allow for uncertainty around the assumptions and allowing for a 2% loss to follow-up, approximately 800 participants will be required. Total sample size for the definitive study will be refined at the interim analysis if progression to the full trial is the option chosen.

Checks of the sample size calculations are detailed in the document ATOMIC2_Samplesize_08Apr2020_SD.rtf stored in the confidential statistical section of the TMF.

##### 3.4 Sample size: revised estimate and change in primary outcome for pivotal phase

Data from the first 109 participants reaching the 28 day primary outcome time-point was reviewed by the DSMC 07Dec2020. At this time no participants had been admitted to hospital requiring level 2 or 3 ventilatory support (the primary outcome), although some participants had been admitted to hospital. The DSMC recommended to the TSC and TMG that the primary outcome should be reviewed in order for the trial to continue and enable it to answer the research question and that any updated primary outcome should not be subjective as the trial is not blinded.

Following further assessment of the blinded data the Research question has been updated from:

'To compare the effect of Azithromycin in participants with a clinical diagnosis of COVID-19 in reducing the proportion with either death or hospital admission with respiratory failure requiring Non-Invasive Mechanical Ventilation (NIV) or Invasive Mechanical Ventilation (IMV) over the 28 days from randomisation.

To:

'To compare the effect of Azithromycin in participants with a clinical diagnosis of COVID-19 in reducing the proportion with either death or hospital admission from any cause over the 28 days from randomisation.'

Note: that the updated primary outcome, death or all-cause hospitalisation, includes the original primary endpoint, death or hospitalisation requiring level 2 or 3 ventilation, and the latter will still be reported as a secondary outcome at the end of the trial.

Following this change to the primary outcome and based on blinded data from the pilot phase of the study the sample size has been updated:

Assuming a rate of all cause hospitalisation/death of 15% in the usual care arm, then a minimum of 276 participants providing primary end-point data, will provide 80% power and 5% (2-sided) significance to detect a difference from 15% to 5% in the Azithromycin arm, a relative reduction of 66%. Allowing for 10% loss to follow-up, this number is increased to a minimum of 308 participants. If additional participants are recruited (this could potentially occur with the current increase in prevalence of COVID-19 and the rapid recruitment of participants) this will provide more power to estimate the treatment effects with the potential to detect a smaller difference if one exists.

##### 3.5 Statistical Interim Analysis, Data Review and Stopping guidelines

The Data and Safety Monitoring Committee (DSMC) is a group of independent experts external to the trial who will assess the progress, conduct, participant safety and, if required, critical endpoints. The DSMC follows the charter as described in the document DSMC Charter_ATOMIC2_V1.0_22May2020. docx stored in the TMF. The DSMC will review the unblinded futility interim analysis and determine if progression to the full trial is recommended after the first 100 participants have completed their 28 days follow-up. Following this interim review, the trial will be stopped for safety or futility (i.e. the trial is unlikely to find that Azithromycin is superior). If not stopped early, the assumptions on which the sample size is based will be reviewed and the final sample size for the main study will be confirmed by the DSMC and TSC. If the result from the futility analysis is uncertain, the DSMC and TSC may advise another futility interim analysis after a minimum of another 200 patients have completed their 28 days follow-up. Additional futility analyses will be undertaken at the request of the DSMC. The meetings will be online virtual conference whenever possible, with teleconference as a second option. Each DSMC meeting will have a mixture of open and closed sessions. Only

DSMC members and others whom they specifically invite (e.g. the Trial Statistician) should be present in the closed sessions.

Note: Whilst the interim analysis is prepared for and undertaken, and the DSMC meeting held – recruitment will continue to the trial as per protocol.

If the recommendation is to continue to the main trial they will also review the assumption on which the sample size is based and the final sample size for the definitive study will be confirmed by the DSMC and TSC. This is a rapidly evolving disease area and information about the control rate for progression to hospitalisation or death is not yet fully known. More details will be available during the trial and will be assessed to refine the sample size.

The interim futility analysis is based on Bayesian predictive probabilities. Bayesian methods are useful for interim futility monitoring based on the probability of observing a statistically significant treatment effect if the trial were to continue to its predefined maximum sample size[5-7]. The predictive probability of success (PPS) is the probability of achieving a successful (significant) result at a future analysis, given the current unblinded interim data[8-10]. If the PPS is sufficiently small, < 0.1, at the maximum sample size, the trial will be stopped for futility.

Table 2 shows simulation results for various scenarios and the probability of stopping and claiming futility at each interim analysis, based on 1000 simulated trials for each scenario and interim analyses after 100, 250, 350, 500, and 600 participants have been followed for 28 days. The results demonstrated that interim analyses after the first 100 and 200 participants are unlikely to stop the trial for futility (i.e. probability of claiming futility = 0). It also shows that if there is no difference between control and treatment groups (i.e. scenarios 1, 3, 5 and 7), the trial will stop for futility around 30% of the time at an interim analysis with 350 participants. For each scenario, the accumulated probability of stopping for futility at each interim (if it had not stopped earlier) could be calculated by adding the probability at the current interim and the previous interims. This cumulative probability of stopping for futility at each interim is also shown in Figure 1. The dashed lines show scenarios where treatment and control response rates are the same (i.e. scenarios 1, 3, 5 and 7), and the solid lines show scenarios (i.e. scenarios 2, 4, 6 and 8) where the response rate in the treatment group is less than the control. In Figure 1, also given are the average trial size and the power, i.e. the probability of claiming success (rejecting the null at the 5% level) at the end of the trial. Under the null, this gives the error rate. The average trial size for each scenario presented in Figure 1 is calculated as the sum of the probability of stopping at each interim * the size of each interim. In the simulations, the sizes of each interim are 100, 200, 350, 500, 600 and 778.

Details of the simulations for interim analyses and results interpretation is attached in Appendix C.

Full details of the interim analyses planned are available in the Interim Statistical Analysis Plan, ATOMIC2_Interim Statistical Analysis Plan_V1.0_19Oct2020 stored in the confidential statistical section of the TMF.

**Table 2: Predictive probability of success at each interim analysis**

| | | Probability of claiming futility | | | | | Overall probability of success |
|---|---|---|---|---|---|---|---|
| Scenario | Interim analysis | $n_1$=100 | $n_2$=250 | $n_3$=350 | $n_4$=500 | $n_6$=600 | |
| 1 | $\theta_c = 0.2, \theta_{t=}0.2$ | 0 | 0 | 0.30 | 0.37 | 0.12 | 0.04 |

| | | | | | | | |
|---|---|---|---|---|---|---|---|
| 2 | $\theta_c = 0.2, \theta_{t=}0.14$ | 0 | 0 | 0.10 | 0.12 | 0.07 | 0.53 |
| 3 | $\theta_c = 0.25, \theta_{t=}0.25$ | 0 | 0 | 0.30 | 0.36 | 0.16 | 0.04 |
| 4 | $\theta_c = 0.25, \theta_{t=}018$ | 0 | 0 | 0.09 | 0.08 | 0.05 | 0.61 |
| 5 | $\theta_c = 0.3, \theta_{t=}0.3$ | 0 | 0 | 0.29 | 0.36 | 0.15 | 0.04 |
| 6 | $\theta_c = 0.3, \theta_{t=}0.2$ | 0 | 0 | 0.03 | 0.02 | 0.01 | 0.88 |
| 7 | $\theta_c = 0.4, \theta_{t=}0.4$ | 0 | 0 | 0.26 | 0.37 | 0.16 | 0.05 |
| 8 | $\theta_c = 0.4, \theta_{t=}0.28$ | 0 | 0 | 0.02 | 0.01 | 0.01 | 0.91 |

**Figure 1: Properties of the trial with interims at 100, 250, 350, 500, 600 and a PPS threshold of 0.1**

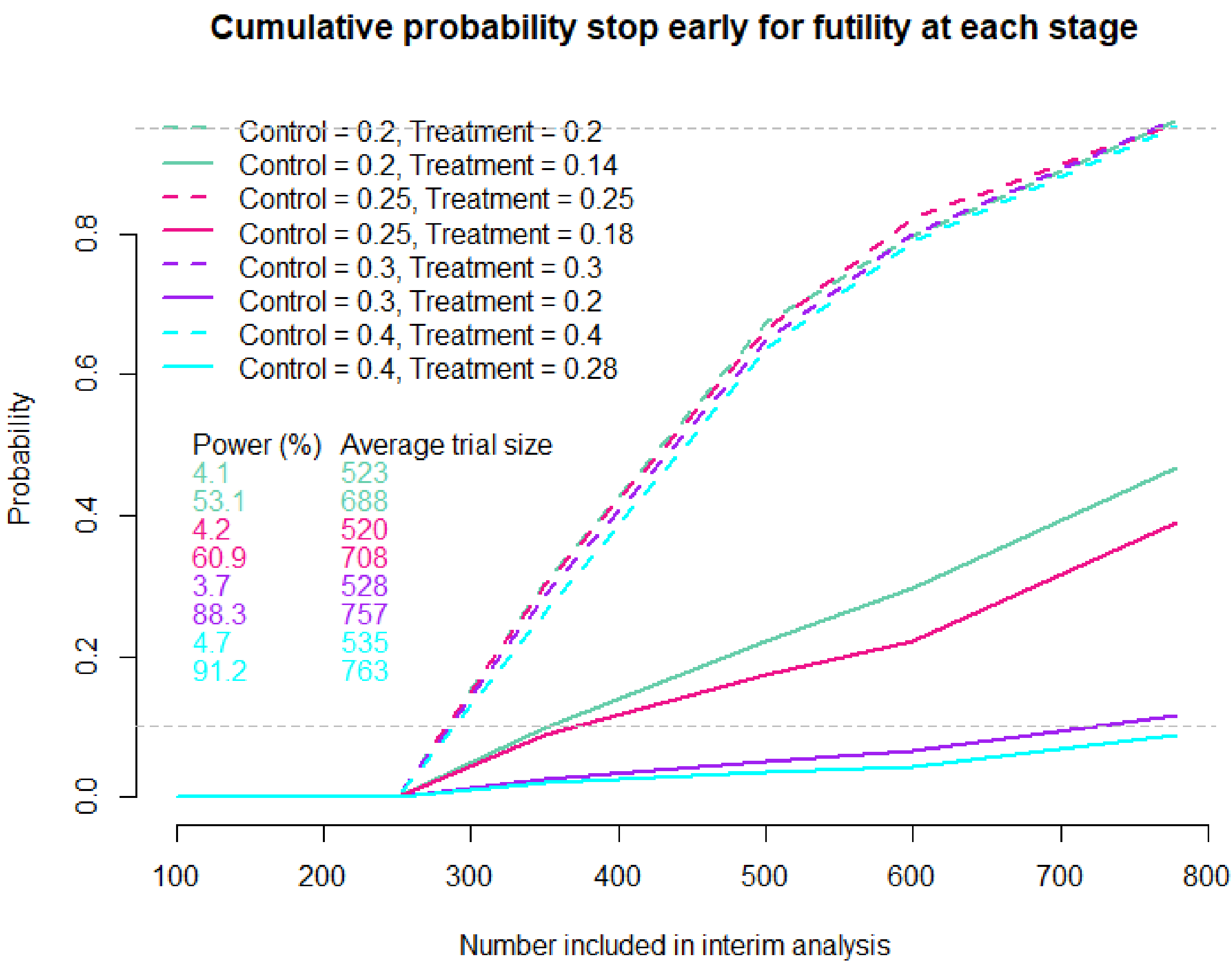

Data from the first 109 participants reaching the 28day primary outcome time-point was reviewed by the DSMC. At this time no participants had been admitted to hospital requiring level 2 or 3 ventilatory support or died (the primary outcome). Therefore, no formal comparative interim analysis was undertaken.

##### 3.6 Timing of Final Analysis

The final analysis of all primary and secondary endpoints will be conducted together when all recruited patients have completed all follow-up.

##### 3.7 Blinded review

A blinded review of the data (not separated by treatment group) will be undertaken prior to the final data lock in order to look into the distribution of variables, missing data distributions, outliers, and to finalise the per protocol population.

##### 3.8 Statistical Analysis Outline for publication

Standard descriptive statistics will be used to describe the demographics between the treatment groups reporting means and standard deviations or medians and interquartile ranges as appropriate for continuous variables and numbers and percentages for binary and categorical variables.

The proportion of patients progressing to hospitalisation or death by day 28 post-randomisation is the primary outcome for this study. The difference in proportions between the treatment arms will be assessed using a chi-squared tests and a 5% (2-sided) significance level. Difference in proportions together with the 95% confidence intervals will be reported. The principal analysis on which the success (or otherwise) of the trial is the adjusted analyses which will be undertaken using logistic regression with progression as the binary outcome, adjusting for stratification factors (centre, hypertension, diabetes and sex). Hypertension, diabetes and sex will be adjusted for as fixed effects. Centre will be included as a random effect. Additional adjustment for other important prognostic variables (age, chronic pulmonary disease, and presence of cancer which will be determined by the presence of any of the following four items: malignancy, leukaemia, lymphoma and metastatic solid tumour in the Charlson comorbidity index) will be undertaken as a supporting analysis if sufficient events are observed to allow for including additional covariates. Time to event analysis will also be undertaken to explore whether the active treatment delays progression. Both relative and absolute differences in proportions will be reported together with 95% confidence intervals.

Other binary outcomes will be assessed using similar methods and continuous variables will be assessed using linear regression analysis. Where appropriate longitudinal methods will incorporate different time points.

The number and percentage of subjects with each score on the severity scale for clinical improvement will be presented at baseline and each post baseline time point. In addition, the change in severity scale score from baseline will be summarised on both a categorical scale, using counts and percentages and on a continuous scale using descriptive statistics. Inferential statistical analyses such as ordered logistic regression, mixed model for repeated measurement (MMRM) or Mann-Whitney may be conducted in an exploratory fashion to aid the understanding of the data. Binary interpretations of the severity scale for clinical improvement may also be defined in the SAP, such as responders (any improvement at day 14) and complete responders (score of 0 at day 14) and these will be compared using logistic regression as for the primary outcome.

Reporting of the results will be based on the CONSORT statement[11].

It is anticipated that all statistical analysis will be undertaken using Stata (StataCorp LP, **www.stata.com**) or other well-validated statistical packages.

### 4. STATISTICAL PRINCIPLES

#### 4.1 Statistical Significance and Multiple Testing

No multiple testing will be undertaken as a single primary outcome is considered. Therefore, the significance level will be 0.05 and 95% confidence intervals will be reported.

All secondary analyses will be considered as supporting the primary analysis and will be analysed using a significance level of 0.05 with 95% confidence intervals.

The DSMC will regularly review accruing data for safety and comparative formal interim analyses of the primary outcome to determine futility (see section 3.5).

#### 4.2 Definition of Analysis Populations

The intention-to-treat (ITT) population is defined as all randomised patients analysed according to their randomised allocation.

A supplementary ITT population (ITT +ve) is defined as all randomised patients with a positive baseline COVID-19 test determined based on baseline swabs.

All efficacy and safety analyses will be based on the ITT population and repeated on the ITT +ve population (if sufficient numbers in this subgroup are available).

### 5. TRIAL POPULATION AND DESCRIPTIVE ANALYSES

#### 5.1 Enrolment of Study Sample and Patient Flow

The participant flow through the trial will be summarised as outlined in Figure 2. This will include the number of individuals screened, eligible, randomised to each group, receiving allocated treatment and included in the analysis as suggested in the CONSORT guidelines. Reasons for ineligibility, loss to follow-up and exclusion from the analysis will be summarised, as will the number of patients who withdraw before each analysis time point.

**Figure 2:** CONSORT flow diagram for participants in trial.

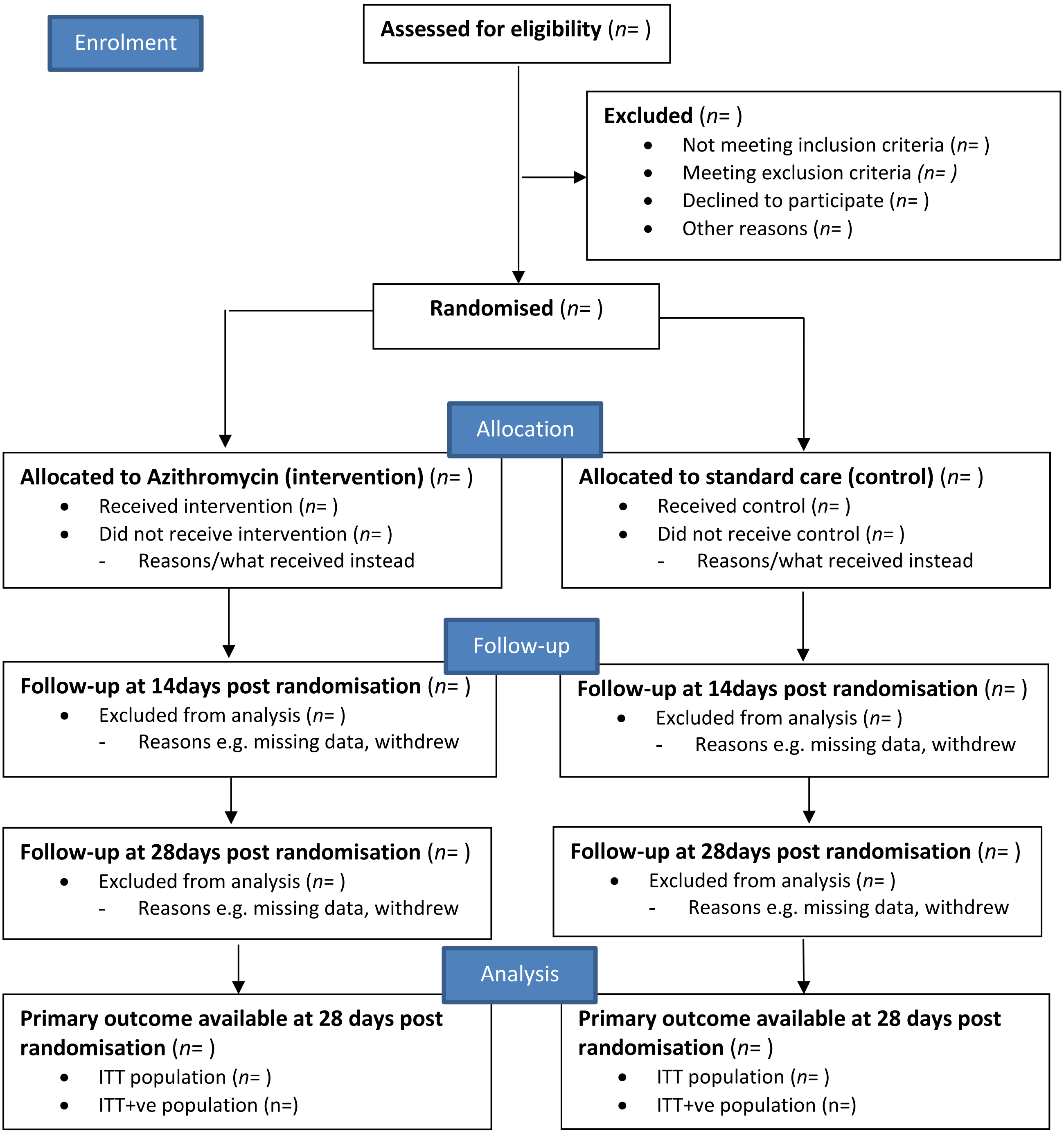

#### 5.2 Withdrawal from treatment and/or follow-up

The numbers (and percentages) of withdrawals by each time point will be reported by treatment group along with reasons for these withdrawals. These will be summarised as outlined in Tables 3&4. Any differential losses will be investigated. Any deaths that are not due to COVID-19 will be reported separately.

**Table 3:** Summary of withdrawals split by treatment group

| | Intervention | Control | Total |
|---|---|---|---|
| Number of withdrawals | X (%) | X (%) | X (%) |
| Median (IQR) time to withdrawal (days) | X (X, X) | X (X, X) | X (X, X) |
| Principal reasons of withdrawal | | | |
| Clinician request | X (%) | X (%) | X (%) |
| Participant did not like allocation | X (%) | X (%) | X (%) |
| Participant felt too ill to continue in study | X (%) | X (%) | X (%) |
| Other reason | X (%) | X (%) | X (%) |

**Table 4:** Details of withdrawals from follow-up (and reasons) split by treatment group

| **Treatment** | **Patient ID** | **Date of withdrawal (DDMonYYYY)** | **Reasons for withdrawal** | **Latest timepoint when information is available (day 0, day 14 or day 28)** | **Data available for outcome analysis** |
|---|---|---|---|---|---|
| **Intervention** | ID 1 | | | | Y/N |
| | ID 2 | | | | Y/N |
| | … | | | | Y/N |
| **Control** | ID 1 | | | | Y/N |
| | ID 2 | | | | Y/N |
| | … | | | | Y/N |

##### 5.3 Baseline Comparability of Randomised Groups

The baseline comparability of the two treatment groups (Azithromycin versus standard care) will be considered. The treatment groups will be reported in terms of stratification factors (see Table 5), baseline demographic characteristics (see Table 6), secondary outcome measures (where appropriate) at baseline (see Table 7), and baseline demographic characteristics by SWAB results (see Table 8). Numbers (with percentages) for binary and categorical variables and the number of available observations with means (and standard deviations), medians (with inter-quartile ranges) and range for continuous variables will be reported. These will be reported for each treatment group and overall. There will be no tests of statistical significance nor confidence intervals for differences between randomised groups on any baseline variable. Comparison of the demographic characteristics of the participants in the total population with the subset of the population who are hospitalised or died (primary outcome) will also be reported (Supplementary Table S1).

**Table 5:** Stratification factors at baseline – split by treatment group and overall

| **Stratification factor** | **Azithromycin** | | **Standard Care** | | **Total** | |
|---|---|---|---|---|---|---|
| | n | % | n | % | n | % |
| **Centre** | | | | | | |
| Basingstoke | X | X | X | X | X | X |
| Burnley Hospital | X | X | X | X | X | X |
| City Hospital | X | X | X | X | X | X |
| Darlington Memorial Hospital | X | X | X | X | X | X |
| Denmark Hill | X | X | X | X | X | X |
| Dundee | X | X | X | X | X | X |
| Horton | X | X | X | X | X | X |
| James Cook University Hospital | X | X | X | X | X | X |
| North Tees | X | X | X | X | X | X |

| | | | | | | |
|---|---|---|---|---|---|---|
| Oxford | X | X | X | X | X | X |
| Princess Royal University Hospital | X | X | X | X | X | X |
| Royal Berkshire Hospital | X | X | X | X | X | X |
| Royal Blackburn Hospital | X | X | X | X | X | X |
| Royal Derby Hospital | X | X | X | X | X | X |
| Royal London Hospital | X | X | X | X | X | X |
| St George's Hospital | X | X | X | X | X | X |
| University College London Hospitals | X | X | X | X | X | X |
| University Hospital Llandough | X | X | X | X | X | X |
| University Hospital Wales | X | X | X | X | X | X |
| **Hypertension** | | | | | | |
| Yes | X | X | X | X | X | X |
| No | X | X | X | X | X | X |
| **Diabetes** | | | | | | |
| Yes | X | X | X | X | X | X |
| No | X | X | X | X | X | X |
| **Sex** | | | | | | |
| Male | X | X | X | X | X | X |
| Female | X | X | X | X | X | X |

**Table 6:** Baseline characteristics of participants – split by treatment group and overall (ITT population)

| | **Azithromycin (N=XX)** | | | **Standard care (N=XX)** | | | **Total (N=XX)** | | |
|---|---|---|---|---|---|---|---|---|---|
| | Mean (SD) | Median (IQR) | Range | Mean (SD) | Median (IQR) | Range | Mean (SD) | Median (IQR) | range |
| **Age (years)** | X (X) | X (X,X) | (X,X) | X (X) | X (X,X) | (X,X) | X (X) | X (X,X) | (X,X) |
| **COVID COS Score †** | X (X) | X (X,X) | (X,X) | X (X) | X (X,X) | (X,X) | X (X) | X (X,X) | (X,X) |
| **COVID COS PLUS Score †** | X (X) | X (X,X) | (X,X) | X (X) | X (X,X) | (X,X) | X (X) | X (X,X) | (X,X) |
| **Charlson Comorbidity Index (CCI)*** | X (X) | X (X,X) | (X,X) | X (X) | X (X,X) | (X,X) | X (X) | X (X,X) | (X,X) |

| | n | % | n | % | n | % |
|---|---|---|---|---|---|---|
| **Ethnicity** | | | | | | |
| White | X | X | X | X | X | X |
| Mixed | X | X | X | X | X | X |
| Asian or Asian British | X | X | X | X | X | X |
| Black or Black British | X | X | X | X | X | X |
| Other Ethnic Groups | X | X | X | X | X | X |
| **Residence** | | | | | | |
| Non-residential care | X | X | X | X | X | X |
| Residential Care | X | X | X | X | X | X |
| No fixed address (NFA) | X | X | X | X | X | X |
| **Live alone** | | | | | | |
| Yes | X | X | X | X | X | X |
| No | X | X | X | X | X | X |
| **Smoking** | | | | | | |
| Never smoked | X | X | X | X | X | X |
| Ex-smoker | X | X | X | X | X | X |
| Current smoker | X | X | X | X | X | X |
| Ex-smoker, current vaper | X | X | X | X | X | X |

| | | | | | | |
|---|---|---|---|---|---|---|
| Never smoked, current vaper | X | X | X | X | X | X |
| Not recorded | X | X | X | X | X | X |
| **Work Status** | | | | | | |
| Retired | X | X | X | X | X | X |
| Working | X | X | X | X | X | X |
| Houseperson | X | X | X | X | X | X |
| **Occupation** | | | | | | |
| Not Healthcare related | X | X | X | X | X | X |
| Healthcare worker | X | X | X | X | X | X |
| Laboratory worker | X | X | X | X | X | X |
| **Have asthma** | | | | | | |
| Yes | X | X | X | X | X | X |
| No | X | X | X | X | X | X |
| **History of previous myocardial infarction** | | | | | | |
| Yes | X | X | X | X | X | X |
| No | X | X | X | X | X | X |
| **Currently undergoing any cancer treatment** | | | | | | |
| Yes | X | X | X | X | X | X |
| No | X | X | X | X | X | X |
| **Have chronic pulmonary disease** | | | | | | |
| Yes | X | X | X | X | X | X |
| No | X | X | X | X | X | X |

† COVID-19 COS Score of clinical symptoms is a total score of six common and important clinical symptoms, including fever, cough, fatigue, shortness of breath, diarrhoea, and body pain, each of which can be scored as 0 (no), 1 (mild), 2 (moderate), or 3 (significant). In ATOMIC2, an amended version COVID-19 COS PLUS is also designed with 2 extra clinical symptoms that also considered as having clinical importance: changes to sense of smell and loss of taste.

* The Charlson Comorbidity Index assigns a numerical value or "weight" from 1,2,3 or 6 to nineteen specific chronic illnesses. The final score (range 0-42) is simply the sum of weighted values with higher scores indicating more comorbidities.

**Table 7:** Clinical assessment outcome measure at baseline – split by treatment group and overall

| | **Azithromycin (N=XX)** | | **Standard care (N=XX)** | | **Total (N=XX)** | |
|---|---|---|---|---|---|---|
| | **mean (SD), median (IQR), range** | | **mean (SD), median (IQR), range** | | **mean (SD), median (IQR), range** | |
| **The Severity scale score** | X (X), X (X, X), (X, X) | | X (X), X (X, X), (X, X) | | X (X), X (X, X), (X, X) | |
| | n | % | n | % | n | % |
| **Pneumonia** | | | | | | |
| Yes | X | X | X | X | X | X |
| No | X | X | X | X | X | X |
| Not available | X | X | X | X | X | X |
| **SWAB results** | | | | | | |
| Positive | X | X | X | X | X | X |
| Negative | X | X | X | X | X | X |
| Failed Assay | X | X | X | X | X | X |
| Not available | X | X | X | X | X | X |

**Table 8:** Baseline characteristics of participants – split by SWAB results and overall

| | **SWAB +ve (N=XX)** | **SWAB -ve & no swab (N=XX)** | **Total (N=XX)** |
|---|---|---|---|

| | Mean (SD) | Median (IQR) | Range | Mean (SD) | Median (IQR) | Range | Mean (SD) | Median (IQR) | range |
|---|---|---|---|---|---|---|---|---|---|
| **Age (years)** | X (X) | X (X,X) | (X,X) | X (X) | X (X,X) | (X,X) | X (X) | X (X,X) | (X,X) |
| **COVID COS Score †** | X (X) | X (X,X) | (X,X) | X (X) | X (X,X) | (X,X) | X (X) | X (X,X) | (X,X) |
| **COVID COS PLUS Score †** | X (X) | X (X,X) | (X,X) | X (X) | X (X,X) | (X,X) | X (X) | X (X,X) | (X,X) |
| **Charlson Comorbidity Index (CCI)*** | X (X) | X (X,X) | (X,X) | X (X) | X (X,X) | (X,X) | X (X) | X (X,X) | (X,X) |
| | n | | % | n | | % | n | | % |
| **Ethnicity** | | | | | | | | | |
| White | X | | X | X | | X | X | | X |
| Mixed | X | | X | X | | X | X | | X |
| Asian or Asian British | X | | X | X | | X | X | | X |
| Black or Black British | X | | X | X | | X | X | | X |
| Other Ethnic Groups | X | | X | X | | X | X | | X |
| **Residence** | | | | | | | | | |
| Non-residential care | X | | X | X | | X | X | | X |
| Residential Care | X | | X | X | | X | X | | X |
| No fixed address (NFA) | X | | X | X | | X | X | | X |
| **Live alone** | | | | | | | | | |
| Yes | X | | X | X | | X | X | | X |
| No | X | | X | X | | X | X | | X |
| **Smoking** | | | | | | | | | |
| Never smoked | X | | X | X | | X | X | | X |
| Ex-smoker | X | | X | X | | X | X | | X |
| Current smoker | X | | X | X | | X | X | | X |
| Ex-smoker, current vaper | X | | X | X | | X | X | | X |
| Never smoked, current vaper | X | | X | X | | X | X | | X |
| Not recorded | X | | X | X | | X | X | | X |
| **Work Status** | | | | | | | | | |
| Retired | X | | X | X | | X | X | | X |
| Working | X | | X | X | | X | X | | X |
| Houseperson | X | | X | X | | X | X | | X |
| **Occupation** | | | | | | | | | |
| Not Healthcare related | X | | X | X | | X | X | | X |
| Healthcare worker | X | | X | X | | X | X | | X |
| Laboratory worker | X | | X | X | | X | X | | X |
| **Have asthma** | | | | | | | | | |
| Yes | X | | X | X | | X | X | | X |
| No | X | | X | X | | X | X | | X |
| **History of previous myocardial infarction** | | | | | | | | | |
| Yes | X | | X | X | | X | X | | X |
| No | X | | X | X | | X | X | | X |
| **Currently undergoing any cancer treatment** | | | | | | | | | |
| Yes | X | | X | X | | X | X | | X |
| No | X | | X | X | | X | X | | X |
| **Have chronic pulmonary disease** | | | | | | | | | |
| Yes | X | | X | X | | X | X | | X |
| No | X | | X | X | | X | X | | X |

† COVID-19 COS Score of clinical symptoms is a total score of six common and important clinical symptoms, including fever, cough, fatigue, shortness of breath, diarrhoea, and body pain, each of which can be scored as 0 (no), 1 (mild), 2 (moderate), or 3 (significant). In ATOMIC2, an amended version COVID-19 COS PLUS is also designed with 2 extra clinical symptoms that also considered as having clinical importance: changes to sense of smell and loss of taste.

* The Charlson Comorbidity Index assigns a numerical value or "weight" from 1,2,3 or 6 to nineteen specific chronic illnesses. The final score (range 0-42) is simply the sum of weighted values with higher scores indicating more comorbidities.

**Supplementary Table S1.** Demographic of participants – total population compared with those hospitalised

| | Hospitalised (N=XX) | | | Total (N=XX) | | |
|---|---|---|---|---|---|---|
| | Mean (SD) | Median (IQR) | Range | Mean (SD) | Median (IQR) | Range |
| **Age (years)** | X (X) | X (X,X) | (X,X) | X (X) | X (X,X) | (X,X) |
| **COVID COS Score at baseline †** | X (X) | X (X,X) | (X,X) | X (X) | X (X,X) | (X,X) |
| **COVID COS PLUS Score at baseline †** | X (X) | X (X,X) | (X,X) | X (X) | X (X,X) | (X,X) |
| **Severity scale score at baseline** | X (X) | X (X,X) | (X,X) | X (X) | X (X,X) | (X,X) |
| **Charlson Comorbidity Index (CCI) *** | X (X) | X (X,X) | (X,X) | X (X) | X (X,X) | (X,X) |
| | n | % | | n | % | |
| **Sex** | | | | | | |
| Male | X | X | | X | X | |
| Female | X | X | | X | X | |
| **Hypertension** | | | | | | |
| Yes | X | X | | X | X | |
| No | X | X | | X | X | |
| **Diabetes** | | | | | | |
| Yes | X | X | | X | X | |
| No | X | X | | X | X | |
| **Ethnicity** | | | | | | |
| White | X | X | | X | X | |
| Mixed | X | X | | X | X | |
| Asian or Asian British | X | X | | X | X | |
| Black or Black British | X | X | | X | X | |
| Other Ethnic Groups | X | X | | X | X | |
| **Residence** | | | | | | |
| Non-residential care | X | X | | X | X | |
| Living alone | X | X | | X | X | |
| Living with others | X | X | | X | X | |
| Residential Care | X | X | | X | X | |
| No fixed address (NFA) | X | X | | X | X | |
| **Smoking** | | | | | | |
| Never smoked | X | X | | X | X | |
| Ex-smoker | X | X | | X | X | |
| Current smoker | X | X | | X | X | |
| Ex-smoker, current vaper | X | X | | X | X | |
| Never smoked, current vaper | X | X | | X | X | |
| Not recorded | X | X | | X | X | |
| **Work Status** | | | | | | |
| Retired | X | X | | X | X | |
| Working | X | X | | X | X | |
| Houseperson | X | X | | X | X | |
| **Occupation** | | | | | | |
| Not Healthcare related | X | X | | X | X | |
| Healthcare worker | X | X | | X | X | |
| Laboratory worker | X | X | | X | X | |
| **Have asthma** | | | | | | |
| Yes | X | X | | X | X | |
| No | X | X | | X | X | |

| | | | | |
|---|---|---|---|---|
| **History of previous myocardial infarction** | | | | |
| Yes | X | X | X | X |
| No | X | X | X | X |
| **Currently undergoing any cancer treatment** | | | | |
| Yes | X | X | X | X |
| No | X | X | X | X |
| **Have chronic pulmonary disease** | | | | |
| Yes | X | X | X | X |
| No | X | X | X | X |

† COVID-19 COS Score of clinical symptoms is a total score of six common and important clinical symptoms, including fever, cough, fatigue, shortness of breath, diarrhoea, and body pain, each of which can be scored as 0 (no), 1 (mild), 2 (moderate), or 3 (significant). In ATOMIC2, an amended version COVID-19 COS PLUS is also designed with 2 extra clinical symptoms that also considered as having clinical importance: changes to sense of smell and loss of taste.

* The Charlson Comorbidity Index assigns a numerical value or "weight" from 1,2,3 or 6 to nineteen specific chronic illnesses. The final score (range 0-42) is simply the sum of weighted values with higher scores indicating more comorbidities.

#### 5.4 Unblinding

ATOMIC2 is an open label study without blinding.

#### 5.5 Description of Compliance with Intervention

Subjects will be randomised to receive Azithromycin 500 mg daily orally for 14 days in addition to standard care or standard care alone.

For patients allocated to AZM, the first dose will be taken within 4 hours of randomisation. Participants will be asked to take AZM at the approximately the same time every day for 14 days. The drug should be taken ideally 1 hour before a meal or 2 hours afterwards. Participants will be asked to take two 250mg capsules each day to ensure a dose of 500mg is taken.

Standard Care: symptomatic relief with rest, as-required paracetamol (where appropriate) and advice to seek further medical attention if significant worsening breathlessness. No specific therapies are yet available for COVID-19. Should additional interventions become evidence-based standard practice during the conduct of this study these would also be permitted to be provided and will be recorded under concomitant medications.

Due to its long half-life AZM accumulates over time, but to achieve a rapid effect we will use 500mg OD for 14 days, similar to the dose recommended for Lyme disease.

There will be a mortality check at day 14 and day 28 before any contact is made with participants, using hospital systems and NHS Spine or equivalent devolved nation systems. If on calling the participant at day 14 or day 28 the participant has been readmitted, data will be collected by hospital note review instead of from the participant.

Compliance will be assessed by telephone discussion with patient on day 14 of treatment with specific questioning as to the number of pills remaining. Adequate compliance will be defined as the first dose being administered within 4 hours of randomisation and at least 80% of doses i.e. a maximum of 4 / 28 tablets remaining at the end of day 14. Average number of tablets taken for patients who completed the first 14 days follow-up will be reported (Table 9).

**Table 9:** Compliance with allocated treatment AZM arm only)

| | Intervention (Azithromycin) |
|---|---|
| Started treatment (n, %) | X (X%) |
| Received 14 days of treatment (n, %) | X (X%) |
| Average numbers of tablets taken (median, IQR) | X (X, X) |

##### 5.6 Reliability

To ensure consistency, validation checks of the data will be conducted. This will include checking for duplicate records, checking the range of variable values and validating potential outliers and referring back to sites if necessary. Calculations and processes performed by a computer program, including the construction of derived data such as total scores on various outcomes, will be checked by hand calculations (where possible). This check will be conducted for a minimum of 5% of the available data or 20 participants randomly sampled from the dataset, whichever is smaller. These checks will also check any merging of different datasets. Clarification will be sought from the trial office in the case of discrepancies.

For each variable, missing value codes will be checked for consistency and the proportion of missing values per variable will be presented. Patterns of missing data will be explored. Sensitivity analyses will be conducted to explore the missing data assumptions used.

#### 6. ANALYSIS

##### 6.1 Outcome Definitions

***Primary outcomes:***

<u>All-cause hospitalisation or death</u>

Differences in proportion requiring hospital admission, from any cause, or death between the control (standard care) and the intervention (AZM) over 28 days follow-up post-randomisation will be assessed as the primary outcome. Data on hospital admission (not admitted, admitted and currently alive, and admitted and currently dead) will be collected at day 14 and at 28 days post-randomisation. Local hospital records and/or NHS Spine (or devolved nation equivalents) will be searched at local sites. If admitted between randomisation and day 28, data will be collected until hospital discharge.

***Secondary outcomes:***

<u>Progression to hospitalisation for level 2/3 ventilation or death</u>

Differences in proportion of progressing to hospitalisation for level 2/3 ventilation or death between the control (standard care) and the intervention (AZM) over 28 days follow-up post-randomisation will be assessed as the secondary outcome.

Progression to hospitalisation for level 2/3 ventilation or death is defined as differences in proportion of progressing to hospitalisation for ventilation between the control (standard care) and the intervention (AZM) over 28 days follow-up post-randomisation. Data on hospital admission (not admitted, admitted and currently alive, and admitted and currently dead) with respiratory failure requiring level 2 ventilation (NIV/CPAP/ nasal high flow) or level 3 invasive mechanical ventilation will be collected at day 14 and at 28 days post-randomisation. Local hospital records and/or NHS Spine (or devolved nation equivalents) will be searched at local sites. If admitted between randomisation and day 28, data will be collected until hospital discharge.

<u>All-cause hospitalisation or death in participants with a PCR-confirmed diagnosis of COVID-19</u>

Differences in proportion requiring hospital admission, from any cause, or death between the control (standard care) and the intervention (AZM) over 28 days follow-up post-randomisation for those who had a positive COVID-19 oropharyngeal swab test, swabs taken at time of randomisation maybe analysed later in the trial, will be assessed as the secondary outcome.

Progression to hospitalisation for ventilation or death in participants with a PCR-confirmed diagnosis of COVID-19

Differences in proportion of progressing to hospitalisation with respiratory failure requiring level 2 ventilation (NIV/CPAP/ nasal high flow) or level 3 invasive mechanical ventilation or death between the control (standard care) and the intervention (AZM) over 28 days follow-up post-randomisation for those who had a positive COVID-19 oropharyngeal swab test, swabs taken at time of randomisation maybe analysed later in the trial, will be assessed as the secondary outcome.

All-cause mortality

All-cause mortality will be assessed as a secondary outcome and will be reported based on data ascertained at 28 days after randomisation in each treatment group, and where possible, hospitalised patients will be followed up until discharge or death. Data on vital status (alive / dead, with date and presumed cause of death if appropriate) will be collected at day 14 and at 28 days post-randomisation. Local hospital records and/or NHS Spine (or devolved nation equivalents) will be searched at local sites. Death is generally expected to happen after hospitalisation.

Pneumonia

Progression to pneumonia as diagnosed by chest x-ray (or CT thorax), with compatible clinical findings, if no pneumonia is present at time of enrolment. Pneumonia will be diagnosed by a medically qualified doctor and data obtained from presenting consolidation finding on a chest x-ray. Data is assessed retrospectively at 28 days post enrolment.

Severe pneumonia

Evolution of pneumonia, as diagnosed by chest x-ray or CT thorax, if pneumonia is present at time of enrolment. To be diagnosed by a medically qualified doctor and data obtained from review of case-notes and relevant radiology. Data is assessed retrospectively at 28 days post enrolment. Severe pneumonia is defined as BTS CURB-65[12] score of 3-5. The CURB-65 is the severity assessment of community acquired pneumonia (CAP) in patients seen in hospital recommended in The British Thoracic Society (BTS) guidelines. CURB65 severity score ranges from 0-5 points based on 1 point for each feature present:

- **C**onfusion
- **U**rea > 7mmol/l
- **R**espiratory rate ≥ 30/min
- **B**lood pressure (SBP < 90 or DBP ≤ 60mmHg)
- Age ≥ **65** years

A score of 0–1 indicates a Low severity (risk of death <3%), a score of 2 indicates a Moderate severity (risk of death 9%), and a score of 3-5 indicates a High severity (risk of death 15% - 40%).

COVID-19 COS

Presence or absence of COVID-19 symptoms using COVID-19 COS scales of clinical symptoms[13]: a total score will be calculated based on the sum of the six common and important clinical symptoms:

- Shortness of Breath
- Fever (Temperature ⩾ 37.8° C, oral/rectal or tympanic)
- New persistent cough
- Diarrhoea
- Body pain
- Fatigue

Each symptom will be scored on a scale from 0-3 (0=no, 1=mild, 2=moderate, 3=significant). The total COVID-19 COS scale is ranged from 0-18, where higher the score worse the symptoms. The COVID-19 COS score will be reported based on the available data.

COVID-19 COS PLUS

The COVID-19 COS PLUS is the amended version of COVID-19 COS scale which includes 2 additional clinical symptoms that also considered as having clinical importance:

- Changes to sense of smell
- Loss of taste

The same scoring manual as the COVID-19 COS scale is used. The total COVID-19 COS PLUS scale ranges from 0-24, where higher scores indicate worse the symptoms. The COVID-19 COS PLUS score will be reported based on the available data.

*Note: these have recently been added as symptoms to monitor and which prompt people to self-isolate.*

The Severity scale score

The Severity scale score should be given based on clinical condition. If a patient is hospitalised for reasons of isolation or quarantine, they should not automatically be given the score of 3. The clinician will ascribe the score which is relevant to their clinical status, which maybe for example ambulatory score 1 even if they are hospitalised. The highest score obtained during the 28-day study period will be used in the final analysis, based on data obtained at 14 and 28 days. The maximum (peak) severity scale score will be reported based on the available data.

**Table 10:** The Severity scale scoring manual

| Descriptor | Score |
|---|---|
| Ambulatory. No limitation of activities | 0 |
| Limitation of simple activities | 1 |
| Hospitalised, mild disease, no oxygen therapy | 2 |
| Hospitalised, oxygen by conventional delivery system [a] ≤40% mask or nasal prongs | 3 |
| Hospitalised, oxygen by conventional delivery system [a] >40% mask | 4 |
| Hospitalised receiving non-invasive ventilation or receiving high-flow oxygen therapy (HFOT, >15 L/min), or continuous positive airway pressure (CPAP) [b] | 5 |
| Intubation and mechanical ventilation [c] | 6 |
| Ventilation + additional organ support | 7 |
| Death | 8 |

*Note: [a] Criteria filled if oxygen is required to maintain saturations >92% or above normal baseline for patients who use home oxygen. [b] Any patient requiring any form of PEEP delivery, or those in whom such devices are not tolerated requiring >50% oxygen with a RR>25, or rising $CO_2$ in the absence of known lung disease. [c] In patients unsuitable for ventilation, criterion is met when requiring >50% oxygen with a RR>25, or rising $CO_2$.*

***Exploratory outcomes of Interest:***

*Mechanistic analysis of blood and nasal biomarkers if available. The following samples may be taken: Blood for serum, Tempus tube (whole blood RNA), EDTA tubes (PBMC), nasal brush to be placed immediately into RNA lysis buffer (for subsequent PCR and transcriptomic analysis).*

RNA will be extracted from blood and nasal brushes, cDNA libraries prepared and subjected to RNA sequencing. Up to 100 blood and 100 nasal samples obtained at baseline, and all samples obtained at follow up will be analysed. Reads will be aligned to reference genome sequences and differential gene expression assessed between groups and their normalised expression values using a false discovery rate of $P<0.05$ and minimum log(2) fold change of ±1, with contrasts between baseline and follow-up samples; between those receiving azithromycin and those not (at follow-up); in baseline samples between those subsequently admitted and those not subsequently admitted; and between PCR-positive individuals at baseline and a comparator group of healthy control samples obtained under separate ethics. Lists of differentially expressed genes will be compared using pathway analysis and geneset enrichment analysis.

#### 6.2 Analysis Methods

***Primary outcomes***

The primary outcome for this study is the proportion of patients progressing to death or hospitalisation from any cause, by day 28 post-randomisation. The number and proportion of participants hospitalised or died by day 28 and the details of participants that been hospitalised will be summarised by treatment arm (Tables 11-12). Time to hospitalisation and the length of hospital inpatient days for this subset of patients will be presented in Figure 3. The difference in proportions between the treatment arms will be assessed using a chi-squared test and a 5% (2-sided) significance level. Relative and absolute differences in proportions together with the 95% confidence intervals will be reported. Adjusted analyses will be undertaken using logistic regression with progression as the binary outcome, adjusting for stratification factors (centre, hypertension, diabetes and sex) and other important prognostic variables (age, chronic pulmonary disease, and presence of cancer which will be determined by the presence of any of the following four items: malignancy, leukaemia, lymphoma and metastatic solid tumour in the Charlson comorbidity index) will be undertaken as a supporting analysis if sufficient events are observed to allow for including additional covariates. Supporting analyses using time-to-event methodology will also be undertaken using Kaplan Meier plots (Figure 4), log rank tests and Cox proportional hazards regression methods to explore whether the intervention delays progression to hospitalisation or death. Unadjusted and adjusted relative (relative risk, odds ratio or hazard ratios as appropriate) and absolute differences in proportions together with the 95% confidence intervals will be reported (Table 13).

The primary outcome will be analysed on the Intention to treat (ITT) population and repeated on the ITT+ve population if there is information available on the results of baseline swab tests. Intention-to-treat with positive baseline COVID-19 swab test (ITT+ve) population is the subset of the ITT population who had a positive COVID-19 test determined based on baseline swabs. The same analysis methods as described above will be undertaken on both populations.

**Table 11:** Summary of primary outcome population by intervention groups

| | **Azithromycin (N=XX)** | | **Standard care (N=XX)** | | **Total (N=XX)** | |
|---|---|---|---|---|---|---|
| | N | % | n | % | n | % |
| **Total hospitalised** | X | X | X | X | X | X |
| Received level 2 ventilation | X | X | X | X | X | X |
| Received level 3 ventilation | X | X | X | X | X | X |

| | | | | | | |
|---|---|---|---|---|---|---|
| Not receive level 2 or level 3 ventilation | X | X | X | X | X | X |
| **Total died** | X | X | X | X | X | X |

**Table 12:** Details of participants that been hospitalised – split by treatment group

| | **Azithromycin (N=XX)** | | | **Standard care (N=XX)** | | | **Total (N=XX)** | | |
|---|---|---|---|---|---|---|---|---|---|
| | Mean (SD) | Median (IQR) | Range | Mean (SD) | Median (IQR) | Range | Mean (SD) | Median (IQR) | range |
| **Age** | X (X) | X (X,X) | (X,X) | X (X) | X (X,X) | (X,X) | X (X) | X (X,X) | (X,X) |
| **COVID COS Score at first admission** | X (X) | X (X,X) | (X,X) | X (X) | X (X,X) | (X,X) | X (X) | X (X,X) | (X,X) |
| **COVID COS Score at last discharge** | X (X) | X (X,X) | (X,X) | X (X) | X (X,X) | (X,X) | X (X) | X (X,X) | (X,X) |
| **COVID COS PLUS Score at first admission** | X (X) | X (X,X) | (X,X) | X (X) | X (X,X) | (X,X) | X (X) | X (X,X) | (X,X) |
| **COVID COS PLUS Score at last discharge** | X (X) | X (X,X) | (X,X) | X (X) | X (X,X) | (X,X) | X (X) | X (X,X) | (X,X) |
| **Severity scale score at first admission** | X (X) | X (X,X) | (X,X) | X (X) | X (X,X) | (X,X) | X (X) | X (X,X) | (X,X) |
| **Severity scale score at last discharge** | X (X) | X (X,X) | (X,X) | X (X) | X (X,X) | (X,X) | X (X) | X (X,X) | (X,X) |
| **Total number of inpatient days** | X (X) | X (X,X) | (X,X) | X (X) | X (X,X) | (X,X) | X (X) | X (X,X) | (X,X) |

Figure 3. Scatter plot of time to hospitalisation and the length of hospital inpatient days

Figure 4. Kaplan Meier plots of time to primary events by treatment groups

**Table 13:** Primary outcome by intervention groups

| | **Azithromycin (n, %)** | **Standard care (n, %)** | **Unadjusted[1]** | | **Unadjusted[2]** | **Adjusted[3]** | | |
|---|---|---|---|---|---|---|---|---|
| | | | **OR (95% CI), p-val** | **RD (95% CI), p-val** | **Z (95% CI), p-val** | **OR (95% CI), p-val[3]** | **RD (95% CI), p-val[4]** | **HR (95% CI), p-val[5]** |
| **ITT** | | | | | | | | |
| **ITT +ve** | | | | | | | | |

[1] *Comparisons made using a chi-squared model*
[2] *Comparisons made using a log-rank test*
[3] *Adjusted for stratification factors (centre, hypertension, diabetes and sex) and other important prognostic variables, with control group as the reference group.*
[4] *Logistic regression model*
[5] *Cox's proportional hazards model*

##### *Secondary outcomes*

Binary outcomes (hospitalisation for level 2/3 ventilation or death (ITT & ITT+ve), all-cause mortality (ITT & ITT+ve), pneumonia & severe pneumonia (ITT & ITT+ve) will be assessed using a chi-squared test and a 5% (2-sided) significance level. Relative and absolute differences in proportions together with the 95% confidence intervals will be reported. For very low numbers of events (< 5 participants in either arm), Fisher's exact test will be used instead of chi-square test. Peak severity of illness (ITT & ITT+ve) will be considered a categorical variable and assessed using ordinal logistic regression analysis. The level of oxygen support received at each inpatient day will be presented for each participant (Figure 5).

**Table 14:** Secondary outcome (hospitalisation for level 2/3 ventilation or death) by intervention groups

| | **Azithromycin (n, %)** | **Standard care (n, %)** | **Unadjusted[1]** | |
|---|---|---|---|---|
| | | | **OR (95% CI), p-val** | **RD (95% CI), p-val** |
| **ITT** | | | | |
| **ITT +ve** | | | | |

[1] *Comparisons made using a chi-squared test or Fisher's exact test*

Figure 5. Scatter plot of level of oxygen support received by inpatient day for each participant

*All- cause mortality* will be reported as ascertained based on data up to 28 days after randomisation in each treatment group, and where possible, hospitalised patients will be followed up until discharge or death. If there are any deaths, the treatment groups will be compared for all-cause mortality using unadjusted Fisher's Exact test. All-cause mortality will be analysed on Intention to treat (ITT) and the Intention-to-treat with positive baseline COVID-19 swab test (ITT+ve) population.

**Table 15:** Results of all-cause mortality by intervention groups

| | Azithromycin (n, %) | Standard care (n, %) | Unadjusted[1] | |
|---|---|---|---|---|
| | | | OR (95% CI), p-val | RD (95% CI), p-val |
| **ITT** | | | | |
| **ITT +ve** | | | | |

[1] *Comparisons made using a chi-squared test or Fisher's exact test*

*The proportion of participants progressing to pneumonia* by day 28 post-randomisation will be analysed within the subset of participants for whom no pneumonia is present at the time of enrolment. The number and proportion of these participants who have been diagnosed with pneumonia during follow-up will be summarised by treatment group. The difference in proportions between the treatment arms will be assessed using a chi-squared test. For very low numbers of events, Fisher's exact test will be used instead of chi-square test.

**Table 16:** Results of progressing to pneumonia by intervention groups

| | Azithromycin (n, %) | Standard care (n, %) | Unadjusted[1] | |
|---|---|---|---|---|
| | | | OR (95% CI), p-val | RD (95% CI), p-val |
| **ITT** | | | | |
| **ITT +ve** | | | | |

[1] *Comparisons made using a chi-squared test or Fisher's exact test*

*The proportion of participants progressing to severe pneumonia* by day 28 post-randomisation will be analysed within the subset of participants for whom pneumonia is present at time of enrolment. A summary of the number and proportion of participants who have been diagnosed with pneumonia at baseline and/or during follow-up will be summarised by treatment groups (Table 17). Severe pneumonia is defined as BTS CURB-65 score of 3-5 (defined in section 6.1). The difference in proportions between the treatment arms will be assessed using a chi-squared test (Table 18). For very low numbers of events, Fisher's exact test will be used instead of chi-square test.

**Table 17:** Summary of diagnosis with pneumonia from baseline until end of follow-up by intervention groups

| | Azithromycin (N=XX) | | Standard care (N=XX) | | Total (N=XX) | |
|---|---|---|---|---|---|---|
| | n | % | n | % | n | % |
| **ITT** | | | | | | |
| **No Pneumonia at baseline** | **X** | **X** | **X** | **X** | **X** | **X** |
| None throughout | X | X | X | X | X | X |
| None to pneumonia | X | X | X | X | X | X |
| None to severe pneumonia | X | X | X | X | X | X |
| **Pneumonia at baseline** | **X** | **X** | **X** | **X** | **X** | **X** |
| Pneumonia to none | X | X | X | X | X | X |

| | | | | | | |
|---|---|---|---|---|---|---|
| Pneumonia throughout | X | X | X | X | X | X |
| Pneumonia to severe pneumonia | X | X | X | X | X | X |
| **Not available at baseline** | **X** | **X** | **X** | **X** | **X** | **X** |
| **ITT +ve** | | | | | | |
| **No Pneumonia at baseline** | **X** | **X** | **X** | **X** | **X** | **X** |
| None throughout | X | X | X | X | X | X |
| None to pneumonia | X | X | X | X | X | X |
| None to severe pneumonia | X | X | X | X | X | X |
| **Pneumonia at baseline** | **X** | **X** | **X** | **X** | **X** | **X** |
| Pneumonia to none | X | X | X | X | X | X |
| Pneumonia throughout | X | X | X | X | X | X |
| Pneumonia to severe pneumonia | X | X | X | X | X | X |
| **Not available at baseline** | **X** | **X** | **X** | **X** | **X** | **X** |

**Table 18:** Results of progressing to severe pneumonia by intervention groups

| | Azithromycin (n, %) | Standard care (n, %) | Unadjusted[1] | |
|---|---|---|---|---|
| | | | OR (95% CI), p-val | RD (95% CI), p-val |
| **ITT** | | | | |
| **ITT +ve** | | | | |

[1] *Comparisons made using a chi-squared test or Fisher's exact test*

*<u>The differences in peak severity of illness</u>* up to day 28 post-randomisation will be analysed. The severity score during the entire study period will be summarised by treatment groups among the ITT population and all hospitalised patients (Table 19). Box plots will be used to compare the severity score at Baseline, Day 14 and Day 28 of the ITT population (Figure 6). The severity scale score at each inpatient day will be presented by patients (Figure 7). Peak severity score post-baseline will be summarised by treatment groups (Table 20). The difference between the treatment arms will be assessed using ordinal logistic regression. Adjusted analyses will also be undertaken with stratification factors (centre, hypertension, diabetes and sex) and other important prognostic variables being adjusted (Table 21). A bar chart will be used to compare the baseline severity score and the peak severity score of all hospitalised patients with the ITT population (Figure 8). The time to the peak severity will be summarised (Table 22).

**Table 19:** Description of the severity scale score during the entire study – ITT & all hospitalised patients

| Time points | Azithromycin (N=XX)<br>mean (SD), median (IQR), range | Standard care (N=XX)<br>mean (SD), median (IQR), range | Total (N=XX)<br>mean (SD), median (IQR), range |
|---|---|---|---|
| **ITT** | | | |
| Baseline | X (X), X (X, X), (X, X) | X (X), X (X, X), (X, X) | X (X), X (X, X), (X, X) |
| Day 14 | X (X), X (X, X), (X, X) | X (X), X (X, X), (X, X) | X (X), X (X, X), (X, X) |
| Day 28 | X (X), X (X, X), (X, X) | X (X), X (X, X), (X, X) | X (X), X (X, X), (X, X) |
| **All hospitalised patients** | | | |
| Baseline | X (X), X (X, X), (X, X) | X (X), X (X, X), (X, X) | X (X), X (X, X), (X, X) |
| Day 14 | X (X), X (X, X), (X, X) | X (X), X (X, X), (X, X) | X (X), X (X, X), (X, X) |
| Day 28 | X (X), X (X, X), (X, X) | X (X), X (X, X), (X, X) | X (X), X (X, X), (X, X) |

Figure 6. Box plots of the severity scale score during the entire study – by intervention groups (ITT population)

Figure 7. Scatter plot of the severity scale score at each hospital inpatient days for each patient – by intervention groups (ITT population)

**Table 20:** Description of peak severity scale score during follow-up – ITT & all hospitalised patients

| | Azithromycin (N=XX) | Standard care (N=XX) | Total (N=XX) |
|---|---|---|---|
| | mean (SD), median (IQR), range | mean (SD), median (IQR), range | mean (SD), median (IQR), range |
| **ITT** | X (X), X (X, X), (X, X) | X (X), X (X, X), (X, X) | X (X), X (X, X), (X, X) |
| **All hospitalised patients** | X (X), X (X, X), (X, X) | X (X), X (X, X), (X, X) | X (X), X (X, X), (X, X) |

*Note: Peak severity scale score will be assessed post-baseline.*

Figure 8. Bar chart of the peak severity scale score during follow-up compared to baseline – by intervention groups (ITT & all hospitalised patients)

**Table 21:** Results of difference in peak severity scale score by intervention groups (ITT & ITT+ve)

| | Azithromycin (n, %) | Standard care (n, %) | Unadjusted[1] | Adjusted[2] |
|---|---|---|---|---|
| | | | OR (95% CI), p-val | OR (95% CI), p-val |
| **ITT** | | | | |
| **ITT +ve** | | | | |

[1] *Comparisons made using an ordinal logistic regression model*
[2] *Adjusted for stratification factors (centre, hypertension, diabetes and sex) and other important prognostic variables, with control group as the reference group.*

**Table 22:** Time to peak and the time points of peak severity scale score during follow-up – ITT & all hospitalised patients

| | Azithromycin (N=XX) | | Standard care (N=XX) | | Total (N=XX) | |
|---|---|---|---|---|---|---|
| **Time to peak severity (Days)** | median (IQR), range | | median (IQR), range | | median (IQR), range | |
| ITT | X (X, X), (X, X) | | X (X, X), (X, X) | | X (X, X), (X, X) | |
| All hospitalised patients | X (X, X), (X, X) | | X (X, X), (X, X) | | X (X, X), (X, X) | |
| | n | % | n | % | n | % |
| **Time points of peak severity (ITT)** | | | | | | |
| Day 14 | X | X | X | X | X | X |
| Day 28 | X | X | X | X | X | X |
| **Time points of peak severity (All hospitalised patients)** | | | | | | |
| Day 14 | X | X | X | X | X | X |
| Day 28 | X | X | X | X | X | X |
| Hospital admission | X | X | X | X | X | X |
| Hospital inpatient days | X | X | X | X | X | X |
| Hospital discharge | X | X | X | X | X | X |

*Note: Peak severity scale score will be assessed post-baseline.*

The COVID COS score and COVID COS PLUS score at Baseline, Day 14 and Day 28 will be summarised by treatment groups (Table 23). A bar chart will be used to compare the Baseline, Day 14 and Day 28 COVID COS PLUS scores of all hospitalised patients with the ITT population (Figure 9). The COVID COS PLUS score at each inpatient day will be presented by patients (Figure 10).

**Table 23:** Description of the COVID COS score and COVID COS PLUS score during the entire study – ITT & all hospitalised patients

| | Azithromycin (N=XX) | Standard care (N=XX) | Total (N=XX) |
|---|---|---|---|
| | mean (SD), median (IQR), range | mean (SD), median (IQR), range | mean (SD), median (IQR), range |
| **COVID COS score** | | | |
| **ITT** | | | |
| Baseline | X (X), X (X, X), (X, X) | X (X), X (X, X), (X, X) | X (X), X (X, X), (X, X) |
| Day 14 | X (X), X (X, X), (X, X) | X (X), X (X, X), (X, X) | X (X), X (X, X), (X, X) |
| Day 28 | X (X), X (X, X), (X, X) | X (X), X (X, X), (X, X) | X (X), X (X, X), (X, X) |
| **All hospitalised patients** | | | |

| | | | |
|---|---|---|---|
| Baseline | X (X), X (X, X), (X, X) | X (X), X (X, X), (X, X) | X (X), X (X, X), (X, X) |
| Day 14 | X (X), X (X, X), (X, X) | X (X), X (X, X), (X, X) | X (X), X (X, X), (X, X) |
| Day 28 | X (X), X (X, X), (X, X) | X (X), X (X, X), (X, X) | X (X), X (X, X), (X, X) |
| **COVID COS PLUS score** | | | |
| **ITT** | | | |
| Baseline | X (X), X (X, X), (X, X) | X (X), X (X, X), (X, X) | X (X), X (X, X), (X, X) |
| Day 14 | X (X), X (X, X), (X, X) | X (X), X (X, X), (X, X) | X (X), X (X, X), (X, X) |
| Day 28 | X (X), X (X, X), (X, X) | X (X), X (X, X), (X, X) | X (X), X (X, X), (X, X) |
| **All hospitalised patients** | | | |
| Baseline | X (X), X (X, X), (X, X) | X (X), X (X, X), (X, X) | X (X), X (X, X), (X, X) |
| Day 14 | X (X), X (X, X), (X, X) | X (X), X (X, X), (X, X) | X (X), X (X, X), (X, X) |
| Day 28 | X (X), X (X, X), (X, X) | X (X), X (X, X), (X, X) | X (X), X (X, X), (X, X) |

Figure 9. Bar chart of the COVID COS PLUS score during the entire study – by intervention groups (ITT population & primary outcome population)

Figure 10. Scatter plot of the COVID COS PLUS score at each hospital inpatient day for each participant – by intervention groups (ITT population)

##### 6.3 Complications experienced during hospitalisation

The number of participants experiencing complications during hospitalisation and the total number of complications during hospitalisation will be summarised by treatment group (Tables 24-25).

**Table 24:** Summary of complications experienced during hospitalisation - by treatment group

| | Azithromycin | | Standard care | |
|---|---|---|---|---|
| | **n** | **%** | **n** | **%** |
| **Total number of participants that had complication(s)** | | | | |
| Had a complication | X | X | X | X |
| Had more than one complication | X | X | X | X |
| **Total number of participants that did not have a complication** | X | X | X | X |

**Table 25:** Details of complications experienced during hospitalisation summarised by treatment group

| | Azithromycin | | Standard care | |
|---|---|---|---|---|
| | **n** | **%** | **n** | **%** |
| **Respiratory** | | | | |
| ARDS | X | X | X | X |
| Pneumothorax | X | X | X | X |
| Pleural effusion | X | X | X | X |
| Cryptogenic organising pneumonia (COP) | X | X | X | X |
| Bronchiolitis | X | X | X | X |
| **Neurological** | | | | |
| Meningitis / Encephalitis | X | X | X | X |
| Seizure | X | X | X | X |
| Stroke / cerebrovascular accident | X | X | X | X |
| **Cardiovascular** | | | | |
| New Congestive heart failure | X | X | X | X |
| Endocarditis / myocarditis / pericarditis | X | X | X | X |
| New cardiac arrhythmia | X | X | X | X |
| Cardiac ischaemia (a clinical diagnosis, not to be based on a raised troponin alone as this is likely suggestive of myocarditis | X | X | X | X |
| Cardiac arrest | X | X | X | X |

| | | | | |
|---|---|---|---|---|
| **Other** | | | | |
| Bacteraemia | X | X | X | X |
| Coagulation disorder /disseminated intravascular coagulation | X | X | X | X |
| Anaemia | X | X | X | X |
| Rhabdomyolysis / myositis | X | X | X | X |
| Acute renal injury / acute renal failure | X | X | X | X |
| Gastrointestinal haemorrhage | X | X | X | X |
| Pancreatitis | X | X | X | X |
| Liver dysfunction | X | X | X | X |
| Hyperglycaemia | X | X | X | X |
| Hypoglycaemia | X | X | X | X |
| Total number of complications during hospitalisation | X | - | X | - |

##### 6.4 Missing Data

Missing data will be minimised by careful data management. Missing data will be described with reasons given where available; the number and percentage of individuals in the missing category will be presented by treatment arm.

##### 6.5 Sensitivity Analysis

Sensitivity analysis will be carried out on a per-protocol basis. The following are the definitions for the per-protocol population in this study:

1. If on the active treatment arm, participants should have adequate compliance defined as the first dose being administered within 4 hours of randomisation and at least 80% of doses i.e. a maximum of 4 / 28 tablets remaining at the end of day 14.

2. If on the standard treatment arm, participants should not receive azithromycin or other macrolide antibiotic during the period between randomisation and day 14 or treatment or hospitalisation, whichever occurs first. (This is important as we specifically excluded macrolide antibiotics in conmed, and we're aware some received clarithromycin in standard care arm, which may have similar properties to AZM; however many patients admitted to hospital may have then been prescribed a macrolide antibiotic and as the primary event will have occurred by that point we would not want to exclude them).

3. Not found to be ineligible after randomisation will be exclude from the per-protocol population.

##### 6.6 Pre-specified Subgroup Analysis

Forest plots will be presented to explore the consistency of results for the pre-specified stratification factors: hypertension, diabetes and sex and age will be presented using treatment by variable interaction tests[15].

##### 6.7 Harms

All SAEs (other than those defined as foreseeable below) occurring within the first 14 days of the IMP administration will be reported.

It is important to consider the natural history of COVID-19 with regards to this study, the expected sequelae of the illness, and the relevance of these complications to the trial treatment. All eligible participants have a potential poor prognosis, and due to the complexity of their condition are at increased risk of experiencing multiple adverse events. Additionally, Azithromycin has a very well safety profile. Therefore taking a risk adapted response, the labelling of a Serious Adverse Event (SAE) will be limited to serious events, which might reasonably occur as a consequence of the trial treatment (i.e. not events that are part of the natural history of the primary disease process, such as hospitalisation or death). SAEs must be reported in the participant's medical notes, on the trial CRF, and reported to the CTU using the SAE Reporting Form, within 24 hours of the site observing or learning of the SAE(s). All sections of the SAE Reporting Form must be completed.

Deaths due to COVID-19 disease under study are exempt from reporting as SAEs as they will be captured as part of the primary outcome. An AE should not be recorded for the positive SAR- CoV-19 infection, this will be known at time of inclusion into the study and should be recorded as medical history. Worsening of COVID-19 symptoms is captured as an efficacy measure and in general will not be considered an adverse event.

The number of people in each arm reporting adverse events will be summarised. The total number of SAES per intervention arm, the number of participants with SAEs and the number of SAES per participant will be reported. Any SUSARs and SARS will also be reported and discussed. Serious Adverse Events (SAEs) are also listed here. SAEs from previous reports will be greyed out and new information of existing SAEs or new SAEs are in standard black font. The definitions of adverse event are listed in Appendix B.

**Table 26:** Summary of serious adverse events including SARs/SUSARs

| | Intervention | Control | Total |
|---|---|---|---|
| Number of SAEs, SARs and SUSARs | X | X | X |
| SAEs | X | X | X |
| SARs | X | X | X |
| SUSARs | X | X | X |
| Number of participants with SAEs, SARs and SUSARs | X (%) | X (%) | X (%) |
| Average number of SAEs, SARs and SUSARs per participant (if ≥1) | X | X | X |
| Number of participants without SAE, SAR or SUSAR | X (%) | X (%) | X (%) |
| Number of deaths that not due to COVID-19 | X (%) | X (%) | X (%) |

#### 7. VALIDATION OF THE PRIMARY ANALYSIS

To validate the primary outcome and key secondary outcomes (i.e. hospitalisation for level 2/3 ventilation or death, and the differences in peak severity of illness), a statistician not involved in the trial will independently repeat the analyses detailed in this SAP, by using different statistical software (if possible). The results will be compared and any discrepancies will be reported in the Statistical report (See OCTRU SOP STATS-005 Statistical Report). This will include derivation of the primary and key secondary outcomes from raw data.

#### 8. SPECIFICATION OF STATISTICAL PACKAGES

All analysis will be carried out using appropriate validated statistical software such as STATA, SAS, SPLUS or R. The relevant package and version number will be recorded in the statistical programs, the Statistical report and primary publications.

#### APPENDIX A: GLOSSARY OF ABBREVIATIONS

| | |
|---|---|
| AE | Adverse event |

| | |
|---|---|
| ALT | Alanine aminotransferase |
| APTT | Activated partial thromboplastin time |
| AR | Adverse reaction |
| AST | Aspartate aminotransferase |
| AZM | Azithromycin |
| BP | Blood pressure |
| BTS | British Thoracic Society |
| CCL | Chemokine (C-C motif) ligand |
| CF | Cystic Fibrosis |
| CI | Chief Investigator |
| COPD | Chronic Obstructive Pulmonary Disease |
| CPAP | Continuous positive airway pressure (ventilation) |
| CRA | Clinical Research Associate (Monitor) |
| CRF | Case Report Form |
| CRP | C-reactive protein |
| CT | Computed tomogram |
| CTA | Clinical Trials Authorisation |
| CTRG | Clinical Trials and Research Governance |
| CURB-65 | Confusion, Urea >7.0 mmol/L, Respiratory Rate >/=30 breaths/min, Blood pressure <90 systolic or </=60 diastolic, Age >/= 65 years. |
| CXR | Chest X-Ray |
| DMSC | Data Monitoring Committee / Data Monitoring and Safety Committee |
| DPB | Diffuse Pan Bronchiolitis |
| DSUR | Development Safety Update Report |
| EDTA | Ethylenediaminetetraacetic acid |
| ePR | Electronic patient record |
| FBC | Full blood count |
| GCP | Good Clinical Practice |
| GCSF | Granulocyte Colony Stimulating Factor |
| GP | General Practitioner |
| HFOT | High-Flow Oxygen Therapy: warmed, humidified oxygen delivered via a nasal mask at >15 L/min |
| HR | Heart Rate |
| HRA | Health Research Authority |

| | |
|---|---|
| ICF | Informed Consent Form |
| ICH | International Conference on Harmonisation |
| IL | Interleukin |
| IMP | Investigational Medicinal Product |
| IMV | Invasive mechanical ventilation (ventilatory support delivered via an endotracheal tube) |
| ISARIC | International Severe Acute Respiratory and Emerging Infection Consortium |
| ISG | Interferon Stimulated Gene |
| MHRA | Medicines and Healthcare products Regulatory Agency |
| MERS | Middle East Respiratory Syndrome |
| MxA | A membrane protein |
| NIV | Non-invasive ventilation (ventilatory support via an external face mask in a non-sedated person) |
| NHS | National Health Service |
| PCR | Polymerase chain reaction |
| PEEP | Positive End-Expiratory Pressure |
| PI | Principal Investigator |
| PIL | Participant/ Patient Information Leaflet |
| PROBE | Prospective randomized open blinded end-point (PROBE) clinical trial |
| PT | Prothrombin time |
| QTc | Corrected QT interval |
| R&D | NHS Trust R&D Department |
| RCT | Randomised Controlled Trial |
| REC | Research Ethics Committee |
| RES | Research Ethics Service |
| RNA | Ribonucleic acid |
| RR | Respiratory Rate |
| RSI | Reference Safety Information |
| RV | RhinoVirus |
| SAE | Serious Adverse Event |
| SAR | Serious Adverse Reaction |
| SARS | Severe Acute Respiratory Syndrome |
| SDV | Source Data Verification |
| SMPC | Summary of Medicinal Product Characteristics |

| | |
|---|---|
| SOP | Standard Operating Procedure |
| SSRI | Selective Serotonin Reuptake Inhibitor |
| SST | Serum Separating Tube |
| SUSAR | Suspected Unexpected Serious Adverse Reactions |
| Tempus tube | Blood collection tubes for RNA purification |
| TMF | Trial Master File |
| U+E | Urea and electrolytes |

#### APPENDIX B: ADVERSE EVENT DEFINITIONS

| | |
|---|---|
| Adverse Event (AE) | Any untoward medical occurrence in a participant to whom a medicinal product has been administered, including occurrences which are not necessarily caused by or related to that product. |
| Adverse Reaction (AR) | An untoward and unintended response in a participant to an investigational medicinal product which is related to any dose administered to that participant.<br>The phrase "response to an investigational medicinal product" means that a causal relationship between a trial medication and an AE is at least a reasonable possibility, i.e. the relationship cannot be ruled out.<br>All cases judged by either the reporting medically qualified professional or the Sponsor as having a reasonable suspected causal relationship to the trial medication qualify as adverse reactions. |
| Serious Adverse Event (SAE) | A serious adverse event is any untoward medical occurrence that:<br>• results in death<br>• is life-threatening<br>• requires inpatient hospitalisation or prolongation of existing hospitalisation<br>• results in persistent or significant disability/incapacity<br>• consists of a congenital anomaly or birth defect*.<br>Other 'important medical events' may also be considered a serious adverse event when, based upon appropriate medical judgement, the event may jeopardise the participant and may require medical or surgical intervention to prevent one of the outcomes listed above.<br>NOTE: The term "life-threatening" in the definition of "serious" refers to an event in which the participant was at risk of death at the time of the event; it does not refer to an event which hypothetically might have caused death if it were more severe. |

| | |
|---|---|
| Serious Adverse Reaction (SAR) | An adverse event that is both serious and, in the opinion of the reporting Investigator, believed with reasonable probability to be due to one of the trial treatments, based on the information provided. |
| Suspected Unexpected Serious Adverse Reaction (SUSAR) | A serious adverse reaction, the nature and severity of which is not consistent with information in the MHRA approved Reference Safety Information |

#### APPENDIX C: SIMULATIONS FOR INTERIM ANALYSES

##### Simulation Assumptions

Simulations were carried out using R version 3.6.1 (2019-07-05). Software has been validated and is stored centrally. The R package checkpoint() is used to install R packages as they were on 10th October 2019 for future reproducibility.

The predictive probability of success (PPS) is the probability of claiming success if the trial continues and recruits to full sample size given the data observed so far. Here, success is defined as rejecting the null hypothesis of no difference with 5% significance. If the predictive probability of success is high then we continue to the end of the trial, otherwise we stop the trial due to futility. We have used a threshold of 0.1 for claiming futility, i.e. the trial is futile if PPS < 0.1.

Let the total sample size be N where N=778 with an interim analysis after $n_1$ participants have been recruited and followed up for 28 days. Let *c* denote control and *t* treatment, and assume Beta (0.3, 0.7) (i.e. based on the probability of hospitalisation that the sample size was based on) priors for both groups. Then,

$$y_c \sim Bernoulli(\theta_c)$$

$$y_t \sim Bernoulli(\theta_t)$$

$$\theta_c \sim Beta(0.3, 0.7)$$

$$\theta_t \sim Beta(0.3, 0.7)$$

$$H_0: \theta_c = \theta_t, H_1: \theta_c \neq \theta_t$$

The trial is successful if we reject the null hypothesis with a 2-sided p-value of 0.05, i.e. a Chi-squared test with p-value < 0.05.

At an interim analysis, suppose we have observed $y_c$ and $y_t$ responses in the control and treatment groups respectively, and therefore $(n_1/2) - y_c$ and $(n_1/2) - y_t$ non-responses. If the trial continues, a further $N-n_1=n_2$ participants will be recruited.

To calculate the PPS:

- Simulate from the posterior for each group
  - $\tilde{\theta}_c \sim Beta(0.3 + y_c, 0.7 + n_1/2 - y_c)$, $\tilde{\theta}_t \sim Beta(0.3 + y_t, 0.7 + n_1/2 - y_t)$
- Simulate from the predictive for each group given the draw from the posterior
  - $\tilde{y}_c \sim Binomial(n_2/2, \tilde{\theta}_c)$, $\tilde{y}_t \sim Binomial(n_2/2, \tilde{\theta}_t)$
- Carry out a chi-squared test using the combined observed and simulated data

Repeat this process 1,000 times and calculate the proportion of times the p-value is < 0.05. This is the PPS at the interim analysis given the data observed in the first part of the trial.

To calculate the properties of the trial we imagine different scenarios for the probabilities of response for treatment and control groups and assume interim analyses after 100, 250, 350, 500 and 600 patients have been recruited and followed for 28 days. Average trial properties are based on 1,000 simulations for each trial. As it is Bayesian and futility rules are based on predictive probabilities you can assess stopping rules at any point you like. Based on the simulations, futility was assessed at (100, 250, 350, 500, 600) and (100, 300, 400, 500, 600) the average sample size is same with both sets of interims, but there is still only a small chance of stopping even at 300. Therefore, a minimum of another 200, or 350 is a preferred next interim point.

The table below gives the scenarios and probability of stopping for futility at each interim analysis, and the overall probability of success. If you look at the (0.2, 0.2) scenario for instance, by the time you get to an interim at n=600, you have already stopped on average 67% (0.30+0.37) of the time, there are on average only 33% of trials that would still be continuing with these underlying true probabilities of hospitalisation. 12% of trials would fail at n=600 if they had not stopped earlier, meaning 21% of trials recruit the full 778 participants.

The accumulated probability of stopping for futility at each interim is: 0, 0, 30%, 67% and 79% and this is also shown in the figure – cumulative probability of stopping at each interim. For simulations from the null (where the treatment and control group response rates are the same), the overall probability of claiming success gives the error rate for the trial. The figure shows the cumulative probability that the trial stops early for futility at each interim, dashed lines show scenarios where there is no difference between treatment and control, and solid lines when treatment has a lower response rate than control. Also given are the power and average sample size. The average trial sizes are calculated as the probability of stopping at each interim * sample size at that interim, and is what you would get on average. However the actual trial size can only be the size of one of the interims. So in the simulations, trials were either of size 100 (not actually as none stopped here), 200 (not actually as none stopped here), 350, 500, 600 or 778. If you look at the (0.2, 0.2) scenario for instance, the average sample size is calculated as 0*100 + 0*250 + 0.3*350 + 0.37*500 + 0.12*600 + 0.21*778 = 525.38 (rounded). Non-rounded calculation is 0.304*350+0.370*500+0.123*600+0.203*778 = 523.134 (same as shows in the figure).

| | Probability of claiming futility | | | | | Overall probability of success |
|---|---|---|---|---|---|---|
| Interim analysis | $n_1$=100 | $n_2$=250 | $n_3$=350 | $n_4$=500 | $n_6$=600 | |
| $\theta_c = 0.2, \theta_{t=}0.2$ | 0 | 0 | 0.30 | 0.37 | 0.12 | 0.04 |
| $\theta_c = 0.2, \theta_{t=}0.14$ | 0 | 0 | 0.10 | 0.12 | 0.07 | 0.53 |
| $\theta_c = 0.25, \theta_{t=}0.25$ | 0 | 0 | 0.30 | 0.36 | 0.16 | 0.04 |
| $\theta_c = 0.25, \theta_{t=}018$ | 0 | 0 | 0.09 | 0.08 | 0.05 | 0.61 |
| $\theta_c = 0.3, \theta_{t=}0.3$ | 0 | 0 | 0.29 | 0.36 | 0.15 | 0.04 |
| $\theta_c = 0.3, \theta_{t=}0.2$ | 0 | 0 | 0.03 | 0.02 | 0.01 | 0.88 |
| $\theta_c = 0.4, \theta_{t=}0.4$ | 0 | 0 | 0.26 | 0.37 | 0.16 | 0.05 |
| $\theta_c = 0.4, \theta_{t=}0.28$ | 0 | 0 | 0.02 | 0.01 | 0.01 | 0.91 |

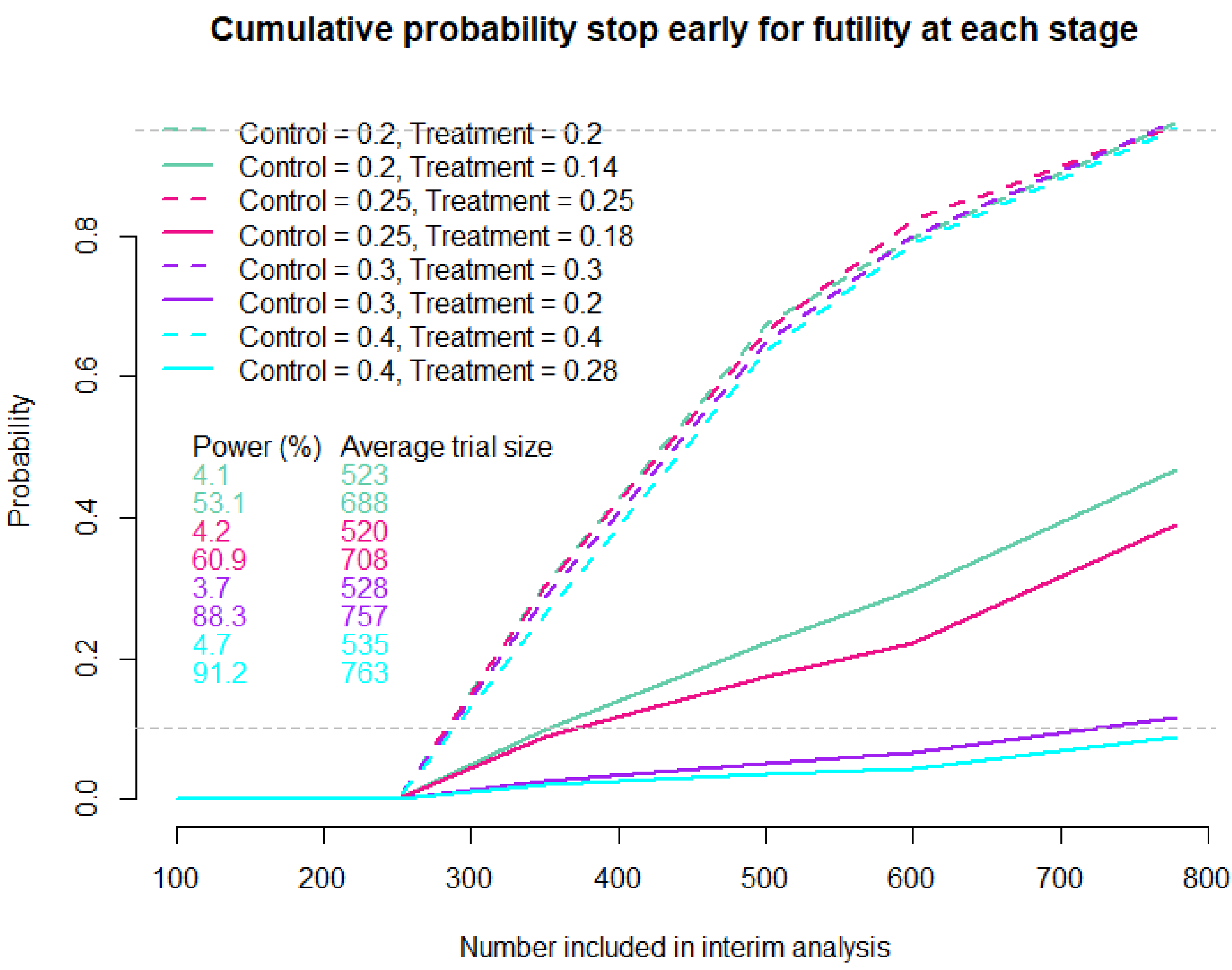
Cumulative probability stop early for futility at each stage
Control = 0.2, Treatment = 0.2
Control = 0.2, Treatment = 0.14
Control = 0.25, Treatment = 0.25
Control = 0.25, Treatment = 0.18
Control = 0.3, Treatment = 0.3
Control = 0.3, Treatment = 0.2
Control = 0.4, Treatment = 0.4
Control = 0.4, Treatment = 0.28
Power (%) Average trial size
4.1 523
53.1 688
4.2 520
60.9 708
3.7 528
88.3 757
4.7 535
91.2 763
Probability
0.0
0.2
0.4
0.6
0.8
100
200
300
400
500
600
700
800
Number included in interim analysis

### Appendix 3: Supplementary Results

**1 Study participants**

Reasons for ineligibility, loss to follow-up and exclusion from the analysis are summarised, as the number of patients who withdraw before each analysis time point. Details of reasons for exclusion are summarised in Table S1.

**Table S1: Reasons for exclusion**

| Reason for exclusion | Number excluded |
|---|---|
| **Not meeting inclusion criteria** | |
| *Not appropriate for initial ambulatory (outpatient) management* | 408 |
| *No clinical diagnosis of highly-probable or confirmed COVID-19 infection* | 39 |
| *Patient had symptoms for more than 14 days** | 81 |
| *Has medical history that might, in the opinion of the attending clinician, put the patient at significant risk if he/she were to participate in the trial* | 95 |
| *Unable to understand written English* | 61 |
| *Unable to give informed consent* | 102 |
| **Meeting exclusion criteria** | |
| *Known hypersensitivity to any Macrolide including Azithromycin, ketolide antibiotic, or the excipients including an allergy to soya or peanuts* | 15 |
| *Known fructose intolerance, glucose-galactose malabsorption, or sucrose- isomaltase-insufficiency* | 3 |
| *Currently on a Macrolide antibiotic (Clarithromycin, Azithromycin, Erythromycin, Telithromycin, Spiramycin)* | 28 |
| *Patient is on SSRI (Selective Serotonin Reuptake Inhibitor)*** | 29 |
| *Elevated cardiac troponin at initial assessment suggestive of significant myocarditis (if clinically the clinical team have felt it appropriate to check the patient's troponin levels)* | 12 |
| *Evidence of QTc prolongation: QTc>480ms* | 19 |
| *Significant electrolyte disturbance (e.g., hypokalaemia K+<3.5 mmol/L)* | 44 |
| *Clinically relevant bradycardia (P<50 bpm), non-sustained ventricular tachycardia or unstable severe cardiac insufficiency* | 8 |
| *Currently on hydroxychloroquine or chloroquine* | 7 |
| *The patient has been previously randomised into ATOMIC2* | 2 |
| **Total ineligible***** | 649 |
| **Eligible but refused** | |
| *Did not want to take part* | 39 |
| *Did not want to complete forms* | 2 |
| *Unwilling to comply with study treatment* | 19 |
| *Patient has ongoing medical issues (such as acute/chronic conditions) and feels the trial would be too much.* | 6 |
| *No reason given* | 14 |
| *Not willing to say* | 2 |
| *Unknown* | 2 |
| **Total declined** | 84 |
| **Other** | |
| *Lack of staff/Out of hours* | 120 |
| *Lack of clinician equipoise* | 0 |
| *More than 14 days symptoms/test-positive* | 3 |
| *Too well* | 3 |
| *No blood tests/ECG to rule out all of the exclusion criteria* | 1 |
| *Pregnant/Breastfeeding* | 2 |
| *No fixed abode* | 1 |
| *Drug overdose* | 1 |
| *Not suitable* | 2 |
| *Missed* | 5 |
| *Unknown* | 23 |
| **Total patients screened but not randomised** | 894 |

**This inclusion criterion was added after a protocol amendment, and so was not applied to the first 180 patients screened*

***This inclusion criterion was removed after a protocol amendment, and so was only applied to the first 180 patients screened*
****This is the total number of participants marked as ineligible. Each participant could give more than one reason for ineligibility.*

**2 Baseline Characteristics**

The baseline comparability of the two treatment groups (azithromycin and standard care) is considered. The treatment groups are reported in terms of stratification factors (see Table S2), baseline demographic characteristics (see Table S3), and baseline demographic characteristics by SWAB results (see Table S4). Details of the comorbidities participants experienced are summarised by treatment group in Table S5. The two groups were well balanced at baseline. In Table S6, baseline demographic data are presented separately for those participants who had a positive COVID swab and those that did not. Numbers (with percentages) for binary and categorical variables and the number of available observations with means (and standard deviations), medians (with inter-quartile ranges) and range for continuous variables are reported for each treatment group and overall. There were no tests of statistical significance nor confidence intervals for differences between randomised groups on any baseline variable. Comparison of the demographic characteristics of the participants in the total population with the subset of the population who are hospitalised or died (primary outcome) is reported (Table S7).

**Table S2: Stratification factors at baseline – split by treatment group and overall**

| | Azithromycin (n = 147) | | Standard Care (n = 148) | | Total (n = 295) | |
|---|---|---|---|---|---|---|
| | n | % | n | % | n | % |
| **Centre** | | | | | | |
| Basingstoke | 3 | 2% | 5 | 3.4% | 8 | 2.7% |
| Burnley Hospital | 3 | 2% | 1 | 0.7% | 4 | 1.4% |
| City Hospital | 2 | 1.4% | 2 | 1.4% | 4 | 1.4% |
| Darlington Memorial Hospital | 1 | 0.7% | 0 | 0% | 1 | 0.3% |
| Denmark Hill | 9 | 6.1% | 10 | 6.8% | 19 | 6.4% |
| Dundee | 0 | 0% | 1 | 0.7% | 1 | 0.3% |
| Horton | 7 | 4.8% | 8 | 5.4% | 15 | 5.1% |
| James Cook University Hospital | 2 | 1.4% | 1 | 0.7% | 3 | 1% |
| North Tees | 0 | 0% | 1 | 0.7% | 1 | 0.3% |
| Oxford | 12 | 8.2% | 9 | 6.1% | 21 | 7.1% |
| Princess Royal University Hospital | 6 | 4.1% | 5 | 3.4% | 11 | 3.7% |
| Royal Berkshire Hospital | 13 | 8.8% | 10 | 6.8% | 23 | 7.8% |
| Royal Blackburn Hospital | 25 | 17% | 27 | 18.2% | 52 | 17.6% |
| Royal Derby Hospital | 17 | 11.6% | 14 | 9.5% | 31 | 10.5% |
| Royal London Hospital | 3 | 2% | 4 | 2.7% | 7 | 2.4% |
| St George's Hospital | 30 | 20.4% | 31 | 20.9% | 61 | 20.7% |
| University College London Hospitals | 8 | 5.4% | 10 | 6.8% | 18 | 6.1% |
| University Hospital Llandough | 2 | 1.4% | 4 | 2.7% | 6 | 2% |
| University Hospital Wales | 4 | 2.7% | 5 | 3.4% | 9 | 3.1% |
| **Hypertension** | | | | | | |
| Yes | 25 | 17.0% | 27 | 18.2% | 52 | 17.6% |
| No | 122 | 83.0% | 121 | 81.8% | 243 | 82.4% |
| **Diabetes** | | | | | | |
| Yes | 11 | 7.5% | 14 | 9.5% | 25 | 8.5% |
| No | 136 | 92.5% | 134 | 90.5% | 270 | 91.5% |
| **Gender** | | | | | | |
| Male | 76 | 51.7% | 76 | 51.4% | 152 | 51.5% |
| Female | 71 | 48.3% | 72 | 48.6% | 143 | 48.5% |

**Table S3: Baseline characteristics of participants – split by treatment group and overall (ITT population)**

| | Azithromycin (n = 147) | | | Standard Care (n = 148) | | | Total (n = 295) | | |
|---|---|---|---|---|---|---|---|---|---|
| **Continuous** | **Mean (SD)** | **Median (IQR)** | **Range** | **Mean (SD)** | **Median (IQR)** | **Range** | **Mean (SD)** | **Median (IQR)** | **Range** |
| **Age(years)** | 45.5 (14.2) | 46.0 (20.0) | (18, 83) | 46.3 (15.5) | 44.0 (21.5) | (19, 91) | 45.9 (14.9) | 45.0 (22.0) | (18,91) |
| **COS*** | 6.4 (3.6) | 6.0 (4.5) | (0, 17) | 7.0 (3.9) | 6.0 (5.0) | (0, 17) | 6.7 (3.8) | 6.0 (5.0) | (0,17) |
| **COS Plus*** | 7.7 (4.6) | 7.0 (6.0) | (0, 21) | 8.9 (5.2) | 8.0 (7.0) | (0, 22) | 8.3 (5.0) | 7.0 (6.0) | (0,22) |
| **CCI**** | 1.1 (1.5) | 1.0 (2.0) | (0, 10) | 1.2 (1.8) | 0.0 (2.0) | (0, 9) | 1.1 (1.7) | 0.0 (2.0) | (0,10) |
| **Duration of symptoms (days)** | 5.8 (3.5) | 6.0 (5.0) | (0, 14) | 6.3 (3.5) | 6.5 (7.0) | (0, 14) | 6.0 (3.5) | 6.0 (6.0) | (0,14) |
| **Categorical** | **n** | **%** | | **n** | **%** | | **n** | **%** | |
| **Ethnicity** | | | | | | | | | |
| White | 103 | 70.1% | | 98 | 66.2% | | 201 | 68.1% | |
| Mixed | 0 | 0.0% | | 4 | 2.7% | | 4 | 1.4% | |
| Asian/ Asian British | 23 | 15.6% | | 24 | 16.2% | | 47 | 15.9% | |
| Black/ Black British | 6 | 4.1% | | 5 | 3.4% | | 11 | 3.7% | |
| Other Ethnic Group | 15 | 10.2% | | 17 | 11.5% | | 32 | 10.8% | |
| **Smoking** | | | | | | | | | |
| Never smoked | 81 | 57.4% | | 76 | 53.1% | | 157 | 55.3% | |
| Ex-smoker | 25 | 17.7% | | 26 | 18.2% | | 51 | 18.0% | |
| Current smoker | 16 | 11.3% | | 17 | 11.9% | | 33 | 11.6% | |
| Ex-smoker & current vaper | 3 | 2.1% | | 4 | 2.8% | | 7 | 2.5% | |
| Never smoked and current vaper | 0 | 0.0% | | 1 | 0.7% | | 1 | 0.4% | |
| Not recorded | 16 | 11.3% | | 19 | 13.3% | | 35 | 12.3% | |
| **Residence** | | | | | | | | | |
| Non-residential care | 132 | 91.7% | | 137 | 95.1% | | 269 | 93.4% | |
| Residential care | 7 | 4.9% | | 3 | 2.1% | | 10 | 3.5% | |
| No fixed address | 5 | 3.5% | | 4 | 2.8% | | 9 | 3.1% | |
| **Live alone** | | | | | | | | | |
| Yes | 17 | 13.6% | | 13 | 10.6% | | 30 | 12.1% | |
| No | 108 | 86.4% | | 110 | 89.4% | | 218 | 87.9% | |
| **Work Status** | | | | | | | | | |
| Retired | 15 | 10.9% | | 23 | 16.5% | | 38 | 13.7% | |
| Working | 101 | 73.2% | | 95 | 68.3% | | 196 | 70.8% | |
| Houseperson | 22 | 15.9% | | 21 | 15.1% | | 43 | 15.5% | |
| **Occupation** | | | | | | | | | |
| Not Healthcare related | 77 | 78.6% | | 69 | 74.2% | | 146 | 76.4% | |
| Healthcare worker | 20 | 20.4% | | 23 | 24.7% | | 43 | 22.5% | |

| | | | | | | |
|---|---|---|---|---|---|---|
| Laboratory worker | 1 | 1.0% | 1 | 1.1% | 2 | 1.0% |
| **Have asthma** | | | | | | |
| Yes | 26 | 17.7% | 27 | 18.2% | 53 | 18.0% |
| No | 121 | 82.3% | 121 | 81.8% | 242 | 82.0% |
| **History of previous myocardial infarction** | | | | | | |
| Yes | 5 | 3.4% | 7 | 4.7% | 12 | 4.1% |
| No | 142 | 96.6% | 141 | 95.3% | 283 | 95.9% |
| **Currently undergoing any cancer treatment** | | | | | | |
| Yes | 1 | 0.7% | 0 | 0.0% | 1 | 0.3% |
| No | 146 | 99.3% | 148 | 100.0% | 294 | 99.7% |
| **Have chronic pulmonary disease** | | | | | | |
| Yes | 7 | 4.8% | 5 | 3.4% | 12 | 4.1% |
| No | 140 | 95.2% | 143 | 96.6% | 283 | 95.9% |

**Note: COVID-19 COS Score of clinical symptoms is a total score of six common and important clinical symptoms, including fever, cough, fatigue, shortness of breath, diarrhoea, and body pain, each of which can be scored as 0 (no), 1 (mild), 2 (moderate), or 3 (significant). In ATOMIC2, an amended version COVID-19 COS PLUS is also designed with 2 extra clinical symptoms that also considered as having clinical importance: changes to sense of smell and loss of taste. COS scores range from 0 to 18, whereas COS Plus scores range from 0 to 24. Higher scores indicating patient has more significant Covid symptoms.*

***Note: The Charlson Comorbidity Index assigns a numerical value or "weight" from 1,2,3 or 6 to nineteen specific chronic illnesses. The final score (range 0-42) is simply the sum of weighted values with higher scores indicating more comorbidities.*

**Table S4: Clinical assessment outcome measure at baseline – split by treatment group and overall**

| | Azithromycin (n = 147) | | Standard Care (n = 148) | | Total (n = 295) | |
|---|---|---|---|---|---|---|
| | n | % | n | % | n | % |
| **The severity scale score (SSS)*** | | | | | | |
| Ambulatory, no limitation of activities. | 61 | 41.5% | 66 | 44.6% | 127 | 43.1% |
| Limitation of simple activities | 85 | 57.8% | 81 | 54.7% | 166 | 56.3% |
| Hospitalised, mild disease, no oxygen therapy | 1 | 0.7% | 1 | 0.7% | 2 | 0.7% |
| **Pneumonia**** | | | | | | |
| Yes | 28 | 19.0% | 34 | 23.0% | 62 | 21.0% |
| No | 119 | 81.0% | 114 | 77.0% | 233 | 79.0% |
| Not available | 0 | 0.0% | 0 | 0.0% | 0 | 0.0% |
| **Swab results***** | | | | | | |
| Positive | 76 | 51.7% | 76 | 51.4% | 152 | 51.5% |
| Negative | 41 | 27.9% | 38 | 25.7% | 79 | 26.8% |
| Failed Assay | 0 | 0.0% | 3 | 2.0% | 3 | 1.0% |
| Not available | 30 | 20.4% | 31 | 20.9% | 61 | 20.7% |

**Note: The severity scale scores (SSS) range from 0 to 8 with higher scores indicating the most severe status, death.*

***Note: Pneumonia is defined as 'consolidation on a chest X-ray', if a chest X-ray was not taken it is assumed there was no pneumonia.*

****Note: SWAB test results are only available for those who had a Covid-19 swab at randomisation.*

**Table S5: Comorbidities – split by treatment group and overall**

| | Azithromycin (n = 147) | | Standard Care (n = 148) | | Total (n = 295) | |
|---|---|---|---|---|---|---|
| | **n** | **%** | **n** | **%** | **n** | **%** |
| **History of myocardial infarction** | 5 | 3.4% | 7 | 4.7% | 12 | 4.1% |
| **Congestive heart failure** | 1 | 0.7% | 3 | 2.0% | 4 | 1.4% |
| **Peripheral vascular disease** | 2 | 1.4% | 1 | 0.7% | 3 | 1.0% |
| **Cerebrovascular disease** | 1 | 0.7% | 1 | 0.7% | 2 | 0.7% |
| **Dementia** | 0 | 0.0% | 0 | 0.0% | 0 | 0.0% |
| **Chronic pulmonary disease** | 7 | 4.8% | 5 | 3.4% | 12 | 4.1% |
| **Rheumatic disease** | 3 | 2.0% | 6 | 4.1% | 9 | 3.1% |
| **Peptic ulcer disease** | 1 | 0.7% | 1 | 0.7% | 2 | 0.7% |
| **Mild liver disease** | 3 | 2.0% | 1 | 0.7% | 4 | 1.4% |
| **Diabetes without end organ damage** | 11 | 7.5% | 14 | 9.5% | 25 | 8.5% |
| **Hemiplegia** | 0 | 0.0% | 0 | 0.0% | 0 | 0.0% |
| **Moderate/severe renal disease** | 1 | 0.7% | 0 | 0.0% | 1 | 0.3% |
| **Diabetes with end organ damage** | 3 | 2.0% | 2 | 1.4% | 5 | 1.7% |
| **Any malignancy** | 3 | 2.0% | 5 | 3.4% | 8 | 2.7% |
| **Leukaemia** | 0 | 0.0% | 0 | 0.0% | 0 | 0.0% |
| **Lymphoma** | 0 | 0.0% | 0 | 0.0% | 0 | 0.0% |
| **Moderate/severe liver disease** | 0 | 0.0% | 1 | 0.7% | 1 | 0.3% |
| **Metastatic solid tumour** | 1 | 0.7% | 1 | 0.7% | 2 | 0.7% |
| **AIDS** | 0 | 0.0% | 2 | 1.4% | 2 | 0.7% |
| **No comorbidities** | 107 | 76.4% | 107 | 74.3% | 214 | 75.4% |
| **≥1 comorbidity** | 33 | 23.6% | 37 | 25.7% | 70 | 24.6% |

*Note: The comorbidities were collected through The Charlson Comorbidity Index*

**Table S6: Baseline characteristics of participants – split by SWAB results and overall**

| | SWAB +ve (n=152) | | | SWAB -ve & no swab result (n=143) | | | Total (n=295) | | |
|---|---|---|---|---|---|---|---|---|---|
| **Continuous** | **Mean (SD)** | **Median (IQR)** | **Range** | **Mean (SD)** | **Median (IQR)** | **Range** | **Mean (SD)** | **Median (IQR)** | **Range** |
| **Age(years)** | 47.7 (14.7) | 47.0 (20.0) | (19, 91) | 44.0 (14.9) | 43.0 (20.0) | (18, 86) | 45.9 (14.9) | 45.0 (22.0) | (18,91) |
| **COS*** | 7.4 (4.0) | 7.0 (6.0) | (0, 17) | 6.0 (3.4) | 6.0 (4.0) | (0, 17) | 6.7 (3.8) | 6.0 (5.0) | (0,17) |
| **COS Plus*** | 9.3 (5.4) | 8.0 (9.0) | (0, 21) | 7.2 (4.4) | 6.0 (6.0) | (0, 22) | 8.3 (5.0) | 7.0 (6.0) | (0,22) |
| **CCI**** | 1.2 (1.7) | 1.0 (2.0) | (0, 10) | 1.1 (1.6) | 0.0 (2.0) | (0, 9) | 1.1 (1.7) | 0.0 (2.0) | (0,10) |
| **Duration of symptoms (days)** | 6.9 (3.3) | 7.0 (6.0) | (0, 14) | 5.1 (3.5) | 4.0 (5.0) | (0, 14) | 6.0 (3.5) | 6.0 (6.0) | (0,14) |
| **Categorical** | **n** | **%** | | **n** | **%** | | **n** | **%** | |
| **Ethnicity** | | | | | | | | | |
| White | 86 | 56.6% | | 115 | 80.4% | | 201 | 68.1% | |
| Mixed | 4 | 2.6% | | 0 | 0.0% | | 4 | 1.4% | |
| Asian/ Asian British | 34 | 22.4% | | 13 | 9.1% | | 47 | 15.9% | |
| Black/ Black British | 7 | 4.6% | | 4 | 2.8% | | 11 | 3.7% | |
| Other Ethnic Group | 21 | 13.8% | | 11 | 7.7% | | 32 | 10.8% | |
| **Smoking** | | | | | | | | | |
| Never smoked | 99 | 67.8% | | 58 | 42.0% | | 157 | 55.3% | |
| Ex-smoker | 24 | 16.4% | | 27 | 19.6% | | 51 | 18.0% | |
| Current smoker | 8 | 5.5% | | 25 | 18.1% | | 33 | 11.6% | |
| Ex-smoker & current vaper | 3 | 2.1% | | 4 | 2.9% | | 7 | 2.5% | |
| Never smoked and current vaper | 0 | 0.0% | | 1 | 0.7% | | 1 | 0.4% | |
| Not recorded | 12 | 8.2% | | 23 | 16.7% | | 35 | 12.3% | |
| **Residence** | | | | | | | | | |
| Non-residential care | 142 | 95.3% | | 127 | 91.4% | | 269 | 93.4% | |
| Residential care | 6 | 4.0% | | 4 | 2.9% | | 10 | 3.5% | |
| No fixed address | 1 | 0.7% | | 8 | 5.8% | | 9 | 3.1% | |
| **Live alone** | | | | | | | | | |
| Yes | 10 | 7.5% | | 20 | 17.4% | | 30 | 12.1% | |
| No | 123 | 92.5% | | 95 | 82.6% | | 218 | 87.9% | |
| **Work Status** | | | | | | | | | |
| Retired | 22 | 15.1% | | 16 | 12.2% | | 38 | 13.7% | |
| Working | 104 | 71.2% | | 92 | 70.2% | | 196 | 70.8% | |
| Houseperson | 20 | 13.7% | | 23 | 17.6% | | 43 | 15.5% | |
| **Occupation** | | | | | | | | | |

| | | | | | | |
|---|---|---|---|---|---|---|
| Not Healthcare related | 70 | 69.3% | 76 | 84.4% | 146 | 76.4% |
| Healthcare worker | 29 | 28.7% | 14 | 15.6% | 43 | 22.5% |
| Laboratory worker | 2 | 2.0% | 0 | 0.0% | 2 | 1.0% |
| **Have asthma** | | | | | | |
| Yes | 27 | 17.8% | 26 | 18.2% | 53 | 18.0% |
| No | 125 | 82.2% | 117 | 81.8% | 242 | 82.0% |
| **History of previous myocardial infarction** | | | | | | |
| Yes | 8 | 5.3% | 4 | 2.8% | 12 | 4.1% |
| No | 144 | 94.7% | 139 | 97.2% | 283 | 95.9% |
| **Currently undergoing any cancer treatment** | | | | | | |
| Yes | 0 | 0.0% | 1 | 0.7% | 1 | 0.3% |
| No | 152 | 100.0% | 142 | 99.3% | 294 | 99.7% |
| **Have chronic pulmonary disease** | | | | | | |
| Yes | 5 | 3.3% | 7 | 4.9% | 12 | 4.1% |
| No | 147 | 96.7% | 136 | 95.1% | 283 | 95.9% |

**Note: COVID-19 COS Score of clinical symptoms is a total score of six common and important clinical symptoms, including fever, cough, fatigue, shortness of breath, diarrhoea, and body pain, each of which can be scored as 0 (no), 1 (mild), 2 (moderate), or 3 (significant). In ATOMIC2, an amended version COVID-19 COS PLUS is also designed with 2 extra clinical symptoms that also considered as having clinical importance: changes to sense of smell and loss of taste. COS scores range from 0 to 18, whereas COS Plus scores range from 0 to 24. Higher scores indicating patient has more significant Covid symptoms.*

***Note: The Charlson Comorbidity Index assigns a numerical value or "weight" from 1,2,3 or 6 to nineteen specific chronic illnesses. The final score (range 0-42) is simply the sum of weighted values with higher scores indicating more comorbidities.*

**Table S7: Demographic characteristics of participants – total population compared with those hospitalised**

| | Hospitalised (n=32) | | | Total (n=295) | | |
|---|---|---|---|---|---|---|
| **Continuous** | **Mean (SD)** | **Median (IQR)** | **Range** | **Mean (SD)** | **Median (IQR)** | **Range** |
| **Age(years)** | 55.1 (12.2) | 54.0 (14.0) | (27, 82) | 45.9 (14.9) | 45.0 (22.0) | (18, 91) |
| **COS*** | 7.2 (3.5) | 7.0 (4.0) | (1, 15) | 6.7 (3.8) | 6.0 (5.0) | (0, 17) |
| **COS Plus*** | 8.7 (5.1) | 8.0 (6.0) | (1, 21) | 8.3 (5.0) | 7.0 (6.0) | (0, 22) |
| **CCI**** | 1.5 (1.3) | 1.0 (1.0) | (0, 5) | 1.1 (1.7) | 0.0 (2.0) | (0, 10) |
| **Duration of symptoms (days)** | 5.4 (3.2) | 5.0 (4.0) | (0, 13) | 6.0 (3.5) | 6.0 (6.0) | (0, 14) |
| **Categorical** | **n** | **%** | | **n** | **%** | |
| **Ethnicity** | | | | | | |
| White | 20 | 62.5% | | 201 | 68.1% | |
| Mixed | 1 | 3.1% | | 4 | 1.4% | |
| Asian or Asian British | 6 | 18.8% | | 47 | 15.9% | |
| Black or Black British | 1 | 3.1% | | 11 | 3.7% | |
| Other Ethnic Group | 4 | 12.5% | | 32 | 10.8% | |
| **Smoking** | | | | | | |
| Never smoked | 16 | 51.6% | | 157 | 55.3% | |
| Ex-smoker | 11 | 35.5% | | 51 | 18.0% | |
| Current smoker | 0 | 0.0% | | 33 | 11.6% | |
| Ex-smoker & current vaper | 0 | 0.0% | | 7 | 2.5% | |
| Never smoked and current vaper | 0 | 0.0% | | 1 | 0.4% | |
| Not recorded | 4 | 12.9% | | 35 | 12.3% | |
| **Residence** | | | | | | |
| Non-residential care | 29 | 90.6% | | 269 | 93.4% | |
| Residential care | 1 | 3.1% | | 10 | 3.5% | |
| No fixed address | 2 | 6.3% | | 9 | 3.1% | |
| **Live alone** | | | | | | |
| Yes | 2 | 7.1% | | 30 | 12.1% | |
| No | 26 | 92.9% | | 218 | 87.9% | |
| **Work Status** | | | | | | |
| Retired | 9 | 28.1% | | 38 | 13.7% | |
| Working | 17 | 53.1% | | 196 | 70.8% | |
| Houseperson | 6 | 18.8% | | 43 | 15.5% | |
| **Occupation** | | | | | | |
| Not Healthcare related | 13 | 76.5% | | 146 | 76.4% | |
| Healthcare worker | 4 | 23.5% | | 43 | 22.5% | |
| Laboratory worker | 0 | 0.0% | | 2 | 1.0% | |

| | | | | |
|---|---|---|---|---|
| **Have asthma** | | | | |
| Yes | 8 | 25.0% | 53 | 18.0% |
| No | 24 | 75.0% | 242 | 82.0% |
| **History of previous myocardial infarction** | | | | |
| Yes | 1 | 3.1% | 12 | 4.1% |
| No | 31 | 96.9% | 283 | 95.9% |
| **Currently undergoing any cancer treatment** | | | | |
| Yes | 0 | 0.0% | 1 | 0.3% |
| No | 32 | 100.0% | 294 | 99.7% |
| **Have chronic pulmonary disease** | | | | |
| Yes | 3 | 9.4% | 12 | 4.1% |
| No | 29 | 90.6% | 283 | 95.9% |

**Note: COVID-19 COS Score of clinical symptoms is a total score of six common and important clinical symptoms, including fever, cough, fatigue, shortness of breath, diarrhoea, and body pain, each of which can be scored as 0 (no), 1 (mild), 2 (moderate), or 3 (significant). In ATOMIC2, an amended version COVID-19 COS PLUS is also designed with 2 extra clinical symptoms that also considered as having clinical importance: changes to sense of smell and loss of taste. COS scores range from 0 to 18, whereas COS Plus scores range from 0 to 24. Higher scores indicating patient has more significant Covid symptoms.*

***Note: The Charlson Comorbidity Index assigns a numerical value or "weight" from 1,2,3 or 6 to nineteen specific chronic illnesses. The final score (range 0-42) is simply the sum of weighted values with higher scores indicating more comorbidities.*

#### 3 Compliance

##### 3.1 Description of Compliance with Intervention

Subjects were randomised to receive Azithromycin 500 mg daily orally for 14 days in addition to standard care or standard care alone.

For patients allocated to AZM, the first dose was to be taken within 4 hours of randomisation. Participants were asked to take AZM at the approximately the same time every day for 14 days. The drug should be taken ideally 1 hour before a meal or 2 hours afterwards. Participants were asked to take two 250mg capsules each day to ensure a dose of 500mg is taken.

Compliance was assessed by telephone discussion with patient on day 14 of treatment with specific questioning as to the number of pills remaining. Adequate compliance is defined as the first dose being administered within 4 hours of randomisation and at least 80% of doses i.e., a maximum of 4/28 tablets remaining at the end of day 14. Table S8 summarises the number of participants allocated to azithromycin who started treatment and who complied with treatment (≤4 tablets left). The average number of tablets taken for participants who did and did not comply is also summarised.

**Table S8: Compliance with allocated treatment (AZM arm only)**

| | **Intervention (AZM)** |
|---|---|
| **Started treatment (n, %)** | 143 (97.3%) |
| **Compliance (n, %)*** | |
| ***Complier*** | 76 (51.7%) |
| ***Non-complier*** | 51 (34.7%) |
| ***Unknown*** | 20 (13.6%) |
| **Number of tablets taken (median, IQR, N)*** | |
| Compliers | 28 (28, 28), 73 |
| Non-compliers | 6 (2, 17), 44 |

**Note: at most sites the number of tablets dispensed was 30 and the number of tablets taken is calculated accordingly. In some instances the compliance status of a participant may be known despite the exact number of tablets taken being unknown.*

##### 3.2 Withdrawals

Three participants withdrew from this trial and requested that all data collected up to the point withdrawal be removed. In addition, a further three participants withdrew from follow-up and did not provide primary outcome data. Details of these participants, including allocated treatment, time of withdrawal, reasons for withdrawal and last questionnaire completed are summarised in Table S9.

**Table S9: Details of withdrawals from follow-up**

| Treatment | Time to withdrawal (days) | Requested by | Reason | Last questionnaire |
|---|---|---|---|---|
| Azithromycin | 14 | Participant | Other reason | Baseline |
| Azithromycin | 0 | Clinician/researcher | Other reason | None |
| Standard care | 14 | Clinician/researcher | Other reason | Baseline |

#### 4 Results

##### 4.1 Primary outcomes

Full details on the number and proportion of participants who had been hospitalised or had died by day 28 (Table S10) and the details of participants that had been hospitalised including time to hospitalisation (Table S11) are summarised by treatment arm. Histograms of time to hospitalisation by treatment group are presented in Figure S1. The difference in proportions between the treatment arms was assessed using a chi-squared test and a 5% (2-sided) significance level. Relative and absolute differences in proportions together with the 95% confidence intervals are reported. Adjusted analyses were undertaken using logistic regression with progression as the binary outcome, adjusting for stratification factors (centre, hypertension, diabetes, and sex). The results of these analyses are reported in Table S12, no significant differences were identified. An analysis adjusting other important prognostic variables (age, chronic pulmonary disease, and presence of cancer which was determined by the presence of any of the following four items: malignancy, leukaemia, lymphoma, and metastatic solid tumour in the Charlson comorbidity index) in addition to the stratification factors was undertaken as a supporting analysis.

Supporting analyses using time-to-event methodology were undertaken log rank tests and Cox proportional hazards regression methods to explore whether the intervention delays progression to hospitalisation or death. Unadjusted, adjusted and fully adjusted results are provided in Table 12 with effect estimates summarised using hazard ratios and associated 95% CIs.

The primary outcome was analysed on the Intention to treat (ITT) population and repeated on the ITT+ve population if there is information available on the results of baseline swab tests. The same analysis methods as described above were undertaken on both populations.

**Table S10: Summary of primary outcome population by intervention groups**

| | **Azithromycin** | | **Standard Care** | | **Total** | |
|---|---|---|---|---|---|---|
| | **n/N** | **%** | **n/N** | **%** | **n/N** | **%** |
| **Total hospitalised** | 15/145 | 10.3% | 17/147 | 11.6% | 32/292 | 11.0% |
| Received level 2 ventilation* | 1/145 | 0.7% | 1/147 | 0.7% | 2/292 | 0.7% |
| Received level 3 ventilation* | 0/145 | 0.0% | 0/147 | 0.0% | 0/292 | 0.0% |
| Did not receive level 2 or level 3 ventilation | 14/145 | 9.7% | 16/147 | 10.9% | 30/292 | 10.3% |
| **Total died**** | 1/145 | 0.7% | 1/147 | 0.7% | 2/292 | 0.7% |

** Note: Participants could have received both level 2 and level 3 ventilations during hospitalisation.*

*** Note: Both deaths occurred following hospitalisation.*

**Table S11: Details of participants that been hospitalised – split by treatment group**

| | Azithromycin | | | Standard Care | | | Total | | |
|---|---|---|---|---|---|---|---|---|---|
| | **Mean (SD)** | **Median (IQR)** | **Range** | **Mean (SD)** | **Median (IQR)** | **Range** | **Mean (SD)** | **Median (IQR)** | **Range** |
| **Age (years)** | 53.27 (11.00) | 51.00 (9.00) | (38, 79) | 56.76 (13.36) | 60.00 (14.00) | (27, 82) | 55.13 (12.24) | 54.00 (14.00) | (27,82) |
| **Time to first hospitalisation (days)** | 2.33 (1.50) | 2.00 (3.00) | (0, 5) | 6.65 (7.36) | 4.00 (5.00) | (0, 27) | 4.63 (5.81) | 3.50 (4.00) | (0,27) |
| **COS at first admission** | 8.27 (3.20) | 7.00 (4.00) | (4, 15) | 6.82 (3.50) | 7.00 (5.00) | (1, 15) | 7.39 (3.40) | 7.00 (4.50) | (1,15) |
| **COS at last discharge** | 3.25 (3.28) | 3.00 (5.50) | (0, 9) | 2.53 (1.96) | 2.00 (3.00) | (0, 6) | 2.78 (2.45) | 3.00 (5.00) | (0,9) |
| **COS Plus at first admission** | 9.00 (4.57) | 7.50 (4.00) | (4, 18) | 7.59 (4.21) | 7.00 (7.00) | (1, 15) | 8.11 (4.32) | 7.00 (7.00) | (1,18) |
| **COS Plus at last discharge** | 5.00 (5.35) | 4.50 (8.00) | (0, 15) | 3.00 (2.45) | 3.00 (3.00) | (0, 9) | 3.70 (3.72) | 3.00 (6.00) | (0,15) |
| **SSS at first admission** | 2.50 (0.94) | 2.00 (1.00) | (1, 5) | 2.47 (1.01) | 3.00 (1.00) | (1, 5) | 2.48 (0.96) | 2.00 (1.00) | (1,5) |
| **SSS at last discharge** | 2.21 (1.81) | 2.00 (0.00) | (0, 8) | 2.00 (1.73) | 2.00 (1.00) | (0, 8) | 2.10 (1.74) | 2.00 (1.00) | (0,8) |
| **Total inpatient days** | 4.38 (2.84) | 4.00 (4.00) | (1, 11) | 4.06 (3.15) | 3.50 (4.00) | (1, 12) | 4.21 (2.97) | 4.00 (4.00) | (1,12) |

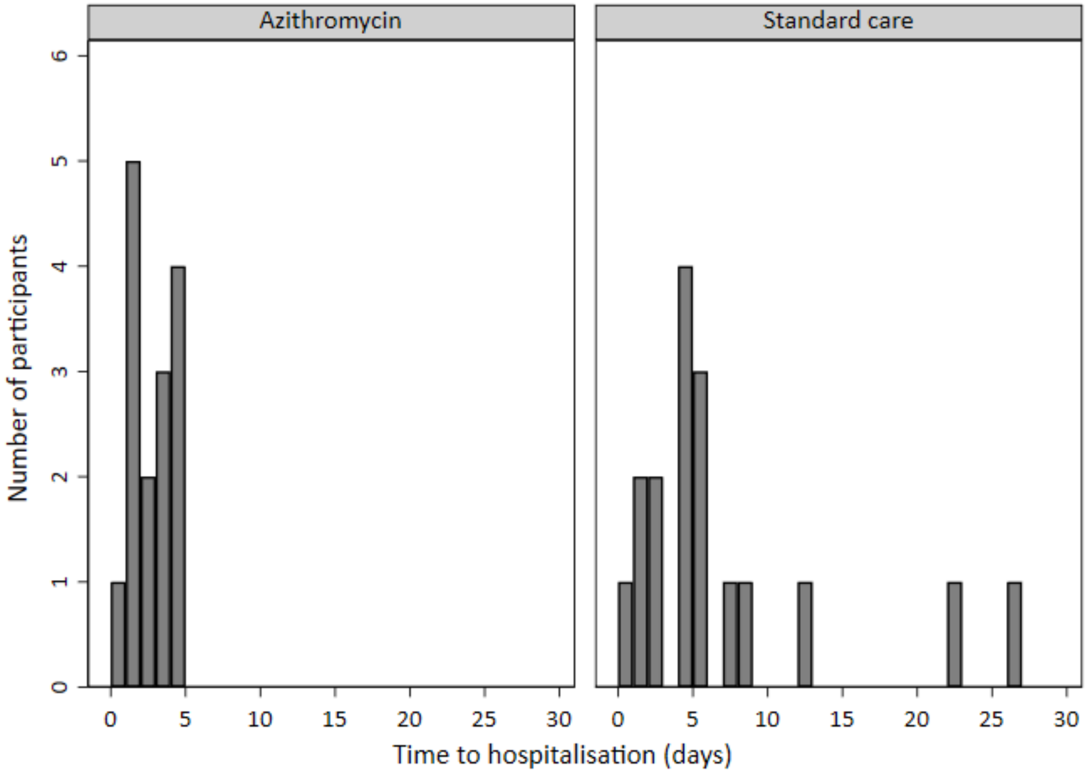

**Figure S1: Histograms of time to hospitalisation (amongst hospitalised patients) by treatment group**

**Table S12: Primary outcome by intervention groups**

| | Azithromycin | | Standard Care | | | Comparison of proportions | | Time-to-event |
|---|---|---|---|---|---|---|---|---|
| | n/N | % | n/N | % | | OR (95%CI), p-val | RD (95%CI) | |
| **ITT** | 15/145 | 10.3% | 17/147 | 11.6% | **Unadjusted** | 0.88 (0.42, 1.84), 0.74 | -1.2% (-8.4%, 5.9%) | 0.79 |
| | | | | | **Adjusted*** | 0.91 (0.43, 1.92), 0.80 | -1.0% (-8.0%, 6.1%) | 0.95 (0.46, 1.96), 0.89 |
| | | | | | **Fully Adjusted**** | 0.91 (0.42, 1.97), 0.82 | -1.2% (-8.2%, 5.7%) | 0.99 (0.49, 2.00), 0.99 |
| **ITT +ve** | 11/75 | 14.7% | 11/75 | 14.7% | **Unadjusted** | 1.00 (0.40, 2.47), 1.00 | 0.0% (-11.3%, 11.3%) | 0.78 |
| | | | | | **Adjusted*** | 1.02 (0.40, 2.57), 0.97 | 0.3% (-10.8%, 11.4%) | 1.17 (0.49, 2.77), 0.72 |
| | | | | | **Fully Adjusted**** | 1.11 (0.43, 2.90), 0.83 | 0.8% (-10.1%, 11.7%) | 1.30 (0.52, 3.21), 0.57 |

** Note: Adjust for stratification factors: centre, hypertension, diabetes, and sex.*

*** Note: Adjust for stratification factors and other important prognostic variables: centre, hypertension, diabetes, sex, and age ≥65 years, presence of chronic lung disease, and treatment for cancer.*

##### *4.2 Secondary outcomes*

Sensitivity analyses were conducted to compare the proportion of level 2/3 ventilation or death, all-cause mortality, and progression to pneumonia in the ITT +ve population (Table S13). The level of oxygen support received at each inpatient day is presented for each participant (Figure S2).

**Table S13: Secondary outcomes by intervention groups for the ITT +ve population**

| | **Azithromycin** | | **Standard Care** | | **Fisher's exact** |
|---|---|---|---|---|---|
| | **n** | **%** | **n** | **%** | **p-val** |
| ***Level 2/3 ventilation or death*** | 2/75 | 2.7% | 2/75 | 2.7% | 1.00 |
| ***All-cause mortality*** | 1/75 | 1.3% | 1/75 | 1.3% | 1.00 |
| ***Progression to pneumonia*** | 0/58 | 0.0% | 2/52 | 3.8% | 0.22 |

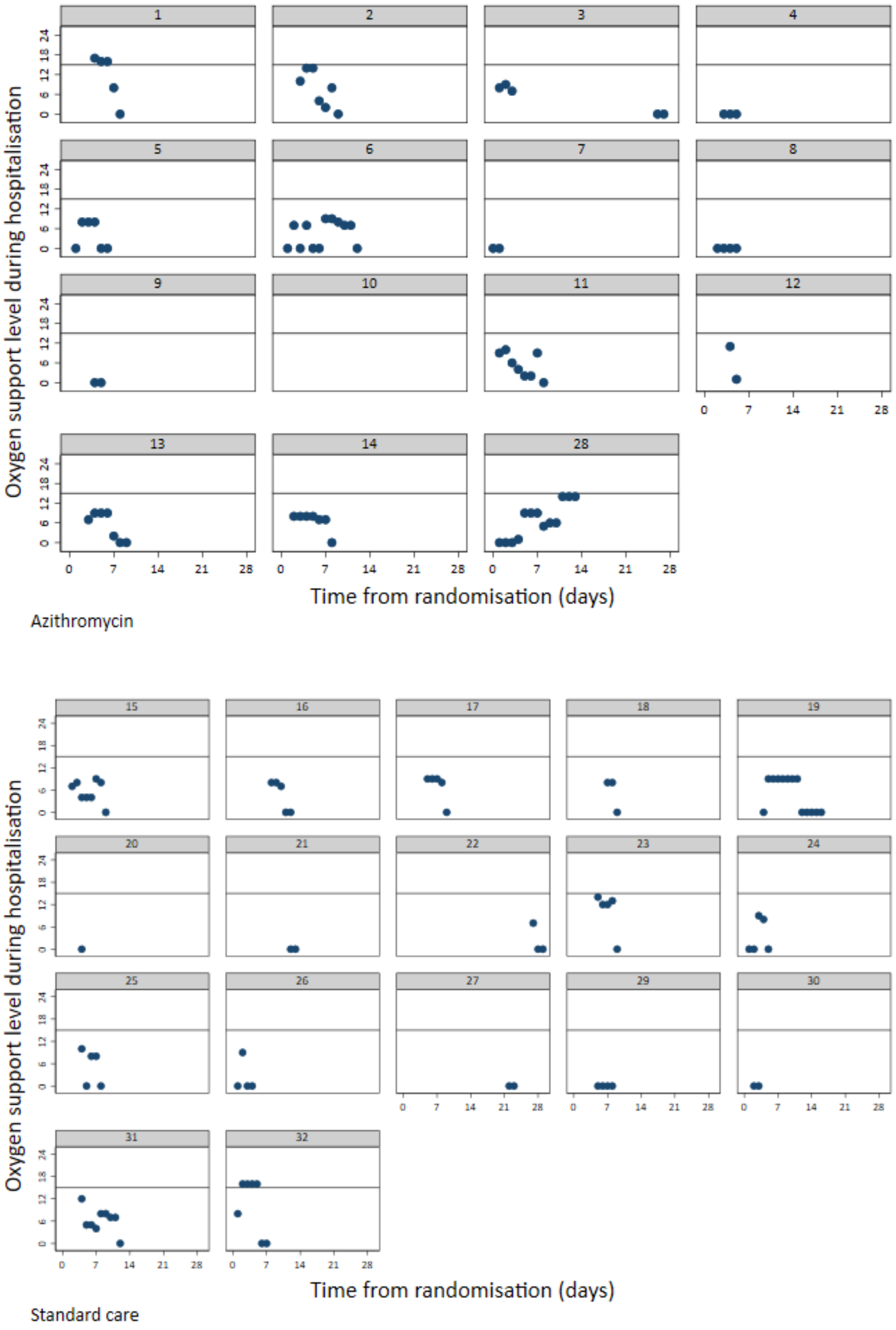

Figure S2: Scatter plot of level of oxygen support received by inpatient day for each participant – by intervention groups (ITT population)

*Note 1: Patient received no oxygen therapy during inpatient days were given score 0.*

*Note 2: The reference line indicates that patient is on level 2 ventilation. For any oxygen received but not yet reached level 2 ventilation is level 1.*

*Any patient had a level 2 or level 3 ventilation is counted as an event.*

The proportion of participants progressing to severe pneumonia by day 28 post-randomisation was analysed within the subset of participants for whom pneumonia was present at time of enrolment. There were no instances of participants progressing to severe pneumonia. Details of participant pneumonia status at baseline and follow-up are summarised in Table S14.

**Table S14: Summary of diagnosis with pneumonia from baseline until end of follow-up by intervention groups**

| | Azithromycin | | Standard Care | | Total | |
|---|---|---|---|---|---|---|
| | n/N | % | n/N | % | n/N | % |
| **ITT** | | | | | | |
| **No pneumonia at baseline** | 119/147 | 81.0% | 114/148 | 77.0% | 233/295 | 79.0% |
| ***Hospitalised - pneumonia*** | 0/119 | 0.0% | 2/114 | 1.8% | 2/233 | 0.9% |
| ***Hospitalised - no pneumonia*** | 5/119 | 4.2% | 5/114 | 4.4% | 10/233 | 4.3% |
| ***Hospitalised - pneumonia status missing*** | 4/119 | 3.4% | 3/114 | 2.6% | 7/233 | 3.0% |
| ***Not hospitalised*** | 108/119 | 90.8% | 104/114 | 91.2% | 212/233 | 91.0% |
| ***Hospitalisation status missing*** | 2/119 | 1.7% | 0/114 | 0.0% | 2/233 | 0.9% |
| **Pneumonia at baseline** | 28/147 | 19.0% | 34/148 | 23.0% | 62/295 | 21.0% |
| ***Hospitalised - pneumonia*** | 5/28 | 17.9% | 2/34 | 5.9% | 7/62 | 11.3% |
| ***Hospitalised - no pneumonia*** | 1/28 | 3.6% | 2/34 | 5.9% | 3/62 | 4.8% |
| ***Hospitalised - pneumonia status missing*** | 0/28 | 0.0% | 3/34 | 8.8% | 3/62 | 4.8% |
| ***Not hospitalised*** | 22/28 | 78.6% | 26/34 | 76.5% | 48/62 | 77.4% |
| ***Hospitalisation status missing*** | 0/28 | 0.0% | 1/34 | 2.9% | 1/62 | 1.6% |
| **ITT +ve** | | | | | | |
| **No pneumonia at baseline** | 58/76 | 76.3% | 52/76 | 68.4% | 110/152 | 72.4% |
| ***Hospitalised - pneumonia*** | 0/58 | 0.0% | 2/52 | 3.8% | 2/110 | 1.8% |
| ***Hospitalised - no pneumonia*** | 4/58 | 6.9% | 2/52 | 3.8% | 6/110 | 5.5% |
| ***Hospitalised - pneumonia status missing*** | 3/58 | 5.2% | 2/52 | 3.8% | 5/110 | 4.5% |
| ***Not hospitalised*** | 50/58 | 86.2% | 46/52 | 88.5% | 96/110 | 87.3% |
| ***Hospitalisation status missing*** | 1/58 | 1.7% | 0/52 | 0.0% | 1/110 | 0.9% |
| **Pneumonia at baseline** | 18/76 | 23.7% | 24/76 | 31.6% | 42/152 | 27.6% |
| ***Hospitalised - pneumonia*** | 3/18 | 16.7% | 2/24 | 8.3% | 5/42 | 11.9% |
| ***Hospitalised - no pneumonia*** | 1/18 | 5.6% | 1/24 | 4.2% | 2/42 | 4.8% |
| ***Hospitalised - pneumonia status missing*** | 0/18 | 0.0% | 2/24 | 8.3% | 2/42 | 4.8% |
| ***Not hospitalised*** | 14/18 | 77.8% | 18/24 | 75.0% | 32/42 | 76.2% |
| ***Hospitalisation status missing*** | 0/18 | 0.0% | 1/24 | 4.2% | 1/42 | 2.4% |

Differences in peak severity of illness

Severity scale scores from baseline to day 28 are summarised by treatment group and overall for the ITT population (Table S15), for the ITT +ve population (Table S15) and for only the hospitalised patients (Table S15, Figure S3). Peak severity score was defined as the highest severity score observed during the follow-up period. This is summarised in Table S16 and Figure S4. The difference between the treatment arms in terms of peak severity score was assessed using ordinal logistic regression. Unadjusted and adjusted analyses were undertaken with stratification factors (centre, hypertension, diabetes, and sex) and odds ratios, 95% CIs and associated p-values are presented (Table S17). The time to the peak severity is also summarised (Table S18).

**Table S15: Description of the severity scale score during the entire study – ITT, ITT +ve and all hospitalised patients**

| | Azithromycin | | | Standard Care | | | Total | | |
|---|---|---|---|---|---|---|---|---|---|
| | **Mean (SD)** | **Median (IQR)** | **Range** | **Mean (SD)** | **Median (IQR)** | **Range** | **Mean (SD)** | **Median (IQR)** | **Range** |
| **ITT** | | | | | | | | | |
| ***Baseline*** | 0.59 (0.51) | 1.00 (1.00) | (0, 2) | 0.56 (0.51) | 1.00 (1.00) | (0, 2) | 0.58 (0.51) | 1.00 (1.00) | (0,2) |
| ***Day 14*** | 0.54 (0.92) | 0.00 (1.00) | (0, 8) | 0.52 (0.87) | 0.00 (1.00) | (0, 8) | 0.53 (0.89) | 0.00 (1.00) | (0,8) |
| ***Day 28*** | 0.35 (0.85) | 0.00 (1.00) | (0, 8) | 0.37 (0.86) | 0.00 (1.00) | (0, 8) | 0.36 (0.85) | 0.00 (1.00) | (0,8) |
| **ITT +ve** | | | | | | | | | |
| ***Baseline*** | 0.61 (0.52) | 1.00 (1.00) | (0, 2) | 0.59 (0.49) | 1.00 (1.00) | (0, 1) | 0.60 (0.51) | 1.00 (1.00) | (0,2) |
| ***Day 14*** | 0.73 (1.17) | 1.00 (1.00) | (0, 8) | 0.59 (1.10) | 0.00 (1.00) | (0, 8) | 0.66 (1.13) | 1.00 (1.00) | (0,8) |
| ***Day 28*** | 0.49 (1.10) | 0.00 (1.00) | (0, 8) | 0.48 (1.09) | 0.00 (1.00) | (0, 8) | 0.48 (1.09) | 0.00 (1.00) | (0,8) |
| **All hospitalised patients** | | | | | | | | | |
| ***Baseline*** | 0.87 (0.52) | 1.00 (0.00) | (0, 2) | 0.47 (0.51) | 0.00 (1.00) | (0, 1) | 0.66 (0.55) | 1.00 (1.00) | (0,2) |
| ***Day 14*** | 1.64 (2.02) | 1.00 (0.00) | (0, 8) | 1.00 (1.97) | 0.50 (1.00) | (0, 8) | 1.30 (1.99) | 1.00 (1.00) | (0,8) |
| ***Day 28*** | 1.00 (2.16) | 0.00 (1.00) | (0, 8) | 0.81 (1.97) | 0.00 (1.00) | (0, 8) | 0.90 (2.02) | 0.00 (1.00) | (0,8) |

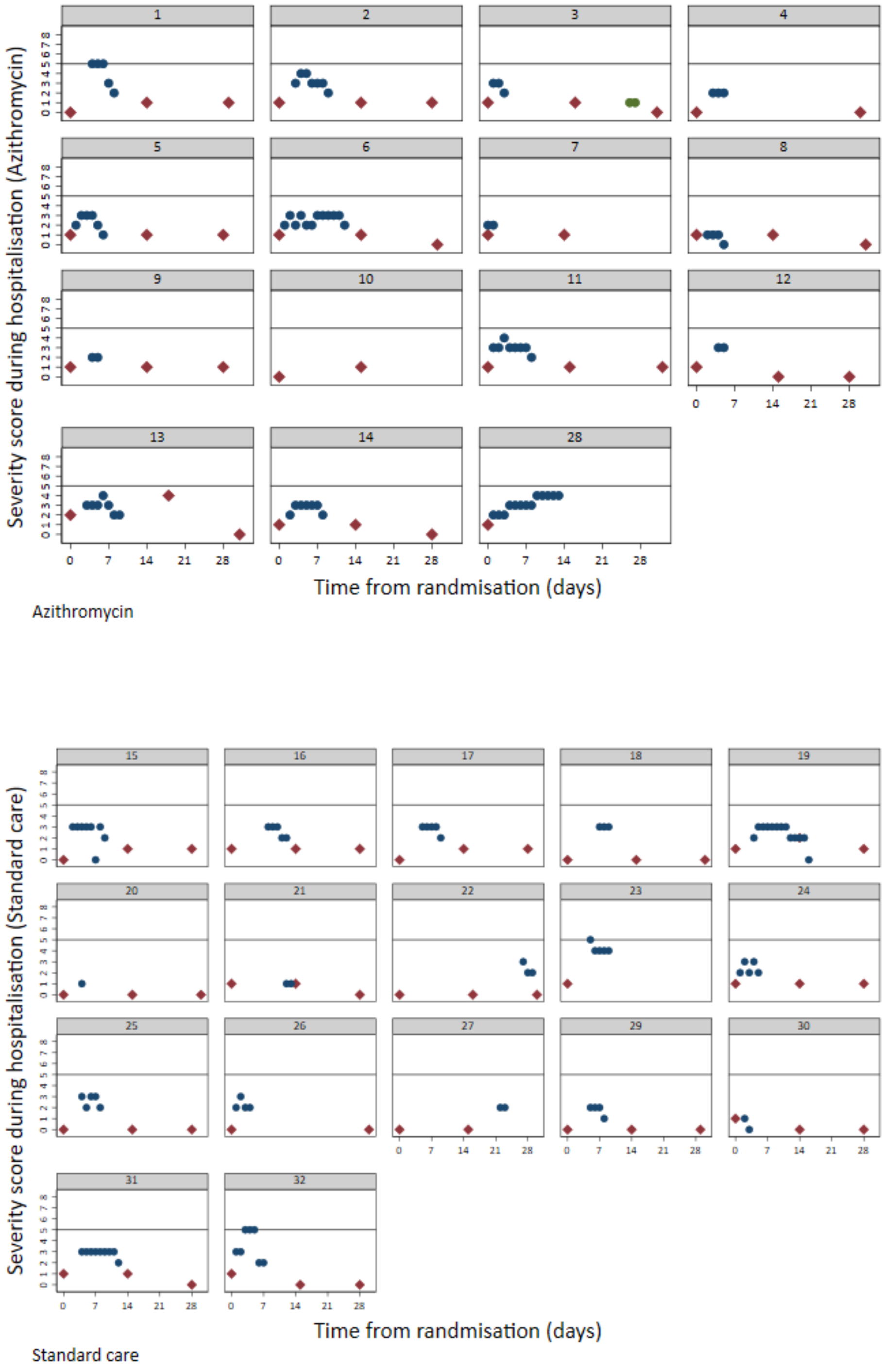

**Figure S3: Scatter plot of the severity scale score at each hospital inpatient days for each patient – by intervention groups (ITT population)**

*Note: the reference line is relevant to participants' clinical status of hospitalised with level 2 or level 3 ventilation.*

**Table S16: Description of peak severity scale score during follow-up – ITT, ITT +ve & all hospitalised patients**

| | Azithromycin | | | Standard Care | | | Total | | |
|---|---|---|---|---|---|---|---|---|---|
| | Mean (SD) | Median (IQR) | Range | Mean (SD) | Median (IQR) | Range | Mean (SD) | Median (IQR) | Range |
| **ITT** | 0.77 (1.18) | 0.50 (1.00) | (0, 8) | 0.79 (1.11) | 1.00 (1.00) | (0, 8) | 0.78 (1.14) | 1.00 (1.00) | (0,8) |
| **ITT +ve** | 1.03 (1.44) | 1.00 (1.00) | (0, 8) | 0.91 (1.35) | 1.00 (1.00) | (0, 8) | 0.97 (1.39) | 1.00 (1.00) | (0,8) |
| **All hospitalised patients** | 3.20 (1.74) | 3.00 (2.00) | (1, 8) | 2.94 (1.64) | 3.00 (1.00) | (1, 8) | 3.06 (1.66) | 3.00 (1.00) | (1,8) |

*Note: Peak severity scale score was assessed post-baseline.*

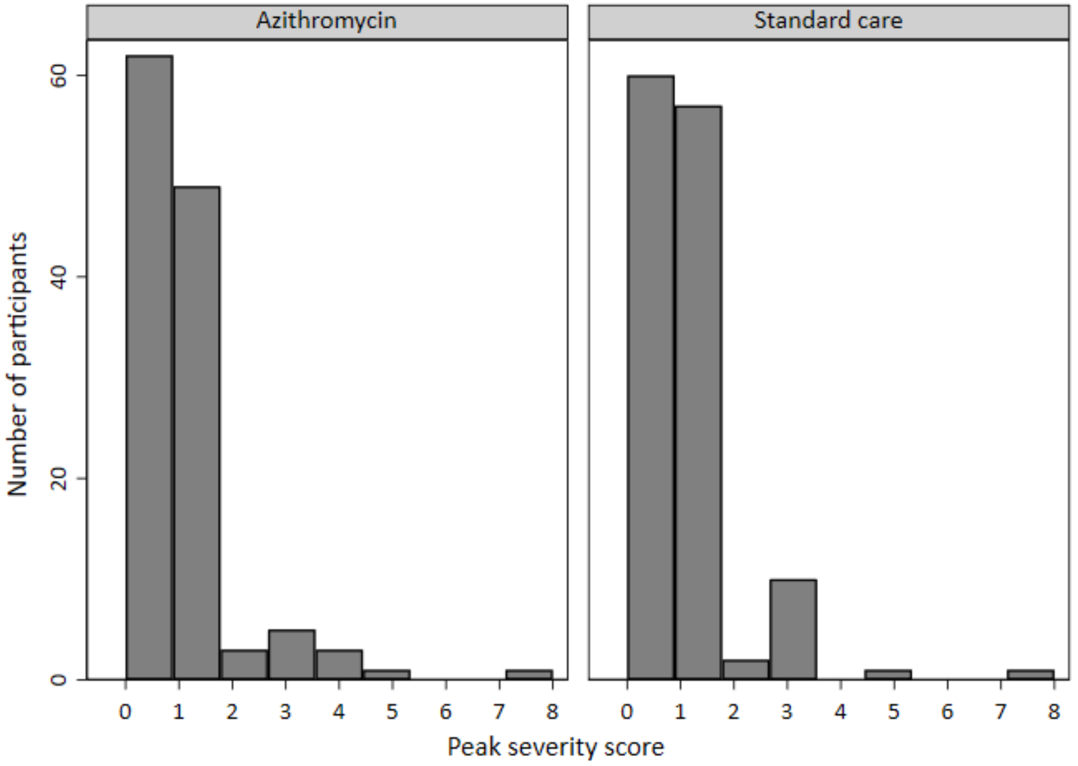

**Figure S4: Histogram of the peak severity scale score during follow-up by intervention groups (ITT)**

*Note: Peak severity is defined as the highest severity score during the follow-up.*

**Table S17: Secondary outcome (differences in peak severity of illness) by intervention groups**

| | Azithromycin | | Standard Care | | Unadjusted | Adjusted* |
|---|---|---|---|---|---|---|
| | n | % | n | % | OR (95%CI), p-val | OR (95%CI), p-val |
| **ITT** | | | | | 0.87 (0.54, 1.40), 0.57 | 0.91 (0.57, 1.46), 0.69 |
| ***Ambulatory, no limitation of activities.*** | 62 | 50.0% | 60 | 45.8% | | |
| ***Limitation of simple activities*** | 49 | 39.5% | 57 | 43.5% | | |
| ***Hospitalised, mild disease, no oxygen therapy*** | 3 | 2.4% | 2 | 1.5% | | |
| ***Hospitalised, oxygen by conventional delivery system ≤40% mask or nasal prongs*** | 5 | 4.0% | 10 | 7.6% | | |
| ***Hospitalised, oxygen by conventional delivery system >40% mask*** | 3 | 2.4% | 0 | 0.0% | | |
| ***Hospitalised receiving non-invasive ventilation or receiving high-flow oxygen therapy (HFOT, >15L/min), or continuous positive airway pressure (CPAP)*** | 1 | 0.8% | 1 | 0.8% | | |
| ***Death*** | 1 | 0.8% | 1 | 0.8% | | |
| **ITT +ve** | | | | | 1.23 (0.65, 2.32), 0.53 | 1.43 (0.73, 2.78), 0.29 |
| ***Ambulatory, no limitation of activities.*** | 26 | 40.0% | 32 | 45.7% | | |
| ***Limitation of simple activities*** | 29 | 44.6% | 28 | 40.0% | | |
| ***Hospitalised, mild disease, no oxygen therapy*** | 2 | 3.1% | 1 | 1.4% | | |
| ***Hospitalised, oxygen by conventional delivery system ≤40% mask or nasal prongs*** | 3 | 4.6% | 7 | 10.0% | | |
| ***Hospitalised, oxygen by conventional delivery system >40% mask*** | 3 | 4.6% | 0 | 0.0% | | |
| ***Hospitalised receiving non-invasive ventilation or receiving high-flow oxygen therapy (HFOT, >15L/min), or continuous positive airway pressure (CPAP)*** | 1 | 1.5% | 1 | 1.4% | | |
| ***Death*** | 1 | 1.5% | 1 | 1.4% | | |

** Note: Adjust for stratification factors: centre, hypertension, diabetes, and sex.*

**Table S18: Time to peak and the time points of peak severity scale score during follow-up**

| | Azithromycin | | Standard Care | | Total | |
|---|---|---|---|---|---|---|
| **Time to peak severity (Days)** | **Median (IQR)** | **Range** | **Median (IQR** | **Range** | **Median (IQR** | **Range** |
| **ITT** | 14.0 (0.0) | (0,28) | 14.0 (0.0) | (2,28) | 14.0 (0.0) | (0,28) |
| **ITT +ve** | 14.0 (0.0) | (2,28) | 14.0 (0.0) | (2,28) | 14.0 (0.0) | (2,28) |
| **Hospitalised** | 3.0 (2.0) | (0,14) | 5.0 (5.0) | (2,27) | 4.0 (4.5) | (0,27) |
| **Time points of peak severity (ITT)** | **n** | **%** | **n** | **%** | **n** | **%** |
| **14 days** | 95 | 76.6% | 97 | 74.0% | 192 | 75.3% |
| **28 days** | 15 | 12.1% | 17 | 13.0% | 32 | 12.5% |
| **During hospitalisation** | 14 | 11.3% | 17 | 13.0% | 31 | 12.2% |

*Note: Peak severity scale score was assessed post-baseline.*

The COVID COS score and COVID COS PLUS

COS and COS plus scores at Baseline, Day 14 and Day 28 were summarised by treatment groups (Table S19). Histograms were used to compare the Baseline, Day 14 and Day 28 COS and COS PLUS between treatment groups (Figure S5). The COVID COS PLUS score at each inpatient day is presented by patients (Figure S6).

**Table S19: Description of the COVID COS score and COVID COS PLUS score during the entire study – ITT & all hospitalised patients**

| | Azithromycin | | | Standard Care | | | Total | | |
|---|---|---|---|---|---|---|---|---|---|
| | **Mean (SD)** | **Median (IQR)** | **Range** | **Mean (SD)** | **Median (IQR)** | **Range** | **Mean (SD)** | **Median (IQR)** | **Range** |
| **COVID COS score** | | | | | | | | | |
| ***ITT*** | | | | | | | | | |
| *Baseline* | 6.36 (3.64) | 6.00 (4.50) | (0, 17) | 7.00 (3.87) | 6.00 (5.00) | (0, 17) | 6.68 (3.77) | 6.00 (5.00) | (0,17) |
| *Day 14* | 6.25 (3.93) | 5.00 (6.00) | (0, 18) | 5.92 (4.03) | 6.00 (6.00) | (0, 16) | 6.09 (3.97) | 6.00 (6.00) | (0,18) |
| *Day 28* | 2.67 (2.92) | 2.00 (4.00) | (0, 18) | 2.92 (2.87) | 2.00 (4.00) | (0, 14) | 2.80 (2.89) | 2.00 (4.00) | (0,18) |
| ***All hospitalised patients*** | | | | | | | | | |
| *Baseline* | 7.64 (3.71) | 7.50 (3.00) | (1, 15) | 6.88 (3.48) | 6.00 (4.00) | (1, 15) | 7.23 (3.55) | 7.00 (4.00) | (1,15) |
| *Day 14* | 7.92 (4.31) | 8.00 (5.00) | (2, 17) | 5.57 (4.42) | 5.00 (5.00) | (0, 15) | 6.70 (4.44) | 6.00 (7.00) | (0,17) |
| *Day 28* | 3.08 (2.43) | 3.50 (3.50) | (0, 8) | 3.13 (2.67) | 2.00 (5.00) | (0, 8) | 3.11 (2.52) | 3.00 (4.00) | (0,8) |
| ***COVID COS PLUS score*** | | | | | | | | | |
| ***ITT*** | | | | | | | | | |
| *Baseline* | 7.66 (4.65) | 7.00 (6.00) | (0, 21) | 8.86 (5.25) | 8.00 (7.00) | (0, 22) | 8.26 (4.98) | 7.00 (6.00) | (0,22) |
| *Day 14* | 7.62 (5.11) | 7.00 (6.00) | (0, 24) | 7.47 (5.19) | 6.50 (8.00) | (0, 21) | 7.55 (5.14) | 7.00 (7.00) | (0,24) |
| *Day 28* | 3.06 (3.61) | 2.00 (4.00) | (0, 24) | 3.38 (3.52) | 2.00 (4.00) | (0, 14) | 3.22 (3.56) | 2.00 (4.00) | (0,24) |
| ***All hospitalised patients*** | | | | | | | | | |
| *Baseline* | 9.29 (5.17) | 8.00 (7.00) | (1, 20) | 8.24 (5.21) | 7.00 (5.00) | (1, 21) | 8.71 (5.13) | 8.00 (6.00) | (1,21) |
| *Day 14* | 9.69 (5.62) | 9.00 (9.00) | (2, 19) | 6.00 (4.61) | 6.00 (7.00) | (0, 15) | 7.78 (5.36) | 7.00 (7.00) | (0,19) |
| *Day 28* | 3.25 (2.63) | 3.50 (3.50) | (0, 8) | 3.53 (3.40) | 2.00 (5.00) | (0, 11) | 3.41 (3.03) | 3.00 (4.00) | (0,11) |

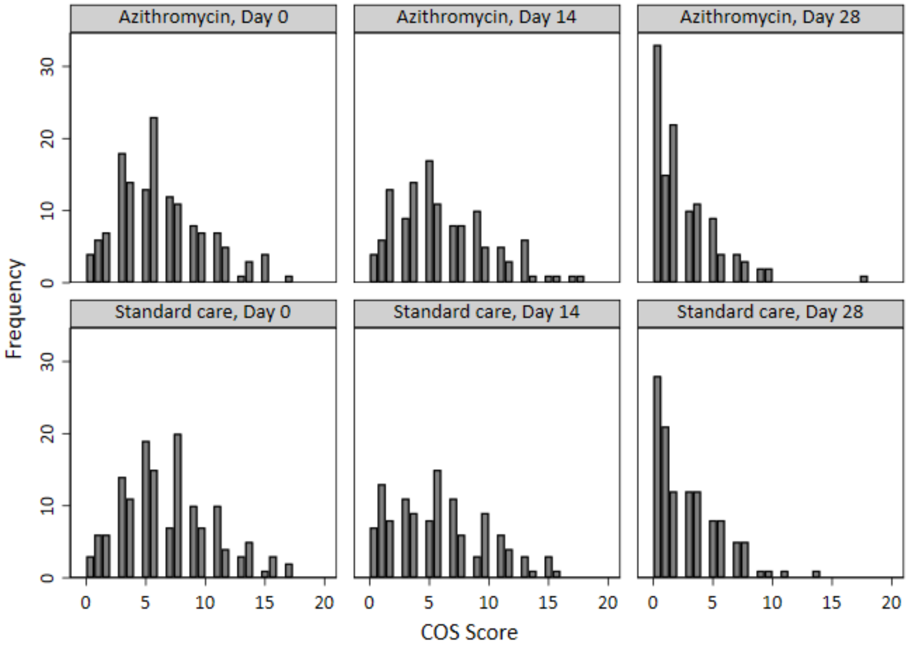
Azithromycin, Day 0
Azithromycin, Day 14
Azithromycin, Day 28
Standard care, Day 0
Standard care, Day 14
Standard care, Day 28
Frequency
COS Score

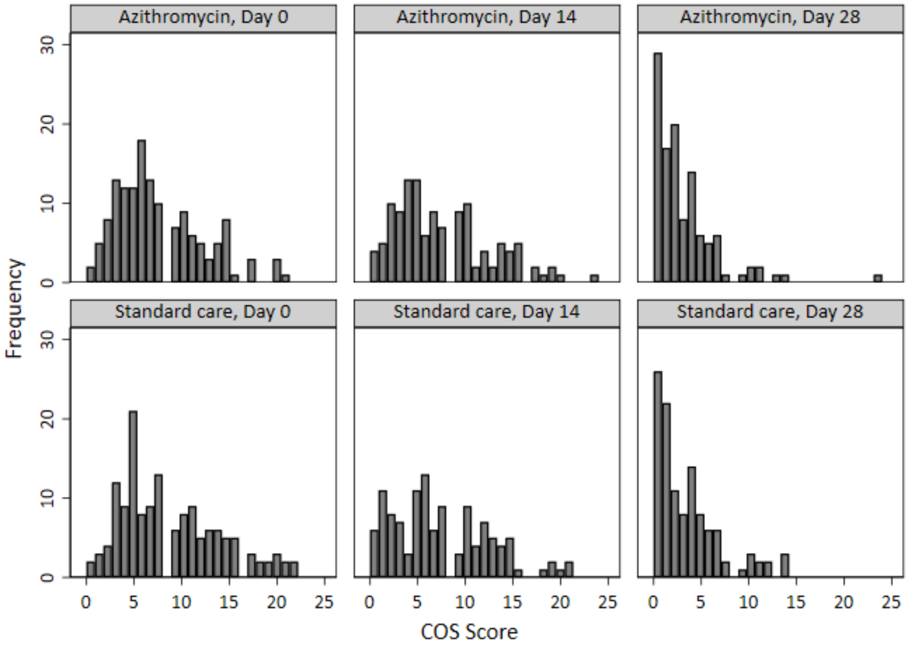
Azithromycin, Day 0
Azithromycin, Day 14
Azithromycin, Day 28
Standard care, Day 0
Standard care, Day 14
Standard care, Day 28
Frequency
COS Score

**Figure S5: Histograms of COS and COS plus scores by treatment group and time point**

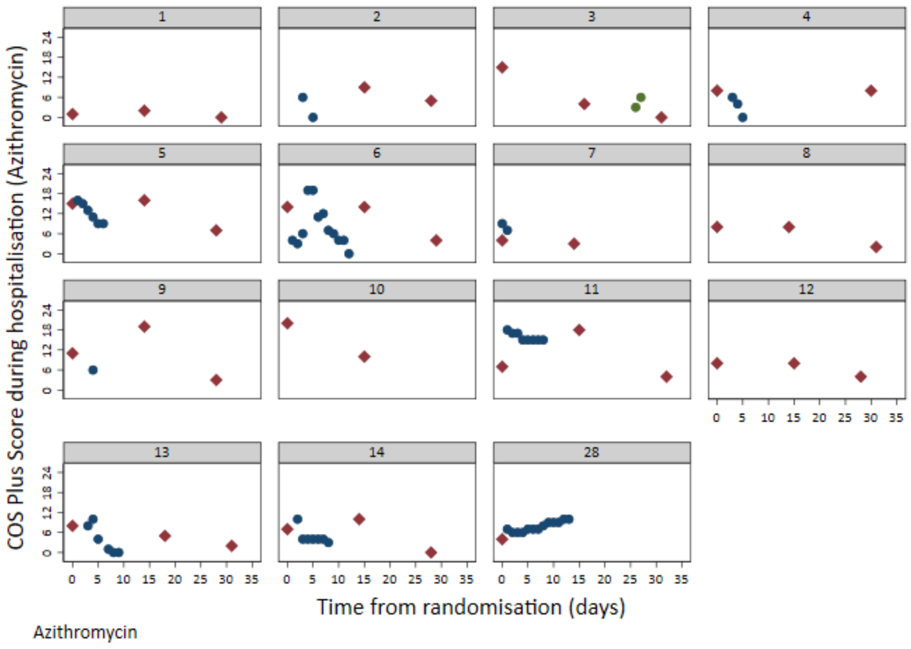

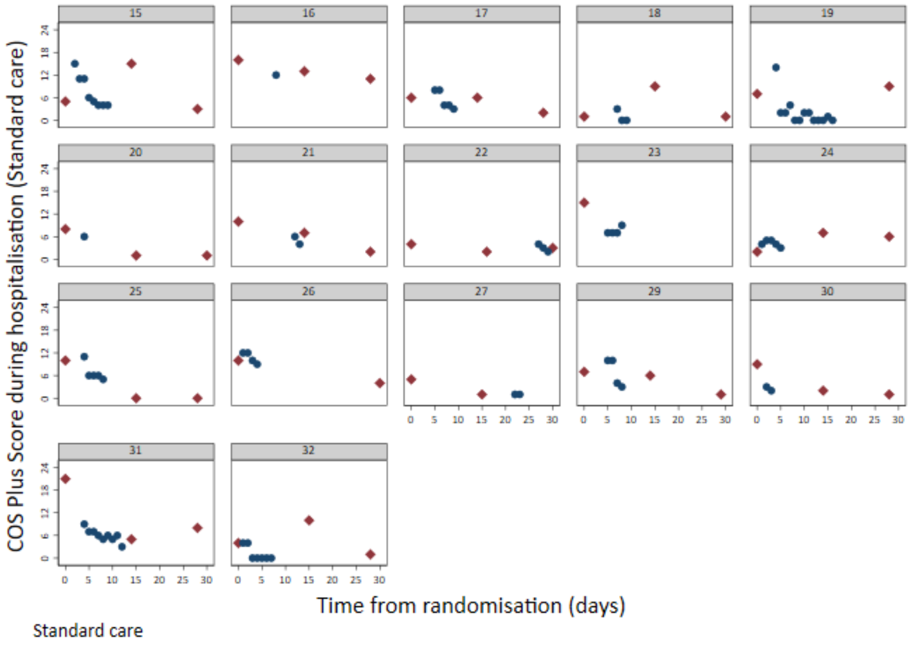

**Figure S6: Scatter plot of the COS PLUS score at each hospital inpatient day for each participant – by intervention groups (ITT population)**

##### 4.3 Complications experienced during hospitalisation

The number of participants experiencing complications during hospitalisation along with details of the complications experienced are summarised by treatment group in Table S20.

**Table S20: Summary of complications experienced during hospitalisation - by treatment group**

| | Azithromycin | | Standard care | |
|---|---|---|---|---|
| | **n** | **%** | **n** | **%** |
| **Total number of participants that had complication(s)** | 3 | 2.1% | 4 | 2.7% |
| **Type of complication** | | | | |
| ***Cryptogenic organising pneumonia (COP)*** | 0 | 0.0% | 2 | 1.4% |
| ***Endocarditis / myocarditis / pericarditis*** | 0 | 0.0% | 1 | 0.7% |
| ***Bacteraemia*** | 1 | 0.7% | 0 | 0.0% |
| ***Hyperglycaemia*** | 2 | 1.4% | 0 | 0.0% |
| ***Hypoglycaemia*** | 0 | 0.0% | 1 | 0.7% |

##### 4.4 Results for per-protocol population

As a sensitivity analysis, the analyses of the primary outcome were repeated for the per protocol population. The following participants were excluded from the per protocol population: (i) those randomised and later found to be ineligible; (ii) those randomised to Azithromycin who did not start treatment; and, (iii) those randomised to standard care who were prescribed a macrolide prior to the day 14 follow-up or hospitalisation (which ever came first). Details of the number and proportion of participants in the per protocol population hospitalised, receiving level 2 or level 3 ventilation, and who died are summarised by treatment group and overall (Table S21). The proportion of participants who were hospitalised or died are compared between the two treatment groups using unadjusted, adjusted, and fully adjusted models with both odds ratios and risk differences presented (Table S22). In addition, supporting analyses using time-to-event methodology were also undertaken with results in Table S22.

**Table S21: Summary of primary outcome population by intervention groups (PP population)**

| | Azithromycin | | Standard Care | | Total | |
|---|---|---|---|---|---|---|
| | **n/N** | **%** | **n/N** | **%** | **n/N** | **%** |
| **Total hospitalised** | 14/142 | 9.9% | 16/144 | 11.1% | 30/286 | 10.5% |
| Received level 2 ventilation* | 1/142 | 0.7% | 1/144 | 0.7% | 2/286 | 0.7% |
| Received level 3 ventilation* | 0/142 | 0.0% | 0/144 | 0.0% | 0/286 | 0.0% |
| Did not receive level 2 or level 3 ventilation | 13/142 | 9.2% | 15/144 | 10.4% | 28/286 | 9.8% |
| **Total died**** | 1/142 | 0.7% | 1/144 | 0.7% | 2/286 | 0.7% |

** Note: Participants could have received both level 2 and level 3 ventilations during hospitalisation.*

*** Note: Both deaths occurred following hospitalisation.*

**Table S22: Primary outcome by intervention groups**

| | Azithromycin | | Standard Care | | | Comparison of proportions | | Time-to-event* |
|---|---|---|---|---|---|---|---|---|
| | n/N | % | n/N | % | | OR (95%CI), p-val | RD (95%CI) | |
| **PP** | 14/142 | 9.9% | 16/144 | 11.1% | **Unadjusted** | 0.88 (0.41, 1.87), 0.73 | -1.3% (-8.4%, 5.8%) | 0.80 |
| | | | | | **Adjusted**** | 0.89 (0.41, 1.92), 0.77 | -1.0% (-8.0%, 6.0%) | 0.95 (0.46, 1.97), 0.89 |
| | | | | | **Fully Adjusted***** | 0.89 (0.40, 1.95), 0.76 | -1.4% (-8.3%, 5.6%) | 0.99 (0.49, 2.00), 0.99 |

*Note: *Comparisons made using a log-rank test (unadjusted) and a Cox's proportional hazards model (adjusted).*
*** Adjust for stratification factors: centre, hypertension, diabetes, and sex.*
**** Adjust for stratification factors and other important prognostic variables: centre, hypertension, diabetes, sex, and age ≥65 years, presence of chronic lung disease, and treatment for cancer.*

##### 4.5 Pre-specified Subgroup Analysis

A forest plots of treatment effects by presence of hypertension, presence of diabetes, sex, and age (<65 vs. ≥65) is provided in Figure S7.

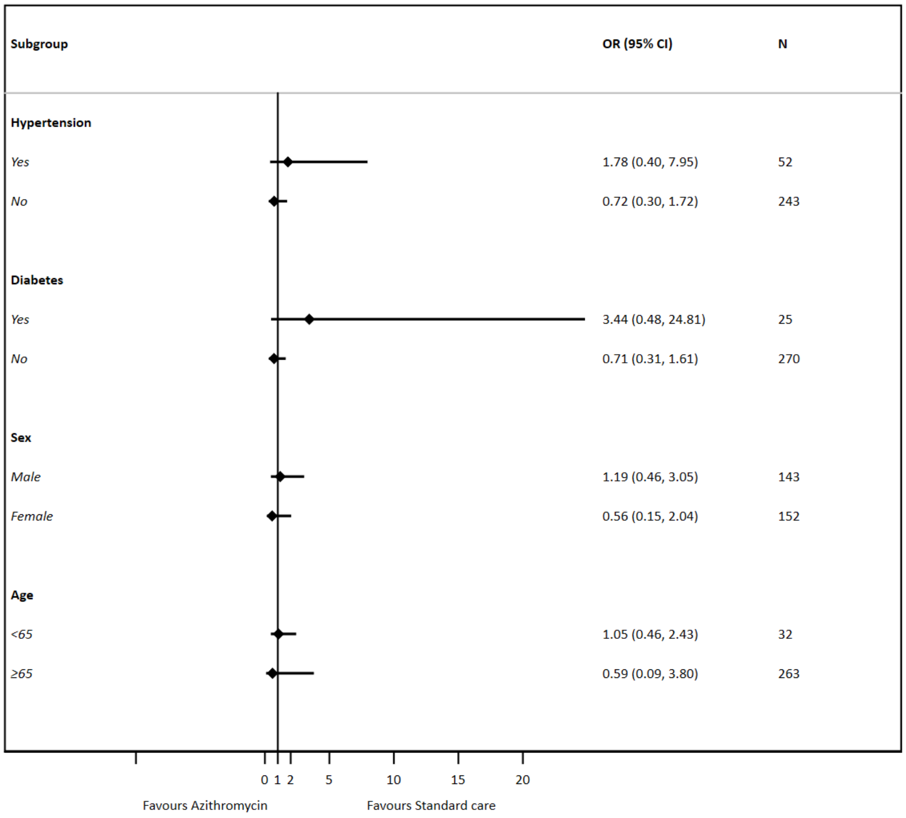

Figure S7: Forest plot of treatment effects in subgroups

##### 4.6 Additional analysis – Antibiotic usage during follow-up

The number and proportion of participants reporting taking antibiotics (other than the study medication) during the follow-up period is summarised by treatment group and overall in Table S23.

**Table 23: Antibiotics (except trial medication) during follow-up**

| | Azithromycin | | Standard Care | | Total | |
|---|---|---|---|---|---|---|
| | n | % | n | % | n | % |
| **Received antibiotics** | 23 | 15.7% | 38 | 25.7% | 61 | 20.7% |
